# Regional Variations in Burden of Chronic Obstructive Pulmonary Disease

**DOI:** 10.1101/2025.01.07.25320159

**Authors:** Jeenat Mehareen, Kayly Choy, Yiwei Yin, Don D. Sin, Mohsen Sadatsafavi

**Author notes:** **Corresponding Author:** Jeenat Mehareen, MA PhD Student, Faculty of Pharmaceutical Sciences University of British Columbia.

## Abstract

**Background:** Chronic Obstructive Pulmonary Disease (COPD) imposes a significant burden on individuals and communities. While differences in such burden are often studied across countries and healthcare systems, differences within jurisdictions have not been well evaluated. The aim of this study was to assess the trends of, and factors explaining, regional variability in the burden of COPD within a well-defined population with publicly funded healthcare system.

**Methodology:** We used population-based health records of people aged 35 years or older diagnosed with COPD across the 16 health regions of British Columbia, Canada (2010–2020). The primary outcomes were prevalence, incidence, all-cause hospital admissions, and all-cause mortality, while secondary outcomes were COPD- and cardiovascular disease (CVD)-specific hospitalization and mortality. We used generalized linear models to examine how outcomes varied by regions and changed over time, controlling for sex, age, socioeconomic status, and rural/urban residence.

**Results:** Over the 11-year study period, there were 312,014 individuals diagnosed with COPD (48.1% female, mean baseline age: 68.2 years). Across the province, standardized prevalence and all-cause mortality remained relatively stable during the study period, whereas incidence and all-cause hospitalization declined. There were up to a three-fold difference in standardized incidence and prevalence, and up to a two-fold difference in standardized all-cause hospitalization and mortality across regions (all p<0.05). These differences remained significant after controlling for case mix. Among COPD patients, both CVD-specific hospitalization and mortality were higher compared to COPD-specific hospitalization and mortality.

**Conclusion:** Despite being governed by the same universal healthcare system, difference in burden of COPD across geographic regions in this Canadian province was significant. Heterogeneity was more prominent in incidence and prevalence, compared with hospitalization and mortality, suggesting that the variation in the process of care leading to diagnosis is more substantial than variation in COPD outcomes.

## INTRODUCTION

Chronic Obstructive Pulmonary Disease (COPD) is a common chronic disease of the airways affecting about two million Canadians^1^, a number that is an underestimate as many individuals remain undiagnosed^2^. It is the sixth leading cause of death in Canada^3^, and the first reason for medical hospitalization after childbirth, with 70,000 admissions in the 2022-2023 fiscal year alone^4^. Relative to its burden on individuals and healthcare systems^5,6^, COPD garners considerably less attention than other chronic diseases in public awareness, health policymaking, and research^7,8^. Because of such under-attention, there might be significant care gaps across the continuum of healthcare, from prevention of airflow obstruction to timely diagnosis to the reduction of adverse outcomes such as exacerbation and mortality. These gaps can affect individuals and communities by various degrees and can compound the already substantial variability in the natural course of COPD.

The path to, and outcomes of, COPD can significantly vary by geographic region^9,10^. A recent rapid review concluded that among various factors such as age, socio-economics status, sex, geography, and ethnicity or race, geography significantly contributed to variation in access to care and health outcomes^11^. Canada has a public healthcare system that is administered provincially. While it is expected that COPD outcomes are varied across provinces due to administrative differences, the universal delivery of care within each province should theoretically narrow the variability in care and outcomes. Unfortunately, evidence on the geographic variability of COPD burden within health systems in lacking, with most studies having focused on trends across the entire population^12–14^. A study from Ontario documented consistently lower incidence, prevalence, healthcare services use, and mortality in the urban and suburban areas of the province^15^. In Alberta, rates and health care use differ by region, with more than two times high prevalence in North Zone compared to Calgary Zone^16^.

British Columbia (BC) is the third-largest Canadian province with a diverse population of around 5.6 million (as of 2024)^17^. In BC, an estimated 140,000 people are diagnosed with COPD, representing about 6% population of ≥45 years of age^18^. BC has a publicly funded healthcare system with complete coverage of healthcare encounter records for its entire population. This, combined with its geographic spread, makes it an ideal setting to study within-system variations in COPD outcomes.

The primary objective of this study was to determine the extent of heterogeneity in prevalence, incidence, all-cause hospitalization and all-cause mortality rates of COPD across geographic areas, and if such heterogeneity could be explained by differences in patient characteristics. The secondary objective was to examine the trends of COPD outcomes across geographic regions.

## METHODS

### Data sources

We used population-based administrative health databases of BC from 1997–2022. The study received ethics approval from the University of British Columbia’s Human Ethics Board (H23-00607). Access to data provided by Data Stewards is subject to approval but can be requested for research projects through Data Stewards or their designated service providers. The following datasets were used: Vital Statistics–Deaths which contain birth and death records, Consolidation files which contain basic demographics such as age and sex, postal code of residence, and registration status with the health system, Hospital Separations data set which includes records of all hospital admissions, and Medical Services Plan which contains information on every outpatient health-service encounter including physician visits^19^. All inferences, opinions, and conclusions drawn in this publication are those of the author(s), and do not reflect the opinions or policies of the Data Steward(s).

### Study Design

This was a retrospective cohort study that included adults aged 35 years and above with a diagnosis of COPD. Individuals were classified as having COPD based on a validated case definition^20^: if they had any of the International Classification of Diseases (ICD), 9th revision (ICD-9) codes of 491, 492, 496, 493.2, or 10th revision (ICD-10) codes of J41, J42, J43, and J44 in the Hospital Separations dataset, or if they had two outpatient visits with the above-mentioned ICD codes within a 24-month rolling period. This case definition has demonstrated a specificity of 91.5% and a positive predictive value of 72.6% in previous chart review studies^20^.

The study period was from January 1, 2010, to December 31, 2020. While we used the data before 2010 as look-back window to identify prevalent COPD patients^21^, we did not include any data before 2010 as the focus was on studying recent trends. We also could not use the last two years of the data as the two-year sliding time window required for case definition means case detection in the last two years will be incomplete. We defined the *index year* as the first year of COPD diagnosis within the study period for incidence cases (those diagnosed at or after 2010), or the first year of the study (2010) for prevalent cases who were diagnosed before 2010. Patients were followed from the index year until any of the following occurred: death, end of registration with healthcare system, last health resource use of any type, or end of the study period. Individuals with less than one year of follow-up were excluded, unless they died within the first year. *Supplementary Material Figure A1* presents a schematic depiction of the study design.

### Exposure and Outcomes

The main exposure was geographic region, defined as the Health Services Delivery Area (HSDA)^22^. The 16 HSDAs in BC are geographically distinct areas that are the focus of planning, reporting, and assessment by health policymakers, making them an ideal geographic unit for the study of heterogeneity in care. The largest HSDA is Fraser South (N=784,977; population density of 937.8/km^2^)^23^, while the smallest one is Northeast region (N=67,885. population density of 0.4/km^2^)^24^.

#### Outcomes

The four co-primary outcomes were COPD incidence, COPD prevalence, all-cause hospitalization among COPD patients, and all-cause mortality among COPD patients. We note that the first two outcomes pertain to the risk of developing the disease, therefore representing the process of care leading to diagnosis (including preventive and diagnostic care), while the others relate to the outcomes among diagnosed patients. Secondary outcomes were COPD- and cardiovascular-disease (CVD)- specific hospitalization and mortality among COPD patients.

#### Prevalence and Incidence

We calculated prevalence and incidence for BC and for each region (HSDA), across the entire study period (primary objective) and for each year (secondary objective) using sex- and age- (within 5 years age bands) population estimates for each region^25^. Period prevalence for a region was defined as total number of individuals 35 years or older who had COPD at any time during the study period, divided by the total number of individuals in the same age group in the same region during the study period. Cumulative incidence was defined as the number of individuals who were diagnosed with COPD for the first time during this period over the total person-years (PY) at risk (those 35 years of age or older who did not have COPD) for each region. For the secondary objective, these outcomes were calculated separately for each region in each calendar year.

#### Hospitalization and Mortality

We calculated cumulative all-cause hospitalization and all-cause mortality among COPD patients as the total number of, respectively, all-cause admissions and deaths, within a given region over the total PY of time patients were diagnosed with COPD in the same region (primary objective). For the secondary objective, these rates were calculated separately by using sex- and age-specific COPD populations for each region for each calendar year.

Hospitalization and mortality were further stratified into COPD-specific and CVD-specific rates (secondary outcomes), with COPD defined as above-mentioned ICD coded, and CVD defined using ICD-9 codes 410-438 and ICD-10 codes I00-I99.

### Statistical Analysis

For all outcomes, we applied direct age and sex standardization with BC population data from Statistics Canada’s 2016 census^26^. Heterogeneity in each outcome across regions by year was visualized through box plots. We reported descriptive statistics for every outcome to present annual data across the entire observational period, stratified by HSDA. All analyses were performed using SAS (version 9.4; Cary, NC, USA), and R (V. 4.2.2, R Foundation). The significance level α = 0.05 was considered throughout the analysis.

To assess to what extent heterogeneity by region was due to differences in case-mix, we used regression models, with each outcome as the dependent variable and region (dummy coded HSDAs, with Fraser South – the largest HSDA – as the reference category) as the independent variable of interest. Regression models were stratified by age groups (35-49 years, 50-64 years, 65-79 years, and 80+ years) and sex, and were further adjusted for the area of residence (urban/rural) and socio-economic status (SES - classified into neighborhood income quintiles with Q5 as the wealthiest quintile)^27^. To account for overdispersion and the clustered nature of the data (data from the same region divided into subgroups and years), we employed generalized linear models with a negative binomial distribution and a log link function^28^.

This model generated adjusted rate ratios (RR) and corresponding 95% confidence intervals (CI) for the association between each region and each outcome. The null hypothesis that there is no heterogeneity in an outcome across regions was assessed via the likelihood ratio test: comparing the log-likelihood of the above model with a similar model without region as an explanatory variable.

The model for the analysis of trends was the same as the main model described in the previous section, with the addition of explanatory variables representing calendar year, and its first-order interaction with variables representing regions. The coefficient for the year in this model captured the common temporal trends across all regions, while the coefficient for the year-by-region term captured the trend specific to each region (with Fraser South taken as the reference category).

## RESULTS

Over the 11 years (2010–2020), there were 312,014 unique individuals who satisfied the case definition of COPD. ***Table 1*** provides demographic and geographic characteristics of this cohort, overall and by region. The mean follow-up years across all regions ranged from 4.7 to 6.1 years (overall mean= 5.8 years, SD=3.6). Females constituted 48.1% of individuals. The average age at diagnosis was 68.2 years (SD=12.8), with minor variations across regions. Most patients resided in urban areas (83.9%) and were aged between 65-79 years (39.0%) while those aged 35-49 years constitute the smallest group (7.6%). 25.2% of participants were in the lowest SES quintile (range across regions: 10.5% to 32.5%) and 15.7% (range across regions: 9.5% to 35.7%) in the highest quintile.

**Table 1:** Characteristics of the COPD Cohort.

| Variables | Value | Fraser<br>South | Okanagan | Fraser<br>North | Vancouver | Central<br>Vancouver<br>Island | South<br>Vancouver<br>Island | Thompson<br>Cariboo<br>Shuswap | Fraser<br>East | North<br>Shore-Coast<br>Garibaldi | North<br>ern<br>Interior | North<br>Vancouver<br>Island | Richmond | Kootenay<br>Boundary | East<br>Kootenay | Northwest | North<br>east | Unknown/<br>Missing |
| --- | --- | --- | --- | --- | --- | --- | --- | --- | --- | --- | --- | --- | --- | --- | --- | --- | --- | --- |
| **Total**<br>**(2010-2020)** | 312,014 | 37606 | 37449 | 34515 | 33131 | 26550 | 24005 | 23683 | 20688 | 16748 | 11503 | 11369 | 8283 | 8136 | 7580 | 5196 | 5133 | 439 |
| **Follow-up**<br>**years, mean**<br>**(SD)** | 5.8<br>(3.6) | 5.9<br>(3.6) | 5.7<br>(3.6) | 6.1<br>(3.6) | 6.0<br>(3.6) | 5.9<br>(3.6) | 5.6<br>(3.6) | 5.8<br>(3.6) | 5.8<br>(3.6) | 5.7<br>(3.6) | 5.5<br>(3.6) | 5.7<br>(3.5) | 6.0<br>(3.7) | 5.9<br>(3.6) | 5.6<br>(3.5) | 5.7<br>(3.6) | 5.6<br>(3.6) | 4.7<br>(3.7) |
| **Sex** |  |  |  |  |  |  |  |  |  |  |  |  |  |  |  |  |  |  |
| Female, % | 149957<br>(48.1) | 18362<br>(48.8) | 18276<br>(48.8) | 16886<br>(48.9) | 14503<br>(43.8) | 12953<br>(48.8) | 12113<br>(50.5) | 11322<br>(47.8) | 10159<br>(49.1) | 8361<br>(49.9) | 5485<br>(47.7) | 5411<br>(47.6) | 3891<br>(46.9) | 3686<br>(45.3) | 3655<br>(48.2) | 2287<br>(44.0) | 2409<br>(46.9) | 198<br>(45.1) |
| Male, % | 162057<br>(51.9) | 19244<br>(51.2) | 19173<br>(51.2) | 17629<br>(51.1) | 18628<br>(56.2) | 13597<br>(51.2) | 11892<br>(49.5) | 12361<br>(52.2) | 10529<br>(50.9) | 8387<br>(50.1) | 6018<br>(52.3) | 5958<br>(52.4) | 4392<br>(53.0) | 4450<br>(54.7) | 3925<br>(51.8) | 2909<br>(56.0) | 2724<br>(53.1) | 241<br>(54.9) |
| **Age at**<br>**baseline**<br>**(years)†,**<br>**mean (SD)** | 68.2<br>(12.8) | 68.0<br>(13.1) | 69.8<br>(12.4) | 67.7<br>(13.2) | 68.0<br>(13.8) | 68.1<br>(12.4) | 70.2<br>(13.0) | 67.9<br>(12.0) | 67.8<br>(12.7) | 69.7<br>(12.9) | 65.1<br>(12.3) | 67.8<br>(11.7) | 68.8<br>(13.0) | 68.0<br>(11.9) | 68.2<br>(12.0) | 65.3<br>(11.8) | 63.5<br>(12.7) | 68.4<br>(15.3) |
| **Age group, %** |  |  |  |  |  |  |  |  |  |  |  |  |  |  |  |  |  |  |
| 35-49 years | 23679<br>(7.6) | 3181<br>(8.5) | 2136<br>(5.7) | 3076<br>(8.9) | 3210<br>(9.7) | 1848<br>(7.0) | 1345<br>(5.6) | 1486<br>(6.3) | 1659<br>(8.0) | 1072<br>(6.4) | 1231<br>(10.7) | 680<br>(6.0) | 589<br>(7.1) | 459<br>(5.6) | 463<br>(6.1) | 470<br>(9.1) | 721<br>(14.1) | 53<br>(12.1) |
| 50-64 years | 100219<br>(32.1) | 12149<br>(32.3) | 10708<br>(28.6) | 11354<br>(32.9) | 10636<br>(32.1) | 8558<br>(32.2) | 7163<br>(29.8) | 7807<br>(33.0) | 6734<br>(32.6) | 4750<br>(28.4) | 4341<br>(37.7) | 3812<br>(33.5) | 2657<br>(32.1) | 2813<br>(34.6) | 2478<br>(32.7) | 2019<br>(38.9) | 2101<br>(40.9) | 139<br>(31.7) |
| 65-79 years | 121608<br>(39.0) | 14165<br>(37.7) | 15537<br>(41.5) | 12735<br>(36.9) | 11589<br>(35.0) | 10839<br>(40.8) | 8909<br>(37.1) | 10097<br>(42.6) | 8181<br>(39.5) | 6642<br>(39.7) | 4472<br>(38.9) | 4930<br>(43.4) | 3091<br>(37.3) | 3364<br>(41.4) | 3189<br>(42.1) | 2051<br>(39.5) | 1686<br>(32.9) | 131<br>(29.8) |
| >=80 years | 66508<br>(21.3) | 8111<br>(21.6) | 9068<br>(24.2) | 7350<br>(21.3) | 7696<br>(23.2) | 5305<br>(20.0) | 6588<br>(27.4) | 4293<br>(18.1) | 4114<br>(19.9) | 4284<br>(25.6) | 1459<br>(12.7) | 1947<br>(17.1) | 1946<br>(23.5) | 1500<br>(18.4) | 1450<br>(19.1) | 656<br>(12.6) | 625<br>(12.2) | 116<br>(26.4) |
| **Geographic area, %** |  |  |  |  |  |  |  |  |  |  |  |  |  |  |  |  |  |  |
| Rural | 50314<br>(16.1) | 47<br>(0.1) | 9543<br>(25.5) | 10<br>(0.0) | 6<br>(0.2) | 5758<br>(21.7) | 829<br>(3.5) | 9431<br>(39.8) | 2513<br>(12.2) | 3939<br>(23.5) | 2666<br>(23.2) | 2648<br>(23.3) | <5<br>(0.0) | 4339<br>(53.3) | 4352<br>(57.4) | 2110<br>(40.6) | 2102<br>(41.0) | 2144<br>(48.8) |
| Urban | 261700<br>(83.9) | 37559<br>(99.9) | 27906<br>(74.5) | 34505<br>(100.0) | 33125<br>(99.8) | 20792<br>(78.3) | 23176<br>(96.5) | 14252<br>(60.2) | 18175<br>(87.9) | 12809<br>(76.5) | 8837<br>(76.8) | 8721<br>(76.7) | 8281<br>(99.9) | 3797<br>(46.7) | 3228<br>(42.6) | 3086<br>(59.4) | 3031<br>(59.0) | 225<br>(51.2) |
| **Socio-economic status, %** |  |  |  |  |  |  |  |  |  |  |  |  |  |  |  |  |  |  |
| Quintile 1 | 78520<br>(25.2) | 7702<br>(20.5) | 10676<br>(28.5) | 8008<br>(23.2) | 10760<br>(32.5) | 6287<br>(23.7) | 6134<br>(25.6) | 5821<br>(24.6) | 6337<br>(30.6) | 1766<br>(10.5) | 3023<br>(26.3) | 3099<br>(27.3) | 2535<br>(30.6) | 2505<br>(30.8) | 1625<br>(21.4) | 1431<br>(27.5) | 753<br>(14.7) | 58<br>(13.2) |
| Quintile 2 | 66425<br>(21.3) | 8700<br>(23.1) | 8690<br>(23.2) | 6976<br>(20.2) | 7050<br>(21.3) | 5154<br>(19.4) | 4965<br>(20.7) | 5778<br>(24.4) | 4644<br>(22.5) | 3256<br>(19.4) | 2133<br>(18.5) | 2431<br>(21.4) | 1492<br>(18.0) | 1788<br>(22.0) | 1614<br>(21.3) | 1011<br>(19.5) | 695<br>(13.5) | 48<br>(10.9) |
| Quintile 3 | 59311<br>(19.0) | 7663<br>(20.4) | 6560<br>(17.5) | 7959<br>(23.1) | 5623<br>(17.0) | 5067<br>(19.1) | 4221<br>(17.6) | 5029<br>(21.2) | 3737<br>(18.1) | 2564<br>(15.3) | 2060<br>(17.9) | 2164<br>(19.0) | 1815<br>(21.9) | 1407<br>(17.3) | 1671<br>(22.0) | 876<br>(16.9) | 847<br>(16.5) | 48<br>(10.9) |
| Quintile 4 | 55118<br>(17.7) | 7553<br>(20.1) | 6487<br>(17.3) | 7443<br>(21.6) | 4121<br>(12.4) | 4609<br>(17.4) | 4051<br>(16.9) | 3923<br>(16.6) | 3123<br>(15.1) | 3043<br>(18.2) | 2480<br>(21.6) | 1798<br>(15.8) | 1571<br>(19.0) | 1383<br>(17.0) | 1430<br>(18.9) | 1028<br>(19.8) | 1019<br>(19.9) | 56<br>(12.8) |
| Quintile 5 | 48541<br>(15.6) | 5943<br>(15.8) | 4603<br>(12.3) | 4087<br>(11.8) | 4564<br>(13.8) | 5185<br>(19.5) | 4343<br>(18.1) | 2256<br>(9.5) | 2700<br>(13.1) | 5978<br>(35.7) | 1760<br>(15.3) | 1721<br>(15.1) | 806<br>(9.7) | 1047<br>(12.9) | 1190<br>(15.7) | 615<br>(11.8) | 1669<br>(32.5) | 74<br>(16.9) |
| Unknown | 4099<br>(1.3) | 45<br>(0.1) | 433<br>(1.2) | 42<br>(0.1) | 1013<br>(3.1) | 248<br>(0.9) | 291<br>(1.2) | 876<br>(3.7) | 147<br>(0.7) | 141<br>(0.8) | 47<br>(0.4) | 156<br>(1.4) | 64<br>(0.8) | 6<br>(0.1) | 50<br>(0.7) | 235<br>(4.5) | 150<br>(2.9) | 155<br>(35.3) |

*Abbreviations:* COPD, chronic obstructive pulmonary disease; SD, standard deviation

*Supplementary Table A1* provides a snapshot of unadjusted regional variation across all outcomes. The unadjusted period prevalence of COPD over the study period was 6.9%, ranging from a low of 4.2% to a high of 10.2% across regions (Coefficient of Variation [CV]=24.9%). Unadjusted cumulative incidence across the province was 569.2/100,000 PY, which varied from 322.9 to 914.4 across regions (CV=29.3%). Rankings of regions for period prevalence and cumulative incidence were consistent. Hospitalization and mortality also varied, albeit not as substantially as prevalence and incidence. Hospitalization varied from 62,929.3 to 89,598.1/100,000PY (province-wide value: 73,136.1, CV=10.5%), while mortality varied from 4,229.6 to 5,653.9/100,000PY (province-wide rate: 5,037.3, CV=8.3%). While there was some overlap in the extremes of mortality and hospitalization ranking across regions, these did not align as closely with the rankings for prevalence and incidence (*Supplementary Table A1)*.

***Figure 1*** presents the adjusted RR and 95% CIs for outcomes across regions from the adjusted models, with Fraser South as the reference group (with an RR of 1). Detailed estimates are provided in *Supplementary Tables A2–A5*. The likelihood ratio test indicated the presence of significant heterogeneity across regions for all outcomes (p<0.01 for all). For all outcomes, the adjusted model revealed remaining variation in RRs after controlling for case mix. Notably, more regions had statistically significant RRs, relative to the reference region, for prevalence and incidence compared to hospitalization and mortality. The region with the highest prevalence had adjusted RR approximately 1.74 times higher than the region with the lowest (*Supplementary Table A2*). Adjusted results for incidence showed even greater variation, with the highest RR being 2.47 times higher than the lowest (*Supplementary Table A3*). The highest RR of all-cause hospitalization and all-cause mortality were 1.24 and 1.40 times higher than the region with lowest RR, respectively (*Supplementary Tables A4–A5*). Compared to Fraser South (RR = 1), most northern regions (Northern Interior and Northeast) exhibited significantly higher RRs for all outcomes except hospitalization.

**Figure 1:**
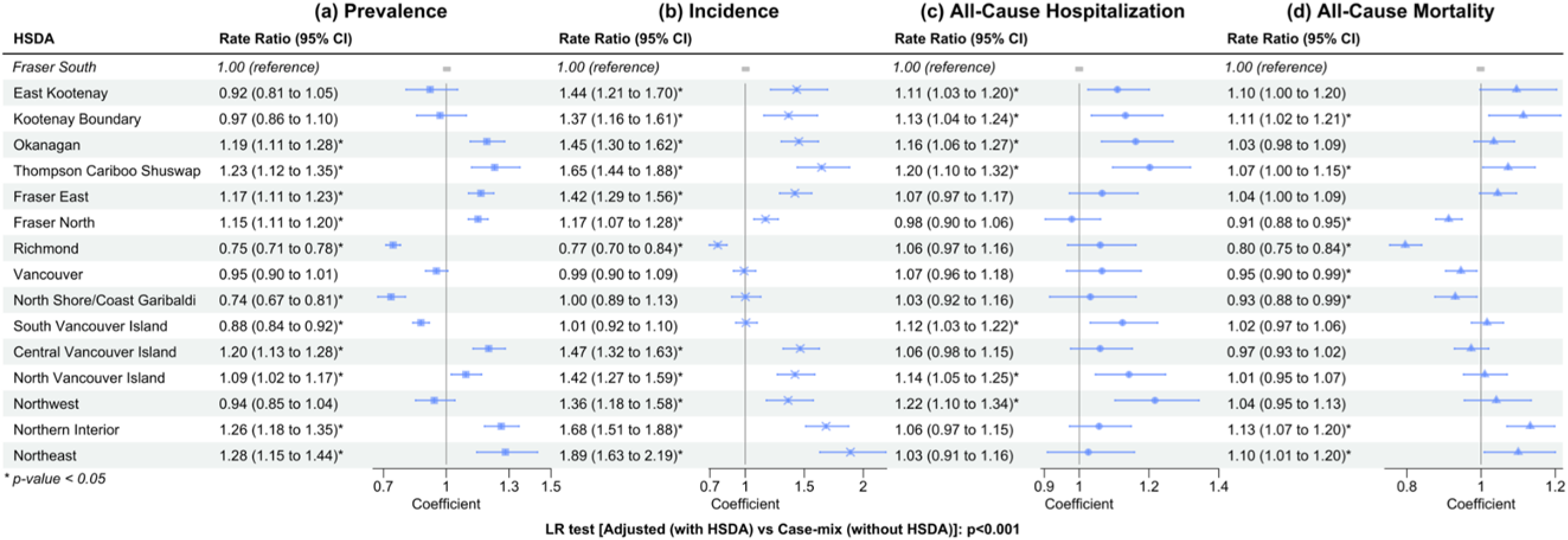
Rate ratios presented in a forest plot for the adjusted models for each primary outcome Prevalence (a), Incidence (b), All-cause hospitalization (c) and All-cause mortality (d) Confidence bands correspond to 95% CI. Adjusted variables: Sex, Socio-Economic Status, Area of Residence, Age group. Abbreviations: LR test: Likelihood-Ratio test; HSDA: Health Service Delivery Area, CI: Confidence Interval

### Trends in outcome*s*

***Figure 2*** provides the standardized annual rates by year and regions for all the outcomes. While incidence and all-cause hospitalizations showed generally declining trends, prevalence and all-cause mortality largely remained stable during the study period.

**Figure 2:**
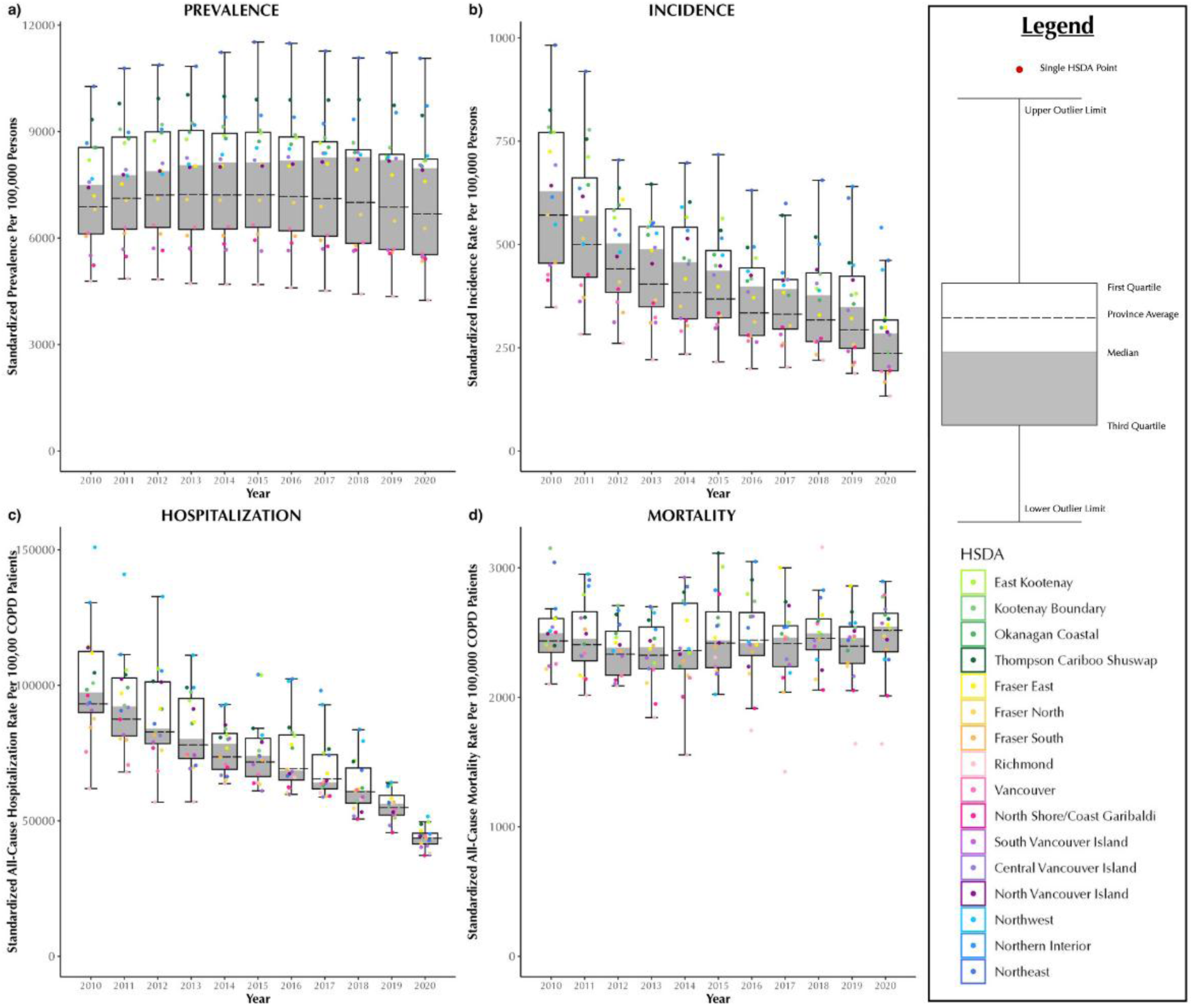
Annual sex and age standardized chronic obstructive pulmonary disease prevalence (a), incidence (b), All-cause Hospitalization (c), and All-Cause Mortality (d) rates in British Columbia, Canada, from 2001 to 2020, stratified by Health Service Delivery Areas (HSDAs) Abbreviations: COPD: chronic obstructive pulmonary disease; Q1: quartile 1; Q3: quartile 3 Note: Each dot represents a region (HSDA). The horizontal line cutting through the plot is the overall provincial average. The median is the line separating the upper (white) and lower (dark grey) boxes.

The number of adults with COPD (raw prevalence per 100,000) increased from 161,661 (3,620.2) in 2010 to 209,285 (4,043.3) (*Supplementary Table A6)*. However, after sex-and age-standardization, prevalence showed a slight decline (***Figure 2(a)***). The number of new COPD cases decreased from 20,229 in 2010 to 10,801 in 2020, a decline that is also reflected in sex-and age-standardized rates (per 100,000 - ***Figure 2(b)****, Supplementary Table A7*). Standardized rates varied substantially across regions, with up to a three-fold difference in incidence and prevalence across location on average each year.

The number of all-cause hospital admissions rate was 141,580 in 2010. It showed a gradual decline especially from 2015 onwards, with a sharp drop during the COVID-19 pandemic. When sex- and age-standardized, the province-wide rate showed a gradual decline from 2010 to 2020 *(Figure 2 (c), Supplementary Table A8*). In contrast, the number of all-cause deaths increased from 7,871 in 2010 to 10,941 in 2020. Standardized all-cause mortality per 100,000 COPD-patient years did not show any noticeable trend during this time period *(Figure 2 (d), Supplementary Table A9).* Again, regional variation was evident in both outcomes, with up to a two-fold difference in standardized all-cause hospitalization and all-cause mortality rates across regions per year.

For all outcomes, there was statistically significant heterogeneity in time trends across regions (p<0.05).

*Figure 3* shows the relationship between cumulative risk across the entire time period (X-axis), and overall time trend (Y-axis) for each outcome across 16 regions over the study period.

**Figure 3:**
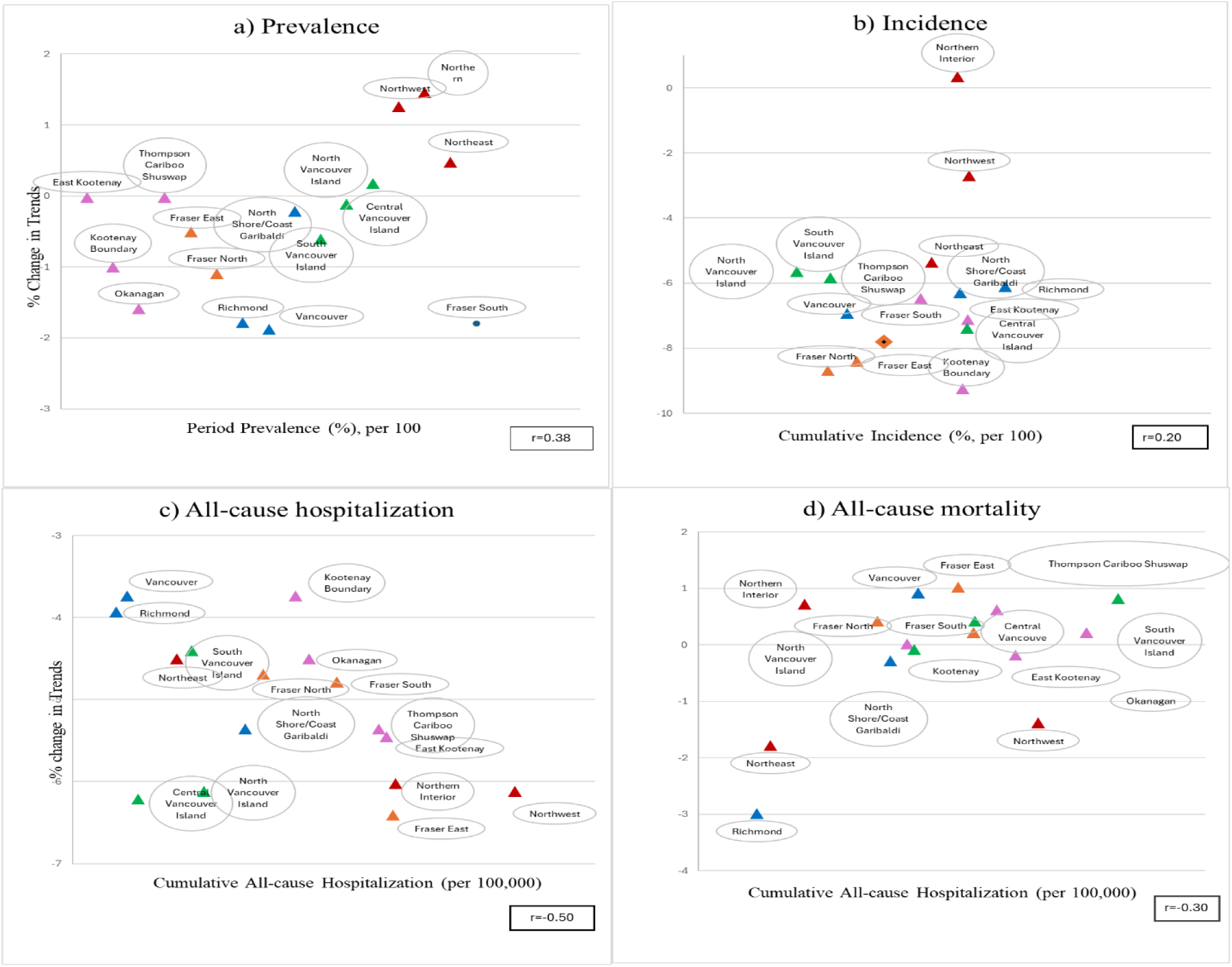
Scatterplots of primary outcomes (X-axis) versus percentage change in time trend (from trend adjusted model, Y-axis) across regions for Prevalence (a), Incidence (b), All-cause Hospitalization (c), and All-cause Mortality (d). Each triangle represents an HSDA, color coded for different health authorities Interior (Pink), Fraser (Orange), Vancouver Coastal (Bue), Vancouver Island (Green) and Northern (Red). The correlation (r) is represented by the Pearson correlation coefficient. Abbreviations: HSDA Health Services Delivery Areas; RR: Rate Ratio

For each additional year, COPD prevalence, incidence, and all-cause hospitalization decreased by approximately 1.8%, 7.8% and 4.8%, after adjusting for region and case-mix (p<0.05) whereas mortality rates showed no overall significant trends. However, there was a variation in trends across HSDAs. The top two panels *(Figure 3(a) and (b))* show that Northern regions (colored in red) stand out in terms of having a high incidence and prevalence as well as an increasing trend. For all-cause hospitalization and mortality, however, northern regions were not any different from others *(Figure 3(c) and (d))*.

### Secondary outcomes

***Figure 4*** presents the sex- and age-standardized rates of COPD- and CVD-related hospitalizations and mortality (per 100,000 COPD patients) across regions over the study period. On average, within the COPD population, CVD-related hospitalization and mortality rates were, respectively, twice and 1.5 times higher than COPD-related hospitalization rates.

**Figure 4:**
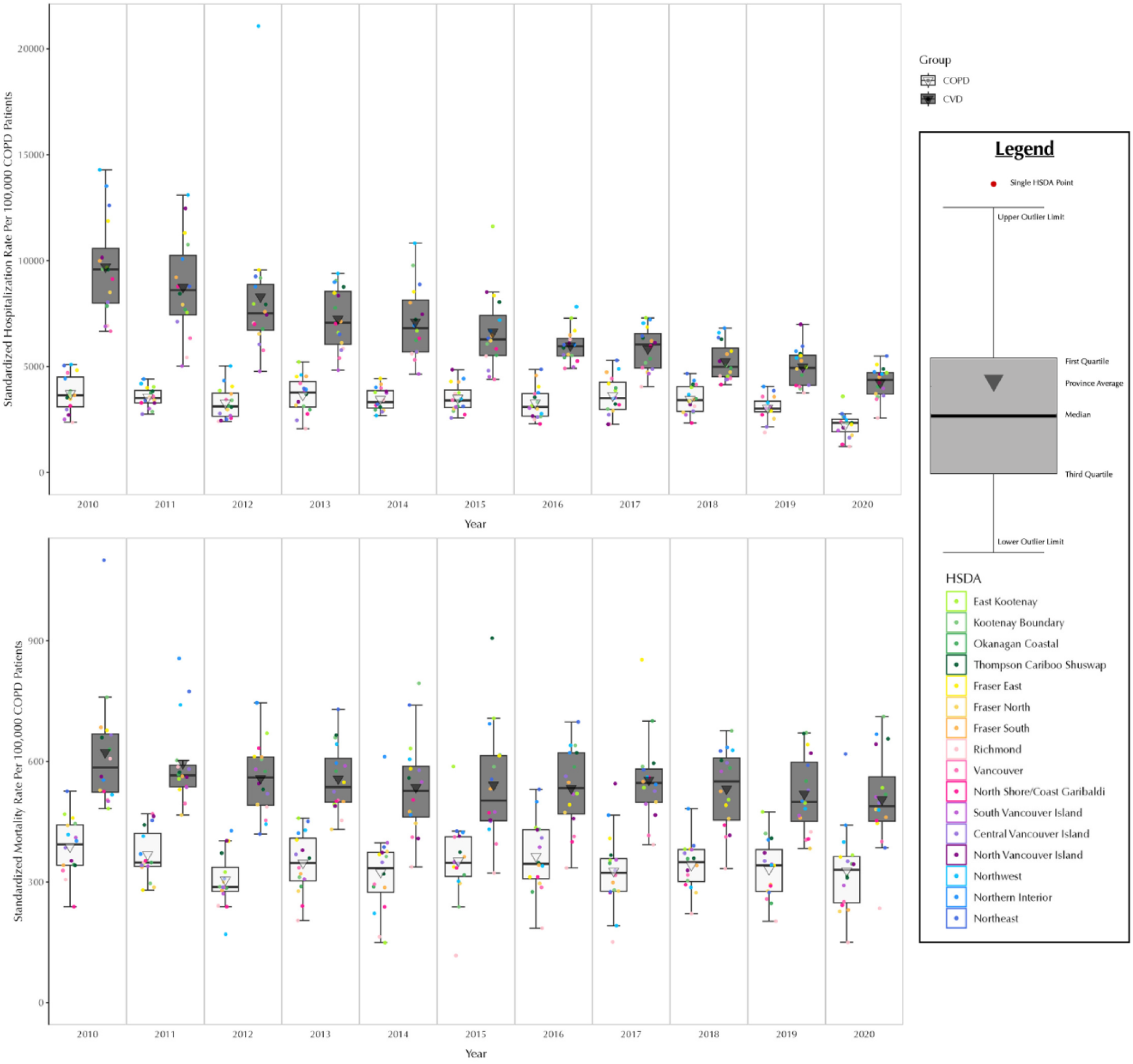
Annual sex and age standardized chronic obstructive pulmonary disease COPD-specific (black boxplot) and CVD-specific (white box plot) rates (per 100,000 COPD patient-year) for all-cause hospitalization (upper panel) and all-cause mortality (lower panel) in British Columbia, Canada, from 2001 to 2020, stratified by Health Service Delivery Area. Abbreviations: COPD: chronic obstructive pulmonary disease; Q1: quartile 1; Q3: quartile 3 Note: Each dot represents a region (HSDA). The horizontal line cutting through the plot is the median. The provincial average is the triangle centered in each box

From 2010 to 2017, the number of COPD-specific hospitalizations increased from 8,379 to 10,527, before declining in more recent years, including the pandemic period. After standardization, the rates showed fluctuations, with a drop in the later years. In contrast, CVD-related hospitalizations showed an overall downward trend *(Supplementary Table A8)*. On average, the standardized rates of COPD and CVD-specific hospitalizations differed up to two-fold across regions. Although COPD and CVD related deaths increased from 2010 to 2020, standardized COPD-specific mortality rates showed no clear trend, aside from a pandemic-related decline (*Supplementary Table A9*). The variation in COPD mortality rates across regions was up to three-fold, whereas CVD-related mortality varied by up to two-fold.

## DISCUSSION

We examined heterogeneity in the risk of developing COPD, manifested as incidence and prevalence, as well as outcomes among COPD patients, namely as all-cause hospitalization and all-cause mortality, over a 11-year period in 16 geographic regions in a large Canadian province. Across the province, standardized prevalence and all-cause mortality remained relatively stable during the study period, whereas incidence and all-cause hospitalization declined. However, there were significant variability in these outcomes across geographic regions. Standardized prevalence and incidence varied up to three-fold, while standardized hospitalization and mortality varied and up-to two-fold, across regions. Differences remained substantial after further adjusting for socioeconomic and rural/urban status. Generally, variation in magnitude of COPD outcomes (all-cause hospitalization and mortality) were less substantial compared to the variation in incidence and prevalence. Furthermore, trend analyses showed that not only COPD outcomes are varied across the health regions, the trajectory of change is also highly variable. Again, variations in trends across regions were more significant for incidence and prevalence, suggesting that variation in care that leads to diagnosis is substantial across regions. Apart from a decline during the COVID-19 pandemic era, COPD-specific hospitalization and mortality showed no discernible trend over time after standardization, with up to two-fold and three-fold variation across regions respectively.

To the best of our knowledge, no large-scale, population-based studies have explored the extent of geographic variations in COPD outcomes and resource use across BC. Previous studies in BC have identified significant regional differences in prevalence of excess weight, tobacco smoking and physical inactivity^29^, multimorbidity^30^, primary care visits for alcohol-attributed diseases^31^, incidence, prevalence, and use of various health care services in airway diseases^32,33^. Despite the fact that BC has a centrally administered public and universal healthcare system, the differences in outcomes across HSDAs were substantial, rivalling in magnitude differences across countries in the burden of COPD^34–36^.

In the trend adjusted model, our results reveal a general decline in COPD in prevalence and incidence trends by 1.8% and 7.8%, respectively. However, northern BC stands out with rising trends in prevalence. This mirrors findings from previous studies^16^, including one from Ontario^15^ and from Alberta^16^, where northern, industrial, and rural areas had higher COPD rates compared to urban areas and areas in the south. Factors such as industrial exposure (e.g., mining, forestry, natural gas), higher smoking rates^29^ compared to Canada wide and province specific rates, and environmental issues^37^ like frequent forest fires^38^ could contribute to the elevated COPD burden in northern BC. The comparatively slower decline in incidence in these regions also highlights the need to address specific risk factors contributing to this growing public health challenge. On the other hand, mortality and hospitalization rates in these regions were not significantly different than other parts of the province. This might indicate that once diagnosed with COPD, there is less variability in care and outcomes across regions. Improved access to care through engagement of nurse practiotioners^39^ or telehealth^40^ likely contributes to the consistency in outcomes. The disparity in preventive care, however, is apparent, which signals that healthcare policymakers and local health authorities should prioritize targeted preventive or early diagnostic interventions. This could include enhancing public health education on COPD risk factors and improving access to preventive services, particularly in underserved and remote areas.

Province-wide, crude hospitalization rates have generally declined over the 11-year period in our study, with all regions experiencing some level of reduction. It appears that the main reason for the steady drop is due to the decline in CVD-specific admissions. In contrast, COPD-specific admissions fluctuated during that time. These results are consistent with a recent Canadian study reporting a 23% decline in all-cause, sex- and age-standardized admissions from 2002 to 2017, though with a contrasting 68% increase in COPD-specific admissions^41^. However, with only 25% of all-cause deaths in our sample being CVD-related, the slight increase in all-cause mortality may reflect the impact of other comorbidities and the COVID-19 pandemic, which overlaps with the final year of the study period.

Our study has several strengths. Unlike insurance claims data^11^ that lack full population coverage, our administrative databases captures the entire population, minimizing selection bias. Additionally, we examined multiple outcomes, enabling us to juxtapose various trends. The availability of long-term health follow-up data further allowed us to analyze temporal patterns. Limitations of this analysis should also be noted. This was a population-based descriptive study that focused on demographic and socioeconomic factors. Some important factors that can contribute to variations in outcomes were not available in our data. These include smoking status^42^ and ambient air quality^43^. Access to such data would have enabled us to assess to what extent between-region variability can be explained. Furthermore, the diagnosis of COPD was based on ICD codes. While the case definition we used is validated, it does not have a perfect diagnostic performance^44^. Importantly, COPD remains an underdiagnoses disease^45^, and as our study was based on a cohort of diagnosed COPD patient, we could not capture variability in the rate of diagnosis. Spirometry is underutilized in COPD diagnosis, and many physicians instead rely on symptom-based assessments, thus missing COPD diagnosis in their early stages of the disease^46^. If regions with poorer outcomes also have lower access to spirometry, this might indicate the variations in the incidence and prevalence of (diagnosed and undiagnosed) COPD might be even more substantial than our results indicate.

In summary, this study reveals significant geographic variability in COPD outcomes, in particular for prevalence and incidence, across BC, a jurisdiction with a publicly funded healthcare system. Clinically, these findings highlight the need for tailored diagnostic and care strategies to address regional differences. From a policy perspective, health planners should be aware of potentially substantial variability in COPD burden and outcome within their jurisdictions. The research underscores the value of identifying when, where, and who needs resources most to guide targeted interventions and optimize resource allocation. The reasons for such variability, beyond demographics, SES, and residential status, are worth exploring.

## Data availability

Access to data provided by Data Stewards is subject to approval but can be requested for research projects through the Data Stewards or their designated service providers. The following data sets were used in this study: (Vital Statistics - Deaths, Consolidation Files, Hospital Separations, MSP Payment Information, PharmaNet). You can find further information regarding these data sets by visiting the PopData project webpage at: https://my.popdata.bc.ca/project_listings/23-081/collection_approval_dates. All inferences, opinions, and conclusions drawn in this publication are those of the author(s), and do not reflect the opinions or policies of the Data Steward(s).

## Authors’ contribution

Conceptualization (Mohsen Sadatsafavi, Jeenat Mehareen), literature review (Kayly Choy, Jeenat Mehareen), methodology (Mohsen Sadatsafavi, Jeenat Mehareen), formal analysis (Jeenat Mehareen), first drafting of the manuscript (Jeenat Mehareen, Mohsen Sadatsafavi), validation (all authors), review, editing and interpretations (all authors). All authors agreed to be accountable for all aspects of the work.

