## Supplementary Materials for "Regional Variations in Burden of Chronic Obstructive Pulmonary Disease"

**Appendices**

Index date

First eligible date when patient entered the cohort following case definition

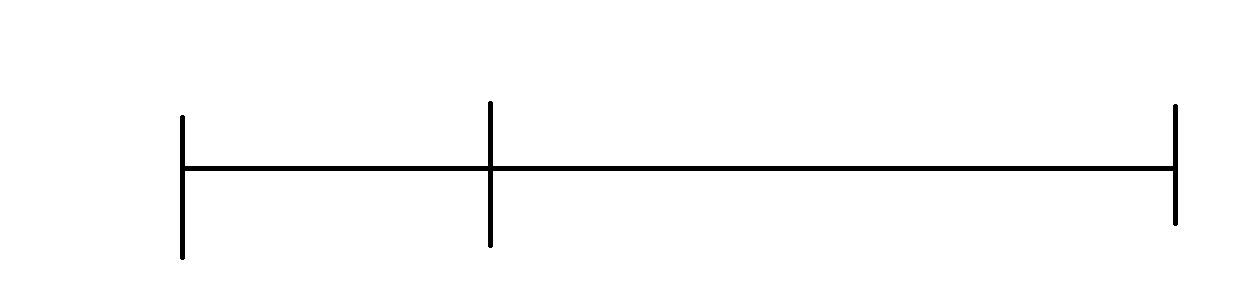

**January 1, 2010**

**December 31, 2020**

**June 30, 2022**

**January 1, 1997**

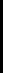

End of observation

Earliest date of death, last date of registration or end of study period

Observation Window

Case Ascertainment Window

### **Figure A1: Retrospective Cohort Study Design**

Note: Results are presented for ‘Observation Window’ period defined as January 1, 2010, to December 31, 2020 (or latest available data

### **Table A1: COPD prevalence, incidence, all-cause hospitalization and all-cause mortality by geographic region, aged 35 or older, BC, 2010 to 2020 (combined)**

| **Health Service Delivery Areas** | | | | | | | | | | | | | | | | | | |
| --- | --- | --- | --- | --- | --- | --- | --- | --- | --- | --- | --- | --- | --- | --- | --- | --- | --- | --- |
| **Outcome** | **Total** | **Fraser South** | **Fraser North** | **Vancouver** | **South Vancouver Island** | **Okanagan** | **Central Vancouver Island** | **North Shore-Coast Garibaldi** | **Fraser East** | **Thompson Cariboo Shuswap** | **Richmond** | **North Vancouver Island** | **Northern Interior** | **Kootenay Boundary** | **East Kootenay** | **Northwest** | **Northeast** | **CV (%)** |
| Population in 2020* | 3,065,668 | 502458 | 404076 | 401603 | 265752 | 263691 | 198734 | 195493 | 189822 | 152714 | 129122 | 88624 | 84414 | 56298 | 55606 | 42827 | 34434 |  |
| Total person-year in the population* | 31,039,615 | 4946661 | 4116484 | 4151170 | 2668466 | 2613411 | 2000915 | 1994890 | 1882828 | 1559185 | 1329379 | 896659 | 898808 | 586344 | 562941 | 460619 | 370855 |  |
| Total person-year at risk in the population | 28,925,909 | 4684930 | 3877426 | 3927398 | 2508063 | 2359997 | 1817126 | 1883568 | 1738095 | 1399543 | 1274055 | 819900 | 824730 | 531325 | 513555 | 427356 | 338842 |  |
| **Prevalence** | | | | | | | | | | | | | | | | | |  |
| Total person-year with diagnosed COPD | 2,113,706 | 261731 | 239058 | 223772 | 160403 | 253414 | 183789 | 111322 | 144733 | 159642 | 55324 | 76759 | 74078 | 55019 | 49386 | 33263 | 32013 |  |
| Period Prevalence (%) | 6.8 | 5.3 | 5.8 | 5.4 | 6.0 | 9.7 | 9.2 | 5.6 | 7.7 | 10.2 | 4.2 | 8.6 | 8.2 | 9.4 | 8.8 | 7.2 | 8.6 | 24.9 |
| Ranking of Period Prevalence^ |  | 15 | 12 | 14 | 11 | 2 | 8 | 13 | 9 | 1 | 16 | 5 | 7 | 3 | 4 | 10 | 6 |  |
| **Incidence** | | | | | | | | | | | | | | | | | |  |
| Number of new COPD Cases | 164,660 | 19198 | 18035 | 16419 | 12392 | 19020 | 14273 | 9279 | 11726 | 12798 | 4114 | 6383 | 6547 | 4287 | 4168 | 3009 | 3012 |  |
| Cumulative Incidence (per 100,000 person-year at risk) | 569.2 | 409.8 | 465.1 | 418.1 | 494.1 | 805.9 | 785.5 | 492.6 | 674.6 | 914.4 | 322.9 | 778.5 | 793.8 | 806.9 | 811.6 | 704.1 | 888.9 | 29.3 |
| Ranking of Cumulative Incidence^ |  | 15 | 13 | 14 | 11 | 5 | 7 | 12 | 10 | 1 | 16 | 8 | 6 | 4 | 3 | 9 | 2 |  |
| **Hospitalization** | | | | | | | | | | | | | | | | | |  |
| No of hospitalizations | 1,545,883 | 203308 | 173966 | 142433 | 109061 | 192174 | 118320 | 79673 | 117907 | 128479 | 34815 | 52805 | 60461 | 41228 | 40001 | 29803 | 21449 |  |
| Cumulative Hospitalization (per 100,000 diagnosed COPD-patient year) | 73136.1 | 77678.2 | 72771.5 | 63650.9 | 67991.9 | 75834.0 | 64378.2 | 71569.9 | 81465.2 | 80479.4 | 62929.3 | 68793.2 | 81618.0 | 74934.1 | 80996.6 | 89598.1 | 67000.9 | 10.5 |
| Ranking of Cumulative Hospitalization^ |  | 15 | 8 | 14 | 11 | 6 | 13 | 9 | 3 | 4 | 16 | 10 | 2 | 7 | 5 | 1 | 12 |  |
| **Mortality** | | | | | | | | | | | | | | | | | |  |
| Number of deaths (all-cause) | 106,474 | 13306 | 11252 | 10887 | 9069 | 14015 | 8743 | 5666 | 7271 | 8263 | 2340 | 3702 | 3593 | 2889 | 2637 | 1470 | 1371 |  |
| Cumulative Mortality (per 100,000 diagnosed COPD-patient year) | 5037.3 | 5083.8 | 4706.8 | 4865.2 | 5653.9 | 5530.5 | 4757.1 | 5089.7 | 5023.7 | 5176.0 | 4229.6 | 4822.9 | 4850.3 | 5250.9 | 5339.6 | 4419.3 | 4282.6 | 8.3 |
| Ranking of Cumulative Mortality^ |  | 7 | 13 | 10 | 1 | 2 | 12 | 6 | 8 | 4 | 16 | 11 | 9 | 5 | 3 | 15 | 14 |  |
| Abbreviations: HSDA: Health Service Delivery Area; CV: Coefficient of Variation  Count and Rates are based on aggregated data from 2010-2020  * Based on Statistics Canada population age 35 year or older  ^ Rankings are based on highest to lowest | | | | | | | | | | | | | | | | | | |

### **Table A2: Rate ratios and 95% confidence interval for the unadjusted, adjusted and trend adjusted models for Prevalence**

|  | **Unadjusted Model** | | | | | **Adjusted Model** | | | | **Trend Adjusted Model** | | | | |
| --- | --- | --- | --- | --- | --- | --- | --- | --- | --- | --- | --- | --- | --- | --- |
| **Term** | **p value** | **RR** | **95% CI (lower)** | **95% CI (upper)** | **p value** | | **RR** | **95% CI (lower)** | **95% CI (upper)** | | **p value** | **RR** | **95% CI (lower)** | **95% CI (upper)** |
| **(Intercept)** | 0.000 | 0.092 | 0.075 | 0.112 | 0.000 | | 0.012 | 0.010 | 0.014 | | 0.000 | 0.013 | 0.011 | 0.015 |
| **HSDA11** | 0.013 | 1.441 | 1.081 | 1.922 | 0.236 | | 0.923 | 0.809 | 1.054 | | 0.040 | 0.860 | 0.745 | 0.993 |
| **HSDA12** | 0.014 | 1.436 | 1.077 | 1.915 | 0.627 | | 0.970 | 0.859 | 1.096 | | 0.476 | 0.953 | 0.836 | 1.087 |
| **HSDA13** | 0.014 | 1.437 | 1.077 | 1.916 | 0.000 | | 1.194 | 1.115 | 1.278 | | 0.000 | 1.193 | 1.094 | 1.301 |
| **HSDA14** | 0.001 | 1.614 | 1.210 | 2.153 | 0.000 | | 1.231 | 1.121 | 1.351 | | 0.025 | 1.133 | 1.016 | 1.264 |
| **HSDA21** | 0.138 | 1.243 | 0.932 | 1.658 | 0.000 | | 1.165 | 1.107 | 1.227 | | 0.011 | 1.099 | 1.022 | 1.183 |
| **HSDA22** | 0.281 | 1.172 | 0.879 | 1.562 | 0.000 | | 1.152 | 1.106 | 1.199 | | 0.003 | 1.107 | 1.036 | 1.184 |
| **HSDA31** | 0.163 | 0.815 | 0.611 | 1.087 | 0.000 | | 0.746 | 0.712 | 0.781 | | 0.000 | 0.744 | 0.693 | 0.800 |
| **HSDA32** | 0.821 | 1.034 | 0.775 | 1.379 | 0.087 | | 0.953 | 0.902 | 1.007 | | 0.270 | 0.958 | 0.889 | 1.034 |
| **HSDA33** | 0.911 | 0.984 | 0.738 | 1.312 | 0.000 | | 0.736 | 0.673 | 0.805 | | 0.000 | 0.703 | 0.634 | 0.780 |
| **HSDA41** | 0.785 | 0.961 | 0.720 | 1.281 | 0.000 | | 0.879 | 0.841 | 0.919 | | 0.000 | 0.833 | 0.778 | 0.893 |
| **HSDA42** | 0.093 | 1.279 | 0.959 | 1.706 | 0.000 | | 1.203 | 1.129 | 1.281 | | 0.006 | 1.121 | 1.034 | 1.216 |
| **HSDA43** | 0.069 | 1.306 | 0.979 | 1.742 | 0.008 | | 1.094 | 1.024 | 1.168 | | 0.954 | 1.002 | 0.923 | 1.089 |
| **HSDA51** | 0.062 | 1.316 | 0.987 | 1.755 | 0.243 | | 0.943 | 0.854 | 1.041 | | 0.000 | 0.816 | 0.731 | 0.912 |
| **HSDA52** | 0.007 | 1.490 | 1.118 | 1.987 | 0.000 | | 1.262 | 1.183 | 1.347 | | 0.050 | 1.088 | 1.000 | 1.183 |
| **HSDA53** | 0.000 | 1.728 | 1.296 | 2.305 | 0.000 | | 1.283 | 1.146 | 1.435 | | 0.009 | 1.181 | 1.043 | 1.338 |
| **HSDA23 (Ref)** | | | | | | | | | | | | | | |
| **Female** |  |  |  |  | 0.000 | | 0.892 | 0.880 | 0.905 | | 0.000 | 0.891 | 0.880 | 0.903 |
| **Male (Ref)** | | | | | | | | | | | | | | |
| **50-64 years** |  |  |  |  | 0.000 | | 5.684 | 5.574 | 5.797 | | 0.000 | 5.698 | 5.590 | 5.807 |
| **65-79 years** |  |  |  |  | 0.000 | | 14.152 | 13.853 | 14.458 | | 0.000 | 14.203 | 13.912 | 14.501 |
| **>=80 years** |  |  |  |  | 0.000 | | 24.691 | 23.881 | 25.530 | | 0.000 | 24.693 | 23.902 | 25.510 |
| **35-49 years (Ref)** | | | | | | | | | | | | | | |
| **Rural** |  |  |  |  | 0.000 | | 2.157 | 1.723 | 2.700 | | 0.000 | 2.089 | 1.676 | 2.604 |
| **Neighborhood income quintile** | | | | | | | | | | | | | | |
| **2** |  |  |  |  | 0.000 | | 0.484 | 0.341 | 0.687 | | 0.000 | 0.493 | 0.349 | 0.695 |
| **3** |  |  |  |  | 0.002 | | 0.555 | 0.380 | 0.811 | | 0.010 | 0.613 | 0.423 | 0.889 |
| **4** |  |  |  |  | 0.003 | | 0.582 | 0.406 | 0.835 | | 0.006 | 0.609 | 0.427 | 0.870 |
| **5 (highest income quintile)** |  |  |  |  | 0.809 | | 1.040 | 0.755 | 1.433 | | 0.776 | 0.955 | 0.694 | 1.314 |
| **Unknown** |  |  |  |  | 0.361 | | 1.543 | 0.608 | 3.914 | | 0.217 | 1.775 | 0.714 | 4.413 |
| **Year** |  |  |  |  |  | |  |  |  | | 0.000 | 0.982 | 0.975 | 0.990 |
| **HSDA11:year** |  |  |  |  |  | |  |  |  | | 0.002 | 1.018 | 1.007 | 1.030 |
| **HSDA12:year** |  |  |  |  |  | |  |  |  | | 0.172 | 1.008 | 0.997 | 1.020 |
| **HSDA13:year** |  |  |  |  |  | |  |  |  | | 0.726 | 1.002 | 0.991 | 1.013 |
| **HSDA14:year** |  |  |  |  |  | |  |  |  | | 0.002 | 1.018 | 1.007 | 1.029 |
| **HSDA21:year** |  |  |  |  |  | |  |  |  | | 0.018 | 1.013 | 1.002 | 1.024 |
| **HSDA22:year** |  |  |  |  |  | |  |  |  | | 0.217 | 1.007 | 0.996 | 1.018 |
| **HSDA31:year** |  |  |  |  |  | |  |  |  | | 0.937 | 1.000 | 0.988 | 1.011 |
| **HSDA32:year** |  |  |  |  |  | |  |  |  | | 0.895 | 0.999 | 0.989 | 1.010 |
| **HSDA33:year** |  |  |  |  |  | |  |  |  | | 0.006 | 1.016 | 1.005 | 1.027 |
| **HSDA41:year** |  |  |  |  |  | |  |  |  | | 0.029 | 1.012 | 1.001 | 1.023 |
| **HSDA42:year** |  |  |  |  |  | |  |  |  | | 0.003 | 1.017 | 1.006 | 1.028 |
| **HSDA43:year** |  |  |  |  |  | |  |  |  | | 0.001 | 1.020 | 1.009 | 1.031 |
| **HSDA51:year** |  |  |  |  |  | |  |  |  | | 0.000 | 1.031 | 1.019 | 1.043 |
| **HSDA52:year** |  |  |  |  |  | |  |  |  | | 0.000 | 1.033 | 1.021 | 1.044 |
| **HSDA53:year** |  |  |  |  |  | |  |  |  | | 0.000 | 1.023 | 1.011 | 1.035 |
| **HSDA23: Year (Ref)** | | | | | | | | | | | | | | |
| Reference levels of factor variables have a RR=1.00.  Adjusted variables for both adjusted and trend-adjusted models: Sex, Socio-Economic Status, Area of Residence, Age group.  Abbreviations: RR: Rate Ratio; HSDA: Health Service Delivery Area, CI: Confidence Interval, HSDA11: East Kootenay, HSDA 12: Kootenay Boundary, HSDA 13: Okanagan, HSDA 14: Thompson Cariboo Shuswap, HSDA 21: Fraser East, HSDA 22: Fraser North, HSDA 23: Fraser South, HSDA 31: Richmond, HSDA 32: Vancouver, HSDA 33: North Shore/Coast Garibaldi, HSDA 41: South Vancouver Island, HSDA 42: Central Vancouver Island, HSDA 43: North Vancouver Island, HSDA 51: Northwest, HSDA 52: Northern Interior, HSDA 53: Northeast. | | | | | | | | | | | | | | |

### **Table A3: Rate ratios and 95% confidence interval for the unadjusted, adjusted and trend adjusted models for Incidence**

|  | **Unadjusted Model** | | | | **Adjusted Model** | | | | | **Trend Adjusted Model** | | | |
| --- | --- | --- | --- | --- | --- | --- | --- | --- | --- | --- | --- | --- | --- |
| **Term** | **p value** | **RR** | **95% CI (lower)** | **95% CI (upper)** | **p value** | **RR** | **95% CI (lower)** | **95% CI (upper)** | **p value** | | **RR** | **95% CI (lower)** | **95% CI (upper)** |
| **(Intercept)** | 0.000 | 0.006 | 0.005 | 0.007 | 0.000 | 0.002 | 0.001 | 0.002 | 0.000 | | 0.002 | 0.002 | 0.002 |
| **HSDA11** | 0.000 | 1.554 | 1.246 | 1.939 | 0.000 | 1.435 | 1.215 | 1.696 | 0.000 | | 1.317 | 1.136 | 1.527 |
| **HSDA12** | 0.001 | 1.479 | 1.185 | 1.845 | 0.000 | 1.366 | 1.158 | 1.611 | 0.000 | | 1.452 | 1.268 | 1.663 |
| **HSDA13** | 0.001 | 1.472 | 1.182 | 1.833 | 0.000 | 1.454 | 1.305 | 1.619 | 0.000 | | 1.505 | 1.359 | 1.668 |
| **HSDA14** | 0.000 | 1.733 | 1.391 | 2.159 | 0.000 | 1.647 | 1.441 | 1.883 | 0.000 | | 1.514 | 1.345 | 1.704 |
| **HSDA21** | 0.004 | 1.385 | 1.112 | 1.726 | 0.000 | 1.421 | 1.291 | 1.563 | 0.000 | | 1.498 | 1.360 | 1.650 |
| **HSDA22** | 0.158 | 1.171 | 0.941 | 1.459 | 0.000 | 1.171 | 1.073 | 1.278 | 0.000 | | 1.218 | 1.111 | 1.335 |
| **HSDA31** | 0.092 | 0.827 | 0.663 | 1.031 | 0.000 | 0.765 | 0.695 | 0.843 | 0.000 | | 0.730 | 0.656 | 0.812 |
| **HSDA32** | 0.976 | 0.997 | 0.800 | 1.241 | 0.818 | 0.989 | 0.900 | 1.087 | 0.877 | | 0.993 | 0.903 | 1.091 |
| **HSDA33** | 0.288 | 1.127 | 0.904 | 1.403 | 0.997 | 1.000 | 0.885 | 1.130 | 0.045 | | 0.891 | 0.797 | 0.997 |
| **HSDA41** | 0.866 | 1.019 | 0.818 | 1.269 | 0.907 | 1.005 | 0.919 | 1.100 | 0.214 | | 0.941 | 0.856 | 1.035 |
| **HSDA42** | 0.004 | 1.375 | 1.104 | 1.713 | 0.000 | 1.466 | 1.321 | 1.627 | 0.000 | | 1.414 | 1.280 | 1.563 |
| **HSDA43** | 0.003 | 1.399 | 1.122 | 1.744 | 0.000 | 1.421 | 1.272 | 1.588 | 0.000 | | 1.299 | 1.165 | 1.449 |
| **HSDA51** | 0.013 | 1.328 | 1.062 | 1.660 | 0.000 | 1.363 | 1.179 | 1.575 | 0.401 | | 1.059 | 0.927 | 1.209 |
| **HSDA52** | 0.000 | 1.540 | 1.235 | 1.920 | 0.000 | 1.684 | 1.513 | 1.875 | 0.033 | | 1.125 | 1.009 | 1.253 |
| **HSDA53** | 0.000 | 1.860 | 1.488 | 2.325 | 0.000 | 1.892 | 1.633 | 2.192 | 0.000 | | 1.605 | 1.406 | 1.832 |
| **HSDA23 (Ref)** | | | | | | | | | | | | | |
| **Female** |  |  |  |  | 0.000 | 0.850 | 0.822 | 0.878 | 0.000 | | 0.841 | 0.824 | 0.858 |
| **Male (Ref)** | | | | | | | | | | | | | |
| **50-64 years** |  |  |  |  | 0.000 | 3.195 | 3.048 | 3.348 | 0.000 | | 3.207 | 3.112 | 3.304 |
| **65-79 years** |  |  |  |  | 0.000 | 5.884 | 5.606 | 6.177 | 0.000 | | 5.839 | 5.660 | 6.023 |
| **>=80 years** |  |  |  |  | 0.000 | 8.017 | 7.621 | 8.434 | 0.000 | | 8.101 | 7.836 | 8.375 |
| **35-49 years (Ref)** | | | | | | | | | | | | | |
| **Rural** |  |  |  |  | 0.024 | 1.336 | 1.039 | 1.718 | 0.000 | | 1.481 | 1.239 | 1.769 |
| **Neighborhood income quintile** | | | | | | | | | | | | | |
| **2** |  |  |  |  | 0.008 | 0.623 | 0.439 | 0.884 | 0.113 | | 0.824 | 0.649 | 1.047 |
| **3** |  |  |  |  | 0.030 | 0.666 | 0.461 | 0.962 | 0.402 | | 1.112 | 0.867 | 1.426 |
| **4** |  |  |  |  | 0.642 | 0.918 | 0.642 | 1.314 | 0.008 | | 1.390 | 1.091 | 1.772 |
| **5 (highest income quintile)** |  |  |  |  | 0.660 | 0.925 | 0.653 | 1.309 | 0.036 | | 1.286 | 1.017 | 1.626 |
| **Unknown** |  |  |  |  | 0.578 | 1.288 | 0.528 | 3.145 | 0.051 | | 1.821 | 0.998 | 3.323 |
| **Year** |  |  |  |  |  |  |  |  | 0.000 | | 0.922 | 0.912 | 0.932 |
| **HSDA11:year** |  |  |  |  |  |  |  |  | 0.480 | | 1.007 | 0.988 | 1.025 |
| **HSDA12:year** |  |  |  |  |  |  |  |  | 0.081 | | 0.984 | 0.966 | 1.002 |
| **HSDA13:year** |  |  |  |  |  |  |  |  | 0.410 | | 0.993 | 0.978 | 1.009 |
| **HSDA14:year** |  |  |  |  |  |  |  |  | 0.103 | | 1.014 | 0.997 | 1.030 |
| **HSDA21:year** |  |  |  |  |  |  |  |  | 0.400 | | 0.993 | 0.977 | 1.009 |
| **HSDA22:year** |  |  |  |  |  |  |  |  | 0.205 | | 0.990 | 0.975 | 1.006 |
| **HSDA31:year** |  |  |  |  |  |  |  |  | 0.048 | | 1.018 | 1.000 | 1.037 |
| **HSDA32:year** |  |  |  |  |  |  |  |  | 0.243 | | 1.009 | 0.994 | 1.025 |
| **HSDA33:year** |  |  |  |  |  |  |  |  | 0.052 | | 1.016 | 1.000 | 1.033 |
| **HSDA41:year** |  |  |  |  |  |  |  |  | 0.010 | | 1.021 | 1.005 | 1.038 |
| **HSDA42:year** |  |  |  |  |  |  |  |  | 0.647 | | 1.004 | 0.988 | 1.020 |
| **HSDA43:year** |  |  |  |  |  |  |  |  | 0.009 | | 1.023 | 1.006 | 1.041 |
| **HSDA51:year** |  |  |  |  |  |  |  |  | 0.000 | | 1.055 | 1.035 | 1.076 |
| **HSDA52:year** |  |  |  |  |  |  |  |  | 0.000 | | 1.088 | 1.069 | 1.107 |
| **HSDA53:year** |  |  |  |  |  |  |  |  | 0.009 | | 1.026 | 1.007 | 1.046 |
| **HSDA23: Year (Ref)** | | | | | | | | | | | | | |
| Reference levels of factor variables have a RR=1.00.  Adjusted variables for both adjusted and trend-adjusted models: Sex, Socio-Economic Status, Area of Residence, Age group.  Abbreviations: RR: Rate Ratio; HSDA: Health Service Delivery Area, CI: Confidence Interval, HSDA11: East Kootenay, HSDA 12: Kootenay Boundary, HSDA 13: Okanagan, HSDA 14: Thompson Cariboo Shuswap, HSDA 21: Fraser East, HSDA 22: Fraser North, HSDA 23: Fraser South, HSDA 31: Richmond, HSDA 32: Vancouver, HSDA 33: North Shore/Coast Garibaldi, HSDA 41: South Vancouver Island, HSDA 42: Central Vancouver Island, HSDA 43: North Vancouver Island, HSDA 51: Northwest, HSDA 52: Northern Interior, HSDA 53: Northeast. | | | | | | | | | | | | | |

### **Table A4: Rate ratios and 95% confidence interval for the unadjusted, adjusted and trend adjusted models for all-cause hospitalization**

|  | | **Unadjusted Model** | | | | **Adjusted Model** | | | | **Trend Adjusted Model** | | | |
| --- | --- | --- | --- | --- | --- | --- | --- | --- | --- | --- | --- | --- | --- |
| **Term** | **p value** | | **RR** | **95% CI (lower)** | **95% CI (upper)** | **p value** | **RR** | **95% CI (lower)** | **95% CI (upper)** | **p value** | **RR** | **95% CI (lower)** | **95% CI (upper)** |
| **(Intercept)** | | 0.000 | 0.701 | 0.659 | 0.747 | 0.002 | 0.569 | 0.399 | 0.813 | 0.063 | 0.760 | 0.569 | 1.015 |
| **HSDA11** | | 0.084 | 1.082 | 0.989 | 1.184 | 0.010 | 1.110 | 1.025 | 1.201 | 0.028 | 1.131 | 1.013 | 1.263 |
| **HSDA12** | | 0.015 | 1.118 | 1.022 | 1.223 | 0.007 | 1.133 | 1.035 | 1.239 | 0.342 | 1.057 | 0.942 | 1.186 |
| **HSDA13** | | 0.094 | 1.079 | 0.987 | 1.179 | 0.001 | 1.162 | 1.064 | 1.269 | 0.044 | 1.122 | 1.003 | 1.255 |
| **HSDA14** | | 0.113 | 1.075 | 0.983 | 1.175 | 0.000 | 1.202 | 1.096 | 1.319 | 0.002 | 1.199 | 1.069 | 1.344 |
| **HSDA21** | | 0.628 | 1.022 | 0.935 | 1.117 | 0.174 | 1.066 | 0.972 | 1.168 | 0.035 | 1.132 | 1.009 | 1.269 |
| **HSDA22** | | 0.593 | 0.976 | 0.893 | 1.067 | 0.602 | 0.979 | 0.903 | 1.061 | 0.830 | 0.988 | 0.886 | 1.102 |
| **HSDA31** | | 0.663 | 1.020 | 0.933 | 1.116 | 0.211 | 1.060 | 0.967 | 1.162 | 0.711 | 1.022 | 0.910 | 1.149 |
| **HSDA32** | | 0.507 | 0.970 | 0.888 | 1.061 | 0.215 | 1.065 | 0.964 | 1.177 | 0.665 | 0.974 | 0.865 | 1.097 |
| **HSDA33** | | 0.308 | 1.048 | 0.958 | 1.145 | 0.605 | 1.032 | 0.916 | 1.163 | 0.672 | 1.028 | 0.905 | 1.168 |
| **HSDA41** | | 0.166 | 1.065 | 0.974 | 1.164 | 0.007 | 1.124 | 1.032 | 1.224 | 0.179 | 1.080 | 0.965 | 1.208 |
| **HSDA42** | | 0.345 | 1.044 | 0.955 | 1.141 | 0.168 | 1.060 | 0.976 | 1.152 | 0.038 | 1.123 | 1.006 | 1.253 |
| **HSDA43** | | 0.046 | 1.096 | 1.002 | 1.198 | 0.003 | 1.143 | 1.047 | 1.247 | 0.003 | 1.186 | 1.059 | 1.327 |
| **HSDA51** | | 0.015 | 1.119 | 1.022 | 1.224 | 0.000 | 1.217 | 1.103 | 1.343 | 0.000 | 1.273 | 1.133 | 1.431 |
| **HSDA52** | | 0.103 | 1.077 | 0.985 | 1.178 | 0.190 | 1.057 | 0.973 | 1.149 | 0.043 | 1.122 | 1.003 | 1.254 |
| **HSDA53** | | 0.168 | 1.065 | 0.974 | 1.165 | 0.676 | 1.026 | 0.909 | 1.158 | 0.934 | 0.994 | 0.872 | 1.134 |
| **HSDA23 (Ref)** | |  |  |  |  |  |  |  |  |  |  |  |  |
| **Female** | |  |  |  |  | 0.091 | 0.976 | 0.949 | 1.004 | 0.035 | 0.977 | 0.957 | 0.998 |
| **Male (Ref)** | |  |  |  |  |  |  |  |  |  |  |  |  |
| **50-64 years** | |  |  |  |  | 0.000 | 0.915 | 0.878 | 0.953 | 0.000 | 0.943 | 0.914 | 0.973 |
| **65-79 years** | |  |  |  |  | 0.000 | 1.196 | 1.148 | 1.246 | 0.000 | 1.237 | 1.199 | 1.277 |
| **>=80 years** | |  |  |  |  | 0.000 | 1.351 | 1.296 | 1.407 | 0.000 | 1.368 | 1.326 | 1.412 |
| **35-49 years (Ref)** | |  |  |  |  |  |  |  |  |  |  |  |  |
| **Rural** | |  |  |  |  | 0.241 | 1.328 | 0.826 | 2.135 | 0.388 | 0.847 | 0.582 | 1.234 |
| **Neighborhood income quintile** | |  |  |  |  |  |  |  |  |  |  |  |  |
| **2** | |  |  |  |  | 0.778 | 0.924 | 0.533 | 1.602 | 0.482 | 1.169 | 0.757 | 1.805 |
| **3** | |  |  |  |  | 0.855 | 1.055 | 0.595 | 1.869 | 0.721 | 0.921 | 0.585 | 1.449 |
| **4** | |  |  |  |  | 0.329 | 1.324 | 0.754 | 2.326 | 0.948 | 1.015 | 0.649 | 1.588 |
| **5 (highest income quintile)** | |  |  |  |  | 0.272 | 1.353 | 0.789 | 2.321 | 0.062 | 1.503 | 0.980 | 2.305 |
| **Unknown** | |  |  |  |  | 0.001 | 0.107 | 0.030 | 0.378 | 0.001 | 0.190 | 0.070 | 0.514 |
| **Year** | |  |  |  |  |  |  |  |  | 0.000 | 0.952 | 0.940 | 0.964 |
| **HSDA11:year** | |  |  |  |  |  |  |  |  | 0.487 | 0.993 | 0.975 | 1.012 |
| **HSDA12:year** | |  |  |  |  |  |  |  |  | 0.243 | 1.011 | 0.993 | 1.030 |
| **HSDA13:year** | |  |  |  |  |  |  |  |  | 0.753 | 1.003 | 0.985 | 1.021 |
| **HSDA14:year** | |  |  |  |  |  |  |  |  | 0.494 | 0.994 | 0.976 | 1.012 |
| **HSDA21:year** | |  |  |  |  |  |  |  |  | 0.060 | 0.983 | 0.965 | 1.001 |
| **HSDA22:year** | |  |  |  |  |  |  |  |  | 0.888 | 1.001 | 0.983 | 1.020 |
| **HSDA31:year** | |  |  |  |  |  |  |  |  | 0.336 | 1.009 | 0.991 | 1.028 |
| **HSDA32:year** | |  |  |  |  |  |  |  |  | 0.239 | 1.011 | 0.993 | 1.029 |
| **HSDA33:year** | |  |  |  |  |  |  |  |  | 0.517 | 0.994 | 0.976 | 1.012 |
| **HSDA41:year** | |  |  |  |  |  |  |  |  | 0.643 | 1.004 | 0.986 | 1.023 |
| **HSDA42:year** | |  |  |  |  |  |  |  |  | 0.103 | 0.985 | 0.967 | 1.003 |
| **HSDA43:year** | |  |  |  |  |  |  |  |  | 0.137 | 0.986 | 0.968 | 1.004 |
| **HSDA51:year** | |  |  |  |  |  |  |  |  | 0.143 | 0.986 | 0.967 | 1.005 |
| **HSDA52:year** | |  |  |  |  |  |  |  |  | 0.173 | 0.987 | 0.969 | 1.006 |
| **HSDA53:year** | |  |  |  |  |  |  |  |  | 0.740 | 1.003 | 0.985 | 1.022 |
| **HSDA23: Year (Ref)** | | | | | | | | | | | | | |
| Reference levels of factor variables have a RR=1.00.  Adjusted variables for both adjusted and trend-adjusted models: Sex, Socio-Economic Status, Area of Residence, Age group.  Abbreviations: RR: Rate Ratio; HSDA: Health Service Delivery Area, CI: Confidence Interval, HSDA11: East Kootenay, HSDA 12: Kootenay Boundary, HSDA 13: Okanagan, HSDA 14: Thompson Cariboo Shuswap, HSDA 21: Fraser East, HSDA 22: Fraser North, HSDA 23: Fraser South, HSDA 31: Richmond, HSDA 32: Vancouver, HSDA 33: North Shore/Coast Garibaldi, HSDA 41: South Vancouver Island, HSDA 42: Central Vancouver Island, HSDA 43: North Vancouver Island, HSDA 51: Northwest, HSDA 52: Northern Interior, HSDA 53: Northeast. | | | | | | | | | | | | | |

### **Table A5: Rate ratios and 95% confidence interval for the unadjusted, adjusted and trend adjusted models for All-cause Mortality**

|  | **Unadjusted Model** | | | | **Adjusted Model** | | | | **Trend Adjusted Model** | | | |
| --- | --- | --- | --- | --- | --- | --- | --- | --- | --- | --- | --- | --- |
| **Term** | **p value** | **RR** | **95% CI (lower)** | **95% CI (upper)** | **p value** | **RR** | **95% CI (lower)** | **95% CI (upper)** | **p value** | **RR** | **95% CI (lower)** | **95% CI (upper)** |
| **(Intercept)** | 0.000 | 0.046 | 0.037 | 0.056 | 0.000 | 0.008 | 0.007 | 0.008 | 0.000 | 0.007 | 0.007 | 0.008 |
| **HSDA11** | 0.560 | 1.092 | 0.813 | 1.466 | 0.057 | 1.095 | 0.997 | 1.203 | 0.095 | 1.112 | 0.982 | 1.259 |
| **HSDA12** | 0.460 | 1.117 | 0.832 | 1.500 | 0.015 | 1.114 | 1.021 | 1.215 | 0.063 | 1.121 | 0.994 | 1.264 |
| **HSDA13** | 0.879 | 1.023 | 0.765 | 1.367 | 0.210 | 1.034 | 0.981 | 1.090 | 0.400 | 1.035 | 0.955 | 1.122 |
| **HSDA14** | 0.705 | 1.058 | 0.791 | 1.415 | 0.036 | 1.073 | 1.005 | 1.145 | 0.311 | 1.050 | 0.956 | 1.154 |
| **HSDA21** | 0.942 | 1.011 | 0.756 | 1.352 | 0.071 | 1.044 | 0.996 | 1.095 | 0.981 | 1.001 | 0.922 | 1.086 |
| **HSDA22** | 0.602 | 0.926 | 0.693 | 1.237 | 0.000 | 0.912 | 0.878 | 0.948 | 0.008 | 0.906 | 0.842 | 0.974 |
| **HSDA31** | 0.216 | 0.830 | 0.618 | 1.115 | 0.000 | 0.795 | 0.754 | 0.839 | 0.215 | **0.939** | 0.849 | 1.038 |
| **HSDA32** | 0.625 | 0.930 | 0.696 | 1.243 | 0.013 | 0.945 | 0.905 | 0.988 | 0.020 | 0.912 | 0.844 | 0.985 |
| **HSDA33** | 0.654 | 0.935 | 0.699 | 1.253 | 0.019 | 0.930 | 0.876 | 0.988 | 0.341 | 0.955 | 0.870 | 1.050 |
| **HSDA41** | 0.811 | 1.036 | 0.775 | 1.386 | 0.463 | 1.016 | 0.974 | 1.060 | 0.730 | 0.987 | 0.914 | 1.065 |
| **HSDA42** | 0.835 | 0.970 | 0.725 | 1.297 | 0.257 | 0.973 | 0.928 | 1.020 | 0.371 | 0.964 | 0.890 | 1.044 |
| **HSDA43** | 0.955 | 0.992 | 0.740 | 1.330 | 0.744 | 1.010 | 0.953 | 1.070 | 0.625 | 1.024 | 0.931 | 1.127 |
| **HSDA51** | 0.944 | 1.011 | 0.751 | 1.360 | 0.363 | 1.041 | 0.955 | 1.135 | 0.069 | 1.133 | 0.991 | 1.296 |
| **HSDA52** | 0.560 | 1.091 | 0.814 | 1.461 | 0.000 | 1.132 | 1.070 | 1.198 | 0.039 | 1.107 | 1.005 | 1.219 |
| **HSDA53** | 0.701 | 1.060 | 0.789 | 1.423 | 0.032 | 1.100 | 1.008 | 1.200 | 0.002 | 1.237 | 1.083 | 1.413 |
| **HSDA23 (Ref)** | | | | | | | | | | | | |
| **Female** |  |  |  |  | 0.000 | 0.807 | 0.794 | 0.821 | 0.000 | 0.807 | 0.794 | 0.821 |
| **Male (Ref)** | | | | | | | | | | | | |
| **50-64 years** |  |  |  |  | 0.000 | 2.240 | 2.066 | 2.429 | 0.000 | 2.231 | 2.058 | 2.419 |
| **65-79 years** |  |  |  |  | 0.000 | 5.400 | 4.986 | 5.849 | 0.000 | 5.377 | 4.964 | 5.824 |
| **>=80 years** |  |  |  |  | 0.000 | 16.793 | 15.508 | 18.184 | 0.000 | 16.721 | 15.442 | 18.106 |
| **35-49 years (Ref)** | | | | | | | | | | | | |
| **Rural** |  |  |  |  | 0.434 | 0.944 | 0.818 | 1.090 | 0.460 | 0.947 | 0.820 | 1.094 |
| **Neighborhood income quintile** | | | | | | | | | | | | |
| **2** |  |  |  |  | 0.919 | 1.008 | 0.857 | 1.186 | 0.574 | 1.048 | 0.890 | 1.235 |
| **3** |  |  |  |  | 0.042 | 1.213 | 1.007 | 1.461 | 0.012 | 1.271 | 1.054 | 1.533 |
| **4** |  |  |  |  | 0.035 | 1.235 | 1.015 | 1.502 | 0.071 | 1.201 | 0.985 | 1.464 |
| **5 (highest income quintile)** |  |  |  |  | 0.926 | 0.991 | 0.811 | 1.210 | 0.830 | 1.022 | 0.836 | 1.250 |
| **Unknown** |  |  |  |  | 0.337 | 1.262 | 0.785 | 2.027 | 0.185 | 1.377 | 0.858 | 2.211 |
| **Year** |  |  |  |  |  |  |  |  | 0.610 | 1.002 | 0.994 | 1.010 |
| **HSDA11:year** |  |  |  |  |  |  |  |  | 0.656 | 0.996 | 0.980 | 1.013 |
| **HSDA12:year** |  |  |  |  |  |  |  |  | 0.840 | 0.998 | 0.983 | 1.014 |
| **HSDA13:year** |  |  |  |  |  |  |  |  | 0.976 | 1.000 | 0.988 | 1.012 |
| **HSDA14:year** |  |  |  |  |  |  |  |  | 0.578 | 1.004 | 0.991 | 1.016 |
| **HSDA21:year** |  |  |  |  |  |  |  |  | 0.206 | 1.008 | 0.995 | 1.021 |
| **HSDA22:year** |  |  |  |  |  |  |  |  | 0.774 | 1.002 | 0.990 | 1.014 |
| **HSDA31:year** |  |  |  |  |  |  |  |  | 0.000 | 0.968 | 0.952 | 0.984 |
| **HSDA32:year** |  |  |  |  |  |  |  |  | 0.256 | 1.007 | 0.995 | 1.019 |
| **HSDA33:year** |  |  |  |  |  |  |  |  | 0.439 | 0.995 | 0.981 | 1.008 |
| **HSDA41:year** |  |  |  |  |  |  |  |  | 0.355 | 1.006 | 0.993 | 1.019 |
| **HSDA42:year** |  |  |  |  |  |  |  |  | 0.776 | 1.002 | 0.989 | 1.014 |
| **HSDA43:year** |  |  |  |  |  |  |  |  | 0.706 | 0.997 | 0.983 | 1.012 |
| **HSDA51:year** |  |  |  |  |  |  |  |  | 0.095 | 0.984 | 0.965 | 1.003 |
| **HSDA52:year** |  |  |  |  |  |  |  |  | 0.538 | 1.005 | 0.990 | 1.020 |
| **HSDA53:year** |  |  |  |  |  |  |  |  | 0.020 | 0.98 | 0.958 | 0.996 |
| **HSDA23:Year (Ref)** | | | | | | | | | | | | |
| Reference levels of factor variables have a RR=1.00.  Adjusted variables for both adjusted and trend-adjusted models: Sex, Socio-Economic Status, Area of Residence, Age group.  Abbreviations: RR: Rate Ratio; HSDA: Health Service Delivery Area, CI: Confidence Interval, HSDA11: East Kootenay, HSDA 12: Kootenay Boundary, HSDA 13: Okanagan, HSDA 14: Thompson Cariboo Shuswap, HSDA 21: Fraser East, HSDA 22: Fraser North, HSDA 23: Fraser South, HSDA 31: Richmond, HSDA 32: Vancouver, HSDA 33: North Shore/Coast Garibaldi, HSDA 41: South Vancouver Island, HSDA 42: Central Vancouver Island, HSDA 43: North Vancouver Island, HSDA 51: Northwest, HSDA 52: Northern Interior, HSDA 53: Northeast. | | | | | | | | | | | | |

### **Table A6: Number of COPD cases and Sex and age Standardized Prevalence (per 100,000 persons) in British Columbia, Canada, from 2001 to 2020, stratified by Health Service Delivery Area**

| **Year** | **2010** | **2011** | | **2012** | | **2013** | | **2014** | | **2015** | | **2016** | | **2017** | | **2018** | | **2019** | | **2020** | | **2010** | **2011** | **2012** | **2013** | **2014** | **2015** | **2016** | **2017** | **2018** | **2019** | **2020** |
| --- | --- | --- | --- | --- | --- | --- | --- | --- | --- | --- | --- | --- | --- | --- | --- | --- | --- | --- | --- | --- | --- | --- | --- | --- | --- | --- | --- | --- | --- | --- | --- | --- |
| **Number of COPD cases** | | | | | | | | | | | | | | | | | | | | | | **Sex and age Standardized Prevalence (per 100,000 persons)** | | | | | | | | | | |
| **Total** | 161661 | | 171768 | | 179379 | | 185680 | | 191257 | | 196587 | | 200237 | | 204300 | | 207014 | | 209150 | | 209285 | 6882.8 | 7121.2 | 7213.3 | 7231.4 | 7215.4 | 7218.1 | 7173.1 | 7114.1 | 7005.0 | 6871.6 | 6682.0 |
| **HSDA11** | 3728 | | 4008 | | 4158 | | 4308 | | 4503 | | 4673 | | 4727 | | 4778 | | 4780 | | 4837 | | 4886 | 8199.0 | 8672.1 | 8744.4 | 8779.3 | 8883.1 | 8962.4 | 8829.0 | 8676.2 | 8420.4 | 8295.6 | 8167.7 |
| **HSDA12** | 4291 | | 4638 | | 4834 | | 4981 | | 5052 | | 5106 | | 5144 | | 5225 | | 5273 | | 5287 | | 5188 | 8550.0 | 9063.3 | 9198.1 | 9235.2 | 9134.8 | 9033.8 | 8903.1 | 8820.6 | 8681.6 | 8547.0 | 8197.4 |
| **HSDA13** | 19950 | | 21001 | | 21948 | | 22604 | | 22881 | | 23328 | | 23762 | | 24198 | | 24474 | | 24601 | | 24667 | 8551.6 | 8798.3 | 8967.7 | 8983.6 | 8804.6 | 8721.7 | 8641.6 | 8557.4 | 8413.5 | 8227.8 | 8018.1 |
| **HSDA14** | 12074 | | 12924 | | 13473 | | 14010 | | 14366 | | 14596 | | 14964 | | 15378 | | 15845 | | 16044 | | 15968 | 9338.5 | 9791.1 | 9929.0 | 10035.6 | 9990.0 | 9903.5 | 9893.1 | 9887.8 | 9898.7 | 9739.0 | 9454.9 |
| **HSDA21** | 10199 | | 10980 | | 11920 | | 12534 | | 12993 | | 13497 | | 13892 | | 14483 | | 14626 | | 14773 | | 14836 | 7188.9 | 7521.9 | 7886.6 | 8032.2 | 8031.6 | 8054.6 | 8028.4 | 8093.5 | 7928.1 | 7775.8 | 7593.8 |
| **HSDA22** | 18342 | | 19688 | | 20519 | | 21176 | | 21908 | | 22511 | | 22887 | | 23161 | | 22994 | | 23004 | | 22868 | 6802.1 | 7060.7 | 7107.2 | 7086.5 | 7066.7 | 7060.8 | 6998.6 | 6891.6 | 6659.8 | 6484.5 | 6270.8 |
| **HSDA23** | 20167 | | 21237 | | 22024 | | 22774 | | 23635 | | 24519 | | 25037 | | 25450 | | 25678 | | 25699 | | 25511 | 6050.5 | 6134.3 | 6118.1 | 6069.3 | 6068.4 | 6107.5 | 6050.4 | 5937.1 | 5792.9 | 5590.4 | 5351.4 |
| **HSDA31** | 4520 | | 4690 | | 4792 | | 4886 | | 5028 | | 5174 | | 5201 | | 5213 | | 5232 | | 5297 | | 5291 | 4785.7 | 4854.5 | 4833.3 | 4721.6 | 4698.3 | 4689.9 | 4598.6 | 4514.4 | 4422.6 | 4353.7 | 4246.5 |
| **HSDA32** | 18170 | | 19033 | | 19747 | | 20185 | | 20746 | | 21277 | | 21155 | | 21059 | | 20799 | | 20775 | | 20826 | 6141.3 | 6291.0 | 6360.9 | 6302.0 | 6320.0 | 6369.0 | 6257.1 | 6093.3 | 5874.4 | 5684.6 | 5556.5 |
| **HSDA33** | 8190 | | 8800 | | 9322 | | 9681 | | 10218 | | 10682 | | 10732 | | 10824 | | 10897 | | 10987 | | 10989 | 5232.4 | 5481.0 | 5647.7 | 5697.2 | 5836.6 | 5941.4 | 5857.0 | 5765.3 | 5660.9 | 5562.6 | 5411.1 |
| **HSDA41** | 12619 | | 13209 | | 13591 | | 13964 | | 14261 | | 14540 | | 14927 | | 15419 | | 15647 | | 16170 | | 16056 | 5509.3 | 5685.3 | 5709.1 | 5712.6 | 5669.0 | 5635.9 | 5646.6 | 5695.7 | 5636.6 | 5661.5 | 5466.5 |
| **HSDA42** | 13421 | | 14371 | | 15132 | | 15837 | | 16262 | | 16672 | | 17336 | | 18107 | | 18741 | | 18894 | | 19016 | 7578.2 | 7925.0 | 8104.4 | 8257.2 | 8220.5 | 8196.7 | 8283.1 | 8389.6 | 8419.1 | 8240.3 | 8057.2 |
| **HSDA43** | 5445 | | 5918 | | 6209 | | 6484 | | 6722 | | 6957 | | 7228 | | 7583 | | 7913 | | 8151 | | 8149 | 7420.8 | 7786.6 | 7890.6 | 7998.2 | 8006.1 | 8030.6 | 8084.8 | 8144.5 | 8210.4 | 8168.6 | 7912.2 |
| **HSDA51** | 2444 | | 2577 | | 2667 | | 2840 | | 3014 | | 3145 | | 3197 | | 3248 | | 3307 | | 3358 | | 3466 | 7666.4 | 7751.4 | 7801.9 | 8082.6 | 8351.8 | 8538.2 | 8510.3 | 8387.8 | 8347.7 | 8288.5 | 8309.4 |
| **HSDA52** | 5442 | | 5828 | | 6065 | | 6324 | | 6660 | | 6839 | | 6952 | | 7004 | | 7314 | | 7663 | | 7987 | 8673.1 | 8974.4 | 9062.0 | 9183.8 | 9407.7 | 9456.3 | 9405.8 | 9219.5 | 9344.9 | 9532.5 | 9721.7 |
| **HSDA53** | 2401 | | 2600 | | 2711 | | 2778 | | 2937 | | 3027 | | 3035 | | 3038 | | 3050 | | 3194 | | 3242 | 10272.2 | 10779.3 | 10877.2 | 10838.7 | 11231.6 | 11522.3 | 11485.5 | 11267.2 | 11073.0 | 11221.9 | 11063.2 |

Abbreviations: COPD: Chronic Obstructive Pulmonary Disease, HSDA: Health Services Delivery Area; HSDA11: East Kootenay, HSDA 12: Kootenay Boundary, HSDA 13: Okanagan, HSDA 14: Thompson Cariboo Shuswap, HSDA 21: Fraser East, HSDA 22: Fraser North, HSDA 23: Fraser South, HSDA 31: Richmond, HSDA 32: Vancouver, HSDA 33: North Shore/Coast Garibaldi, HSDA 41: South Vancouver Island, HSDA 42: Central Vancouver Island, HSDA 43: North Vancouver Island, HSDA 51: Northwest, HSDA 52: Northern Interior, HSDA 53: Northeast.

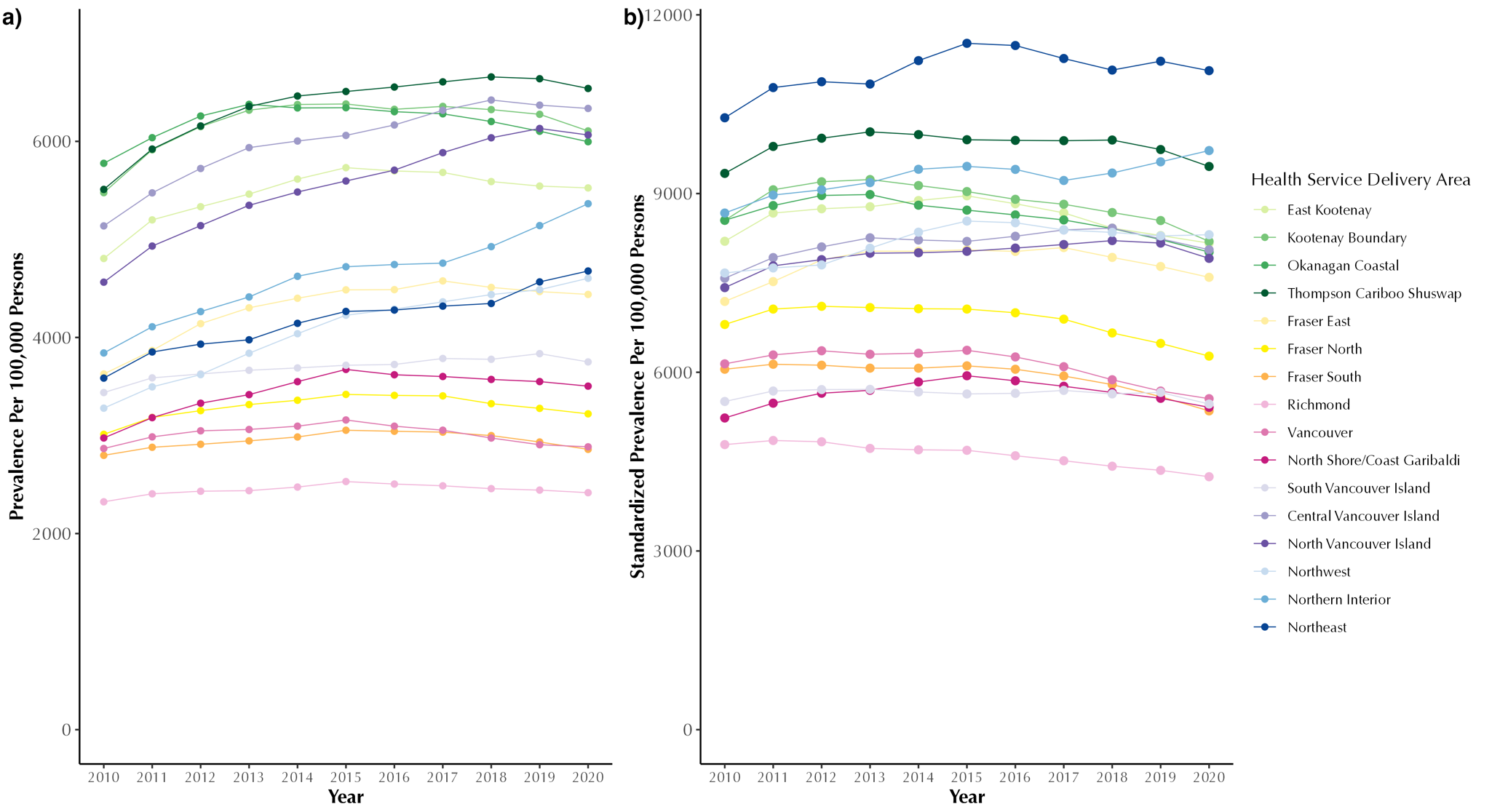

### **Figure A2: Annual crude (a) and sex and age standardized (b) chronic obstructive pulmonary disease prevalence (per 100,000 persons) in British Columbia, Canada, from 2001 to 2020, stratified by Health Service Delivery Area**

### **Table A7: Number of new COPD cases and Sex and age Standardized Incidence (per 100,000) in British Columbia, Canada, from 2001 to 2020, stratified by Health Service Delivery Area**

| **Year of Diagnosis** | | | | | | | | | | | | | | | | | | | | | | |
| --- | --- | --- | --- | --- | --- | --- | --- | --- | --- | --- | --- | --- | --- | --- | --- | --- | --- | --- | --- | --- | --- | --- |
|  | **2010** | **2011** | **2012** | **2013** | **2014** | **2015** | **2016** | **2017** | **2018** | **2019** | **2020** | **2010** | **2011** | **2012** | **2013** | **2014** | **2015** | **2016** | **2017** | **2018** | **2019** | **2020** |
|  | **Number of new COPD cases** | | | | | | | | | | | **Sex and age Standardized Incidence Rate (per 100,000 persons at risk)*** | | | | | | | | | | |
| **Total** | 20229 | 18058 | 16321 | 15360 | 14994 | 14721 | 13680 | 13909 | 13684 | 13060 | 10807 | 570.9 | 499.8 | 440.9 | 403.9 | 383.5 | 368 | 334.2 | 331.1 | 317.2 | 293.5 | 236.5 |
| **HSDA11** | 512 | 472 | 387 | 391 | 407 | 413 | 358 | 326 | 316 | 314 | 272 | 770.8 | 711.6 | 564.3 | 554.5 | 565.9 | 562.6 | 467.3 | 414.8 | 393.1 | 380.7 | 321.4 |
| **HSDA12** | 575 | 577 | 434 | 411 | 359 | 359 | 340 | 339 | 366 | 320 | 207 | 783.5 | 777.6 | 582.8 | 541.9 | 460.6 | 448.8 | 416.5 | 414.7 | 428.5 | 377.4 | 239.3 |
| **HSDA13** | 2535 | 2130 | 2011 | 1809 | 1659 | 1738 | 1543 | 1456 | 1449 | 1440 | 1250 | 771.5 | 644.1 | 595.1 | 523.1 | 466.9 | 475.3 | 411.5 | 377 | 365 | 356 | 298.6 |
| **HSDA14** | 1544 | 1413 | 1207 | 1249 | 1198 | 1081 | 1022 | 1204 | 1133 | 1018 | 729 | 825 | 755 | 636.6 | 645.6 | 602.5 | 533.8 | 492.7 | 570.4 | 517.8 | 455.3 | 315.6 |
| **HSDA21** | 1552 | 1232 | 1366 | 1119 | 988 | 970 | 926 | 987 | 870 | 878 | 838 | 724.6 | 560.3 | 608.2 | 485 | 416.4 | 397.9 | 371.3 | 383.7 | 329.1 | 320.8 | 298.1 |
| **HSDA22** | 2365 | 2202 | 1796 | 1672 | 1642 | 1562 | 1521 | 1515 | 1355 | 1372 | 1033 | 571.4 | 514 | 409 | 365.6 | 350.4 | 328 | 312.3 | 303.1 | 264.5 | 257.3 | 188.7 |
| **HSDA23** | 2356 | 1979 | 1857 | 1782 | 1800 | 1793 | 1753 | 1682 | 1558 | 1438 | 1200 | 456.1 | 371.6 | 335.1 | 310.3 | 303.2 | 296.9 | 277.6 | 260.3 | 233.4 | 208 | 166.9 |
| **HSDA31** | 502 | 421 | 390 | 347 | 383 | 362 | 347 | 355 | 396 | 355 | 256 | 347.9 | 282.5 | 260.6 | 220.9 | 234.5 | 215.7 | 199.3 | 202.5 | 219.8 | 188 | 133.2 |
| **HSDA32** | 1947 | 1854 | 1706 | 1553 | 1437 | 1538 | 1357 | 1329 | 1410 | 1184 | 1104 | 426.6 | 401.4 | 360.5 | 323.1 | 290.4 | 305.9 | 266.1 | 255.1 | 264 | 214.1 | 193.2 |
| **HSDA33** | 990 | 1032 | 972 | 911 | 823 | 888 | 753 | 826 | 771 | 734 | 579 | 413.4 | 427 | 391.7 | 357.8 | 316.2 | 333.6 | 280.4 | 299.7 | 272.4 | 251.2 | 194.5 |
| **HSDA41** | 1538 | 1242 | 1097 | 1122 | 1187 | 1122 | 1025 | 1122 | 1075 | 998 | 864 | 450.3 | 362 | 311.6 | 311 | 321.2 | 296.9 | 263.9 | 282.2 | 265.3 | 241.5 | 204.4 |
| **HSDA42** | 1807 | 1528 | 1439 | 1351 | 1273 | 1226 | 1136 | 1257 | 1266 | 1077 | 913 | 692.1 | 578.9 | 535.5 | 492.6 | 452.4 | 425.8 | 385 | 415 | 405.7 | 340.5 | 281.6 |
| **HSDA43** | 712 | 698 | 549 | 539 | 630 | 567 | 546 | 546 | 606 | 581 | 409 | 642.3 | 616.1 | 470.7 | 453.5 | 514.3 | 447.9 | 425 | 412.9 | 438.6 | 414 | 287.9 |
| **HSDA51** | 305 | 260 | 256 | 295 | 345 | 272 | 252 | 241 | 232 | 281 | 270 | 548.2 | 501.1 | 481.5 | 527 | 590.3 | 474 | 435.1 | 401.1 | 388.7 | 449.6 | 438.3 |
| **HSDA52** | 601 | 644 | 544 | 574 | 573 | 553 | 537 | 482 | 584 | 793 | 662 | 614.7 | 635.7 | 523.4 | 546.7 | 533.5 | 515 | 493.9 | 431.4 | 500.6 | 640.2 | 540.8 |
| **HSDA53** | 357 | 351 | 290 | 216 | 283 | 273 | 249 | 238 | 277 | 265 | 213 | 982.1 | 918.8 | 704.3 | 552.3 | 697.1 | 717.4 | 630.9 | 599.2 | 655.4 | 612.2 | 461.4 |

***** Rates are expressed as new cases per 100,000 persons at risk per year (equivalent to per 100,000 person-years at risk).

Abbreviations: COPD: Chronic Obstructive Pulmonary Disease, HSDA: Health Services Delivery Area; HSDA11: East Kootenay, HSDA 12: Kootenay Boundary, HSDA 13: Okanagan, HSDA 14: Thompson Cariboo Shuswap, HSDA 21: Fraser East, HSDA 22: Fraser North, HSDA 23: Fraser South, HSDA 31: Richmond, HSDA 32: Vancouver, HSDA 33: North Shore/Coast Garibaldi, HSDA 41: South Vancouver Island, HSDA 42: Central Vancouver Island, HSDA 43: North Vancouver Island, HSDA 51: Northwest, HSDA 52: Northern Interior, HSDA 53: Northeast.

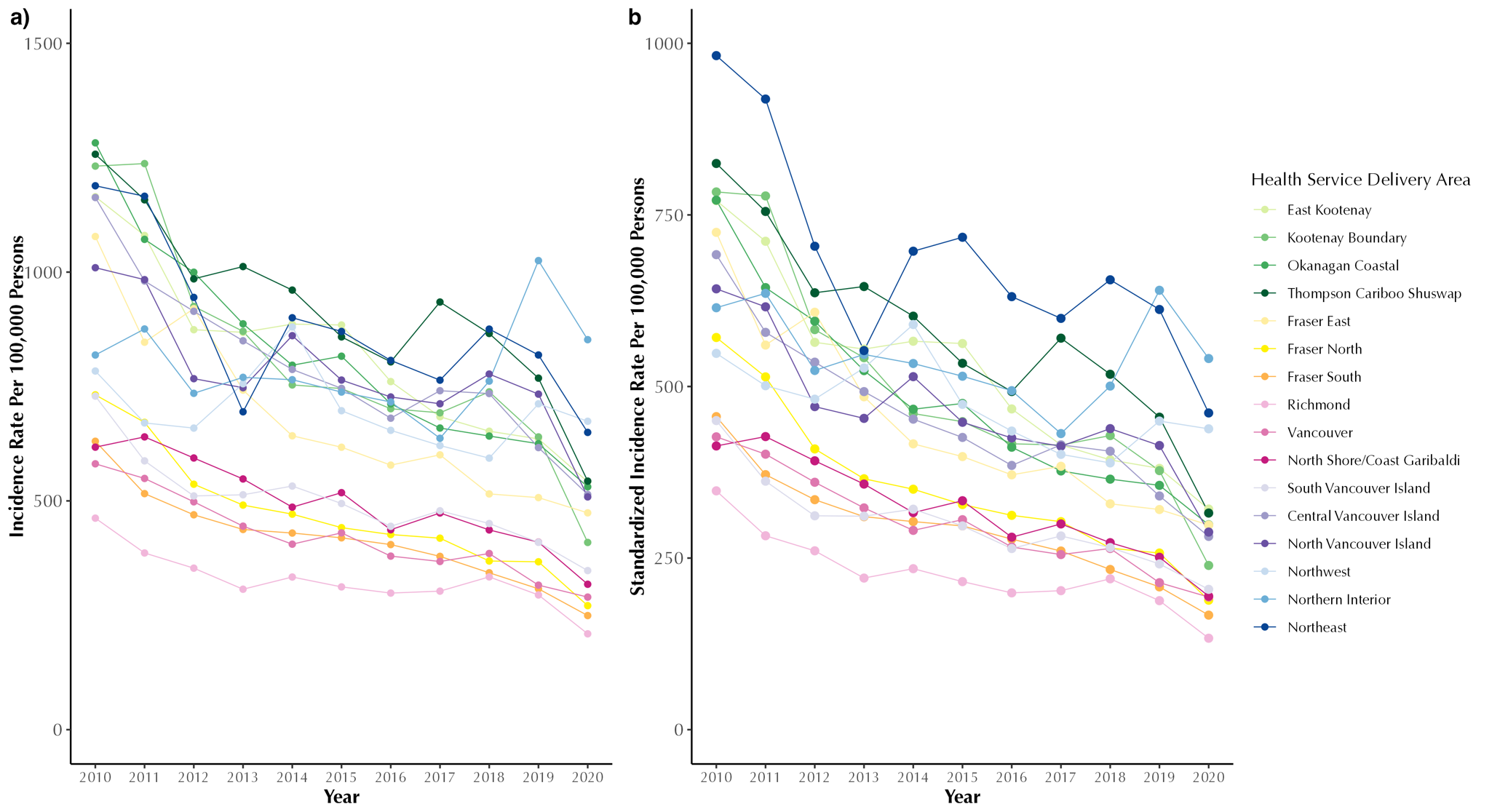

### **Figure A3: Annual crude (a) and sex and age standardized (b) chronic obstructive pulmonary disease incidence rate (per 100,000 persons) in British Columbia, Canada, from 2001 to 2020, stratified by Health Service Delivery Area**

Rates are expressed as new cases per 100,000 persons at risk per year (equivalent to per 100,000 person-years at risk).

### **Table A8: All-cause, COPD-specific and CVD-specific number of hospital admissions and sex and age standardized hospitalization rate (per 100,000 COPD patients) from 2001 to 2020, stratified by Health Service Delivery Area**

| **Admission Year** | | | | | | | | | | | | | | | | | | | | | | |
| --- | --- | --- | --- | --- | --- | --- | --- | --- | --- | --- | --- | --- | --- | --- | --- | --- | --- | --- | --- | --- | --- | --- |
|  | **2010** | **2011** | **2012** | **2013** | **2014** | **2015** | **2016** | **2017** | **2018** | **2019** | **2020** | **2010** | **2011** | **2012** | **2013** | **2014** | **2015** | **2016** | **2017** | **2018** | **2019** | **2020** |
|  | **Number of All-cause hospitalizations** | | | | | | | | | | | **Sex and age-standardized All-cause hospitalization rate (per 100,000 COPD patients)*** | | | | | | | | | | |
| **Total** | 141580 | 145797 | 147532 | 149056 | 146880 | 146589 | 145109 | 141028 | 138170 | 134172 | 111744 | 93165.2 | 87530.8 | 82811.4 | 78038.6 | 73606.7 | 71700.1 | 69258.1 | 65491.5 | 60761.8 | 54878.3 | 43578.4 |
| **HSDA11** | 3800 | 3897 | 3848 | 3751 | 3894 | 4116 | 3706 | 3554 | 3405 | 3284 | 2746 | 114348.3 | 105627.4 | 106101.5 | 94365.6 | 81673.6 | 103769.7 | 81396.2 | 75011.8 | 72308.0 | 63326.8 | 49633.2 |
| **HSDA12** | 4017 | 4051 | 4082 | 4051 | 3961 | 3884 | 3693 | 3516 | 3628 | 3534 | 2811 | 100803.4 | 92574.6 | 82375.2 | 97470.4 | 83927.7 | 75882.9 | 82588.6 | 62311.8 | 57189.1 | 51165.4 | 43479.4 |
| **HSDA13** | 17965 | 18243 | 18361 | 18486 | 18255 | 18504 | 18251 | 16913 | 16882 | 16392 | 13922 | 98400.1 | 99175.6 | 91264.5 | 85921.7 | 80083.3 | 80079.0 | 76865.0 | 63073.7 | 62142.3 | 56132.1 | 44676.0 |
| **HSDA14** | 11582 | 12066 | 12062 | 12447 | 12381 | 12089 | 11748 | 11587 | 11850 | 11349 | 9318 | 104611.5 | 103901.4 | 101460.6 | 99191.4 | 80769.8 | 84139.8 | 84440.9 | 76475.6 | 71856.4 | 63609.7 | 48779.3 |
| **HSDA21** | 10908 | 10889 | 11434 | 11393 | 11108 | 11117 | 11069 | 10646 | 10470 | 10265 | 8608 | 112001.7 | 97038.2 | 91239.5 | 86466.7 | 76861.8 | 74029.0 | 78135.9 | 67551.6 | 61776.5 | 58268.6 | 46291.8 |
| **HSDA22** | 15577 | 16642 | 16547 | 16752 | 16444 | 16866 | 16312 | 15945 | 15296 | 15067 | 12518 | 87611.3 | 80239.1 | 76087.6 | 69778.3 | 65042.3 | 63502.6 | 60312.2 | 59048.7 | 54672.5 | 51786.3 | 41728.7 |
| **HSDA23** | 17824 | 18436 | 18989 | 19177 | 19267 | 19660 | 19461 | 19204 | 18664 | 18008 | 14618 | 84483.7 | 79945.6 | 81667.2 | 73888.7 | 73366.0 | 72395.4 | 68930.3 | 64375.9 | 61061.7 | 56837.1 | 43659.1 |
| **HSDA31** | 3290 | 3403 | 3314 | 3258 | 3288 | 3310 | 3435 | 3146 | 3135 | 2867 | 2369 | 61933.5 | 68018.6 | 56894.9 | 57015.8 | 63724.3 | 64097.9 | 65955.6 | 60351.2 | 57916.7 | 56458.0 | 37994.4 |
| **HSDA32** | 13527 | 13528 | 14287 | 14644 | 13817 | 13353 | 12944 | 12689 | 12441 | 11520 | 9683 | 75461.7 | 70618.1 | 68395.4 | 74479.8 | 70712.7 | 67118.4 | 62029.5 | 63566.4 | 61347.9 | 54065.9 | 44555.4 |
| **HSDA33** | 7674 | 7679 | 7590 | 8065 | 7330 | 7430 | 7430 | 7223 | 6997 | 6876 | 5379 | 96271.2 | 87436.5 | 76859.0 | 74224.2 | 69728.3 | 63868.4 | 62456.3 | 59161.7 | 50677.5 | 45674.1 | 37233.6 |
| **HSDA41** | 9603 | 10199 | 10290 | 10290 | 10564 | 10324 | 10391 | 10197 | 9734 | 9454 | 8015 | 90774.6 | 81701.1 | 79036.2 | 74350.4 | 80365.0 | 70783.5 | 68226.6 | 63967.3 | 58793.9 | 52245.7 | 40264.3 |
| **HSDA42** | 10325 | 10777 | 11238 | 11290 | 11230 | 10797 | 11132 | 11187 | 10500 | 10541 | 9303 | 93076.5 | 91864.8 | 81499.2 | 69285.5 | 66824.9 | 61077.7 | 59755.6 | 58815.6 | 51730.5 | 48335.8 | 40783.5 |
| **HSDA43** | 4806 | 5039 | 4838 | 4783 | 4766 | 4955 | 5082 | 4796 | 4620 | 4978 | 4142 | 113884.0 | 102333.4 | 101199.0 | 91278.9 | 85384.0 | 79009.6 | 67346.9 | 64915.7 | 53235.0 | 53226.2 | 44379.0 |
| **HSDA51** | 3008 | 3084 | 2736 | 2740 | 2738 | 2692 | 2749 | 2728 | 2682 | 2504 | 2142 | 150937.5 | 140920.5 | 132767.0 | 111122.4 | 92934.9 | 81597.0 | 101553.3 | 92827.1 | 79456.6 | 64175.0 | 51592.6 |
| **HSDA52** | 5373 | 5652 | 5783 | 5827 | 5698 | 5470 | 5597 | 5479 | 5578 | 5469 | 4535 | 130518.2 | 111357.9 | 105294.6 | 99180.3 | 92553.2 | 103947.6 | 102398.4 | 98051.0 | 83635.9 | 62741.4 | 45166.9 |
| **HSDA53** | 2106 | 2047 | 1957 | 1953 | 1992 | 1873 | 1962 | 2038 | 2064 | 1913 | 1544 | 93615.2 | 90665.8 | 85840.8 | 70441.4 | 66358.5 | 73944.6 | 66561.9 | 74225.0 | 68692.4 | 56739.6 | 42757.1 |
|  | **Number of COPD-specific hospitalizations** | | | | | | | | | | | **Sex and age-standardized COPD-specific hospitalization rate (per 100,000 COPD patients)*** | | | | | | | | | | |
| **Total** | 8379 | 8942 | 8912 | 9423 | 9484 | 10032 | 9986 | 10527 | 9893 | 9347 | 6647 | 3749.2 | 3518.0 | 3264.5 | 3664.4 | 3484.8 | 3439.2 | 3435.0 | 3671.0 | 3396.5 | 3028.3 | 2179.7 |
| **HSDA11** | 200 | 212 | 256 | 278 | 285 | 315 | 252 | 292 | 258 | 225 | 181 | 3639.6 | 4044.8 | 3864.9 | 5220.7 | 3787.5 | 3768.6 | 3139.6 | 3877.7 | 4020.9 | 2974.4 | 3591.9 |
| **HSDA12** | 212 | 195 | 221 | 222 | 198 | 221 | 214 | 239 | 242 | 216 | 183 | 3845.8 | 2876.1 | 3415.1 | 3132.3 | 3078.0 | 2902.7 | 2648.8 | 3001.5 | 2853.2 | 2916.4 | 2638.6 |
| **HSDA13** | 1062 | 1074 | 1070 | 1056 | 1096 | 1132 | 1217 | 1281 | 1199 | 1162 | 851 | 3767.3 | 3765.6 | 3147.3 | 2964.9 | 2957.2 | 3123.2 | 3329.5 | 3991.8 | 3486.2 | 3025.6 | 2334.4 |
| **HSDA14** | 606 | 700 | 672 | 734 | 736 | 812 | 817 | 829 | 790 | 812 | 573 | 3530.1 | 3837.5 | 3093.2 | 3698.2 | 3089.0 | 3386.9 | 3555.6 | 3234.1 | 4164.4 | 3021.1 | 2353.5 |
| **HSDA21** | 595 | 611 | 708 | 751 | 786 | 757 | 807 | 842 | 703 | 653 | 510 | 4831.9 | 3998.7 | 4063.9 | 4533.9 | 4438.6 | 4298.8 | 4055.0 | 4202.1 | 3371.3 | 2912.6 | 2268.8 |
| **HSDA22** | 971 | 1028 | 977 | 1053 | 959 | 1099 | 1118 | 1101 | 988 | 907 | 616 | 3463.1 | 3416.1 | 2965.3 | 3179.0 | 2935.2 | 3007.5 | 3009.2 | 2923.0 | 2838.6 | 2540.1 | 1763.2 |
| **HSDA23** | 1040 | 1143 | 1203 | 1334 | 1405 | 1620 | 1590 | 1619 | 1460 | 1398 | 950 | 3653.8 | 3559.1 | 3711.1 | 4544.5 | 4170.1 | 4278.9 | 4583.4 | 4739.1 | 3463.6 | 3575.5 | 2401.4 |
| **HSDA31** | 194 | 229 | 191 | 186 | 217 | 224 | 254 | 240 | 255 | 208 | 118 | 2371.4 | 3483.5 | 2415.7 | 2069.7 | 3421.8 | 3719.1 | 2500.3 | 2283.6 | 3131.1 | 1892.5 | 1230.6 |
| **HSDA32** | 1023 | 997 | 948 | 1041 | 1050 | 975 | 928 | 1067 | 945 | 843 | 602 | 4707.9 | 3585.4 | 3435.4 | 4212.0 | 3985.0 | 3424.5 | 3711.4 | 4434.4 | 4151.9 | 3582.0 | 2396.7 |
| **HSDA33** | 468 | 498 | 433 | 476 | 472 | 462 | 422 | 464 | 414 | 429 | 267 | 3147.8 | 3289.9 | 2564.0 | 2768.4 | 3173.8 | 2736.3 | 2295.2 | 3198.1 | 2337.6 | 2727.0 | 1324.5 |
| **HSDA41** | 537 | 673 | 655 | 721 | 699 | 710 | 696 | 792 | 704 | 697 | 453 | 2512.3 | 3020.1 | 2681.8 | 3863.8 | 3744.9 | 3090.4 | 3046.0 | 3532.4 | 2724.4 | 3026.5 | 1981.2 |
| **HSDA42** | 601 | 616 | 645 | 636 | 667 | 630 | 652 | 735 | 733 | 666 | 495 | 2970.6 | 2759.6 | 2788.6 | 2467.4 | 2869.1 | 2577.1 | 2621.1 | 2730.7 | 2898.4 | 2158.0 | 1643.9 |
| **HSDA43** | 230 | 301 | 260 | 273 | 268 | 322 | 304 | 281 | 345 | 351 | 270 | 2716.9 | 3283.2 | 2452.3 | 3334.5 | 3826.1 | 4852.2 | 2664.6 | 2277.7 | 3203.0 | 3089.7 | 2121.9 |
| **HSDA51** | 158 | 168 | 169 | 179 | 136 | 171 | 134 | 159 | 195 | 155 | 126 | 5096.0 | 4197.7 | 5028.9 | 4606.3 | 2687.4 | 3477.7 | 2781.1 | 4891.8 | 3679.9 | 3299.5 | 2482.6 |
| **HSDA52** | 333 | 371 | 397 | 365 | 370 | 451 | 402 | 382 | 449 | 421 | 308 | 4435.4 | 4415.2 | 4340.0 | 3922.5 | 4021.3 | 4431.5 | 3769.4 | 3496.0 | 4340.4 | 4061.0 | 2619.3 |
| **HSDA53** | 135 | 107 | 94 | 110 | 132 | 123 | 171 | 193 | 194 | 184 | 136 | 5050.1 | 3406.3 | 2515.4 | 3964.0 | 3223.5 | 3121.6 | 4867.6 | 5296.8 | 4675.5 | 3865.0 | 2772.7 |
|  | **Number of CVD-specific hospitalizations** | | | | | | | | | | | **Sex and age-standardized CVD-specific hospitalization rate (per 100,000 COPD patients) *** | | | | | | | | | | |
| **Total** | 19040 | 19068 | 18150 | 18190 | 18168 | 17626 | 17521 | 17343 | 16570 | 16194 | 14005 | 9157.4 | 8342.0 | 7187.4 | 6848.8 | 6785.3 | 6235.6 | 5886.6 | 5697.9 | 5172.9 | 5006.0 | 4250.3 |
| **HSDA11** | 459 | 481 | 420 | 437 | 434 | 494 | 464 | 440 | 381 | 366 | 351 | 9674.8 | 7552.6 | 7961.3 | 6600.9 | 6690.4 | 11621.9 | 7288.2 | 7303.1 | 4543.4 | 4990.3 | 4709.3 |
| **HSDA12** | 567 | 583 | 514 | 517 | 522 | 473 | 468 | 457 | 462 | 382 | 329 | 9600.3 | 10760.1 | 9184.9 | 9062.2 | 9774.6 | 6071.7 | 5523.5 | 5183.1 | 5044.3 | 3962.4 | 3569.7 |
| **HSDA13** | 2366 | 2313 | 2111 | 2135 | 2153 | 2132 | 2020 | 1973 | 1820 | 1822 | 1664 | 7864.2 | 7697.7 | 6777.6 | 7771.6 | 6244.7 | 5537.3 | 5430.4 | 5370.9 | 4929.6 | 4597.2 | 4050.4 |
| **HSDA14** | 1476 | 1492 | 1363 | 1430 | 1513 | 1378 | 1369 | 1406 | 1361 | 1302 | 1097 | 9581.6 | 8442.5 | 7932.8 | 8762.6 | 7223.9 | 8046.6 | 5880.7 | 6353.5 | 6295.5 | 5605.0 | 4889.1 |
| **HSDA21** | 1602 | 1589 | 1584 | 1587 | 1653 | 1584 | 1420 | 1460 | 1363 | 1367 | 1201 | 11875.3 | 11308.0 | 9565.5 | 8477.3 | 8535.5 | 8354.9 | 6697.6 | 6864.5 | 5729.2 | 5515.7 | 5097.8 |
| **HSDA22** | 2061 | 2141 | 2029 | 1975 | 1952 | 1949 | 2029 | 1885 | 1859 | 1735 | 1510 | 8505.5 | 7924.9 | 6546.5 | 6120.3 | 5684.2 | 6227.4 | 5572.7 | 4738.8 | 4742.0 | 4889.2 | 3718.4 |
| **HSDA23** | 2642 | 2620 | 2469 | 2519 | 2586 | 2502 | 2410 | 2600 | 2460 | 2351 | 1916 | 9993.3 | 9217.9 | 7588.9 | 7124.3 | 8015.4 | 6461.4 | 6483.3 | 6213.5 | 5593.0 | 5271.9 | 4502.7 |
| **HSDA31** | 503 | 423 | 469 | 445 | 489 | 464 | 472 | 405 | 385 | 374 | 290 | 6928.4 | 5430.9 | 7055.5 | 5852.6 | 5613.5 | 5493.5 | 6283.6 | 4053.7 | 4558.7 | 3760.8 | 2572.6 |
| **HSDA32** | 1643 | 1717 | 1639 | 1605 | 1578 | 1478 | 1493 | 1407 | 1312 | 1295 | 1074 | 6670.1 | 6343.6 | 5767.4 | 5399.3 | 5320.3 | 4398.3 | 4915.1 | 4952.9 | 4451.8 | 4059.2 | 3457.7 |
| **HSDA33** | 1034 | 1070 | 1008 | 1106 | 972 | 966 | 963 | 956 | 893 | 843 | 723 | 9134.0 | 8783.5 | 6999.0 | 7024.0 | 6333.9 | 5835.7 | 5262.0 | 5966.2 | 4150.5 | 4107.2 | 4235.0 |
| **HSDA41** | 1115 | 1072 | 1072 | 1077 | 1080 | 1024 | 1106 | 1080 | 1086 | 1114 | 993 | 6899.0 | 5025.3 | 4777.7 | 4833.0 | 4650.9 | 4521.3 | 4982.4 | 4672.2 | 4394.3 | 4800.1 | 3971.6 |
| **HSDA42** | 1425 | 1352 | 1404 | 1463 | 1307 | 1269 | 1368 | 1314 | 1302 | 1300 | 1236 | 8038.9 | 7121.0 | 6051.5 | 5776.9 | 5697.5 | 4812.2 | 5556.8 | 4882.0 | 4385.6 | 4126.8 | 3638.4 |
| **HSDA43** | 673 | 726 | 680 | 608 | 672 | 687 | 668 | 673 | 625 | 728 | 630 | 10147.4 | 12464.8 | 7442.3 | 8349.7 | 7472.2 | 8518.8 | 6098.9 | 6125.8 | 5254.0 | 6993.0 | 4639.9 |
| **HSDA51** | 439 | 450 | 398 | 358 | 357 | 334 | 366 | 360 | 296 | 277 | 237 | 14286.9 | 13097.5 | 21071.7 | 9391.8 | 10823.4 | 7200.9 | 7829.6 | 7050.9 | 6601.0 | 5407.2 | 4648.9 |
| **HSDA52** | 685 | 732 | 675 | 659 | 609 | 609 | 640 | 654 | 679 | 666 | 530 | 13522.0 | 10078.1 | 8775.5 | 8989.2 | 6940.0 | 6689.5 | 6035.6 | 7223.9 | 6817.4 | 5727.7 | 4753.7 |
| **HSDA53** | 321 | 284 | 294 | 241 | 275 | 273 | 246 | 260 | 254 | 252 | 217 | 12605.9 | 8785.5 | 9257.3 | 6524.8 | 8879.9 | 6338.2 | 6034.4 | 6448.9 | 6368.4 | 5959.4 | 5498.5 |

***** Rates are expressed as hospitalizations per 100,000 persons with COPD per year (equivalent to per 100,000 COPD patient-years).

Abbreviations: COPD: Chronic Obstructive Pulmonary Disease, CVD: Cardio-vascular Disease; HSDA: Health Services Delivery Area; HSDA11: East Kootenay, HSDA 12: Kootenay Boundary, HSDA 13: Okanagan, HSDA 14: Thompson Cariboo Shuswap, HSDA 21: Fraser East, HSDA 22: Fraser North, HSDA 23: Fraser South, HSDA 31: Richmond, HSDA 32: Vancouver, HSDA 33: North Shore/Coast Garibaldi, HSDA 41: South Vancouver Island, HSDA 42: Central Vancouver Island, HSDA 43: North Vancouver Island, HSDA 51: Northwest, HSDA 52: Northern Interior, HSDA 53: Northeast

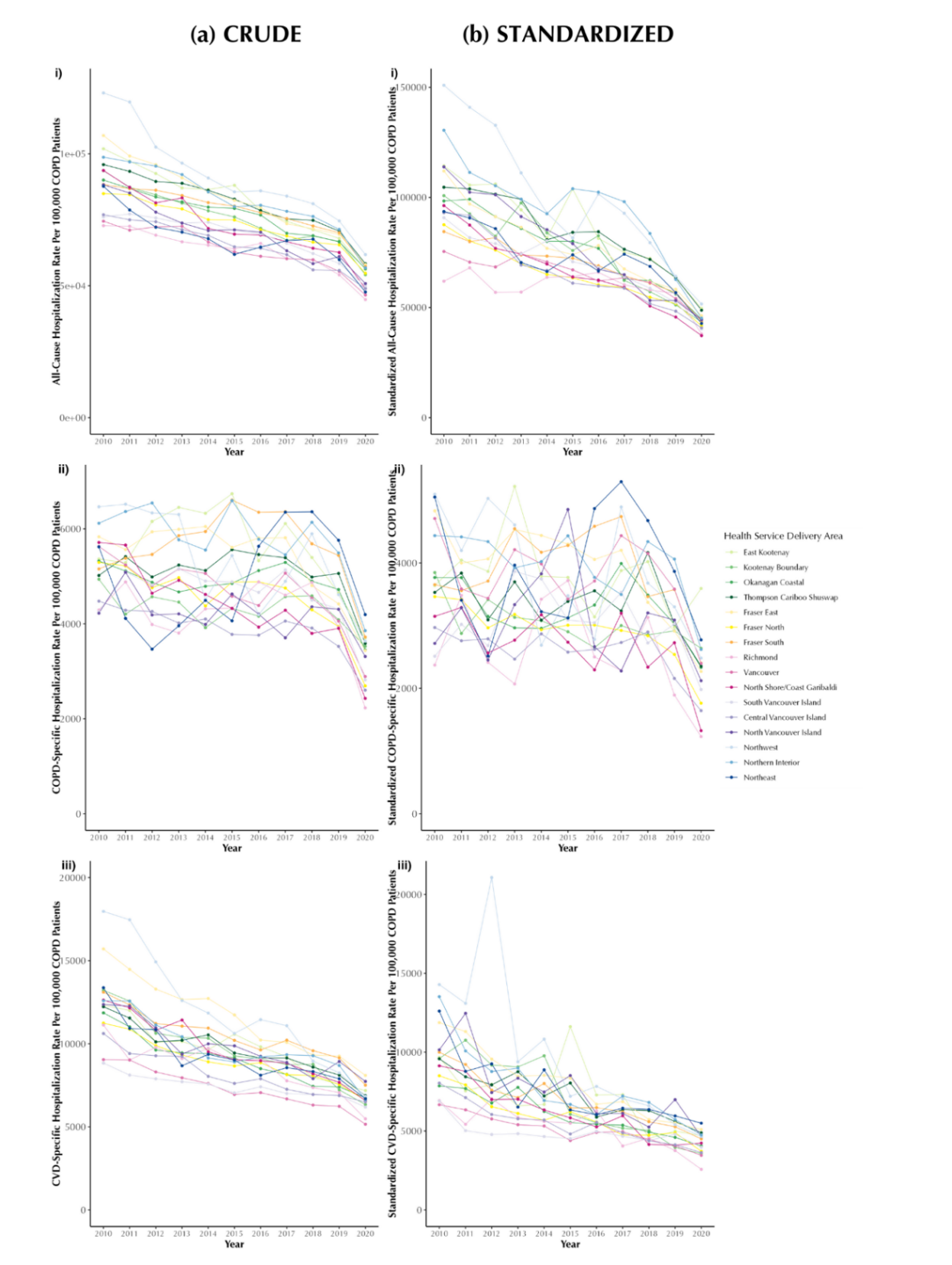

**All-cause Hospitalization**

**COPD-Specific Hospitalization**

**CVD-specific Hospitalization**

### **Figure A4: Annual crude (a) and age- and sex-standardized (b) all-cause (i), COPD-specific (ii), and CVD-specific (iii) hospitalization rates (per 100,000 persons with COPD) in British Columbia, Canada, 2001–2020, stratified by Health Service Delivery Area.**

Rates are expressed as hospitalizations per 100,000 persons with COPD per year (equivalent to per 100,000 COPD patient-years).

### **Table A9: Crude mortality count and sex and age standardized mortality rate (per 100,000 COPD patient) from 2001 to 2020, stratified by Health Service Delivery Area**

| **Year of Death** | | | | | | | | | | | | | | | | | | | | | | |
| --- | --- | --- | --- | --- | --- | --- | --- | --- | --- | --- | --- | --- | --- | --- | --- | --- | --- | --- | --- | --- | --- | --- |
|  | **2010** | **2011** | **2012** | **2013** | **2014** | **2015** | **2016** | **2017** | **2018** | **2019** | **2020** | **2010** | **2011** | **2012** | **2013** | **2014** | **2015** | **2016** | **2017** | **2018** | **2019** | **2020** |
|  | **Number of All-cause deaths** | | | | | | | | | | | **Sex and age-standardized All-cause mortality rate (per 100,000 COPD patients)** | | | | | | | | | | |
| **Total** | 7871 | 8518 | 8802 | 9046 | 9371 | 9981 | 10171 | 10536 | 10652 | 10723 | 10941 | 2435.3 | 2406.2 | 2333.1 | 2325.0 | 2360.7 | 2418.1 | 2441.9 | 2414.3 | 2455.8 | 2394.9 | 2517.1 |
| **HSDA11** | 190 | 227 | 224 | 202 | 215 | 301 | 262 | 276 | 241 | 245 | 254 | 2405.4 | 2792.3 | 2462.4 | 2264.5 | 2341.3 | 3006.9 | 2797.0 | 2576.5 | 2501.0 | 2454.9 | 2530.6 |
| **HSDA12** | 237 | 223 | 249 | 262 | 263 | 253 | 266 | 266 | 274 | 315 | 281 | 3150.6 | 2324.4 | 2708.1 | 2686.9 | 2742.1 | 2559.4 | 2740.7 | 2456.8 | 2483.6 | 2536.0 | 2776.2 |
| **HSDA13** | 1106 | 1156 | 1219 | 1236 | 1157 | 1307 | 1353 | 1391 | 1356 | 1348 | 1386 | 2613.8 | 2411.3 | 2492.9 | 2453.9 | 2237.4 | 2614.8 | 2622.9 | 2490.3 | 2595.6 | 2360.5 | 2637.5 |
| **HSDA14** | 550 | 620 | 666 | 715 | 757 | 775 | 813 | 844 | 843 | 866 | 814 | 2398.6 | 2617.0 | 2639.7 | 2596.2 | 2810.7 | 3112.0 | 2906.6 | 2736.8 | 2575.1 | 2659.4 | 2604.6 |
| **HSDA21** | 489 | 493 | 557 | 568 | 645 | 693 | 703 | 774 | 746 | 843 | 760 | 2605.5 | 2170.4 | 2421.4 | 2372.9 | 2594.6 | 2459.7 | 2404.7 | 2999.6 | 2635.4 | 2859.5 | 2559.4 |
| **HSDA22** | 811 | 876 | 938 | 910 | 1004 | 1090 | 1077 | 1114 | 1152 | 1074 | 1206 | 2217.8 | 2147.5 | 2171.3 | 2109.6 | 2167.5 | 2200.7 | 2183.2 | 2039.3 | 2139.3 | 2240.6 | 2474.3 |
| **HSDA23** | 963 | 1061 | 1073 | 1097 | 1152 | 1289 | 1294 | 1286 | 1368 | 1330 | 1393 | 2376.8 | 2525.2 | 2359.7 | 2399.5 | 2279.0 | 2389.0 | 2351.8 | 2466.6 | 2449.5 | 2268.9 | 2370.3 |
| **HSDA31** | 217 | 226 | 222 | 214 | 191 | 206 | 218 | 192 | 220 | 211 | 223 | 2102.7 | 2016.4 | 2088.2 | 1843.3 | 1555.7 | 2308.9 | 1744.1 | 1426.3 | 3158.1 | 1641.7 | 1639.5 |
| **HSDA32** | 843 | 907 | 930 | 967 | 969 | 1025 | 1004 | 1096 | 1036 | 1005 | 1105 | 2256.7 | 2340.0 | 2168.0 | 2213.5 | 2149.5 | 2239.1 | 2402.3 | 2449.3 | 2569.2 | 2471.8 | 2789.3 |
| **HSDA33** | 417 | 445 | 471 | 482 | 511 | 565 | 515 | 580 | 557 | 578 | 545 | 2501.6 | 2142.2 | 2132.8 | 1948.8 | 2003.3 | 2797.5 | 1913.9 | 2150.5 | 2056.3 | 2051.9 | 2010.9 |
| **HSDA41** | 675 | 757 | 742 | 768 | 821 | 805 | 843 | 906 | 923 | 916 | 913 | 2241.0 | 2612.3 | 2317.9 | 2304.0 | 2925.9 | 2185.5 | 2625.8 | 2252.9 | 2393.4 | 2464.2 | 2571.3 |
| **HSDA42** | 629 | 670 | 665 | 749 | 797 | 774 | 846 | 842 | 917 | 905 | 949 | 2542.3 | 2318.5 | 2105.4 | 2222.2 | 2378.4 | 2180.9 | 2409.4 | 2293.9 | 2438.9 | 2162.2 | 2680.1 |
| **HSDA43** | 256 | 291 | 301 | 318 | 300 | 306 | 342 | 377 | 355 | 434 | 422 | 2490.5 | 2490.6 | 2405.0 | 2435.9 | 2332.8 | 2420.4 | 2236.9 | 2708.2 | 2209.5 | 2512.3 | 2445.2 |
| **HSDA51** | 112 | 140 | 112 | 132 | 133 | 123 | 139 | 137 | 140 | 144 | 158 | 2512.1 | 2950.7 | 2358.3 | 2652.2 | 2373.8 | 2022.5 | 2428.5 | 2188.5 | 2290.4 | 2574.1 | 2293.6 |
| **HSDA52** | 231 | 275 | 289 | 279 | 329 | 345 | 371 | 323 | 376 | 369 | 406 | 2683.0 | 2859.1 | 2670.4 | 2524.6 | 2721.2 | 2827.0 | 3047.8 | 2544.4 | 2827.0 | 2568.7 | 2894.1 |
| **HSDA53** | 116 | 124 | 113 | 124 | 127 | 124 | 125 | 132 | 143 | 131 | 112 | 3040.8 | 2905.0 | 2560.9 | 2699.4 | 2854.2 | 2550.1 | 2536.0 | 2540.3 | 2768.6 | 2439.6 | 2290.8 |
|  | **Number of COPD-specific deaths** | | | | | | | | | | | **Sex and age-standardized COPD-specific mortality rate (per 100,000 COPD patients)** | | | | | | | | | | |
| **Total** | 1283 | 1364 | 1241 | 1368 | 1411 | 1542 | 1604 | 1607 | 1597 | 1567 | 1460 | 378.0 | 365.5 | 309.3 | 330.1 | 333.0 | 358.5 | 343.0 | 332.5 | 343.9 | 318.9 | 302.5 |
| **HSDA11** | 41 | 28 | 33 | 42 | 19 | 55 | 52 | 43 | 33 | 44 | 44 | 469.0 | 346.9 | 324.8 | 458.8 | 149.3 | 587.6 | 435.3 | 353.9 | 382.0 | 474.6 | 367.1 |
| **HSDA12** | 40 | 31 | 32 | 33 | 38 | 29 | 41 | 34 | 37 | 45 | 41 | 445.4 | 296.6 | 288.3 | 311.9 | 363.6 | 238.3 | 431.0 | 280.3 | 303.6 | 420.9 | 345.2 |
| **HSDA13** | 163 | 181 | 158 | 158 | 153 | 168 | 176 | 206 | 196 | 165 | 184 | 343.3 | 348.6 | 285.1 | 289.1 | 288.4 | 317.9 | 275.8 | 347.0 | 357.8 | 247.1 | 316.1 |
| **HSDA14** | 91 | 115 | 106 | 109 | 102 | 115 | 123 | 123 | 120 | 143 | 109 | 341.7 | 442.3 | 372.2 | 359.3 | 320.2 | 374.5 | 348.2 | 366.9 | 358.8 | 409.0 | 310.7 |
| **HSDA21** | 94 | 70 | 84 | 106 | 102 | 103 | 105 | 126 | 118 | 116 | 124 | 459.3 | 280.1 | 401.9 | 404.8 | 369.0 | 337.7 | 312.4 | 408.7 | 382.1 | 341.4 | 362.5 |
| **HSDA22** | 133 | 116 | 132 | 135 | 147 | 155 | 173 | 174 | 157 | 151 | 140 | 441.2 | 287.0 | 300.7 | 277.5 | 376.0 | 296.7 | 296.6 | 277.6 | 273.7 | 277.4 | 226.9 |
| **HSDA23** | 150 | 164 | 146 | 154 | 165 | 216 | 200 | 186 | 194 | 190 | 153 | 342.0 | 337.3 | 279.1 | 320.7 | 286.5 | 363.0 | 342.4 | 298.7 | 341.4 | 293.7 | 230.5 |
| **HSDA31** | 39 | 43 | 31 | 28 | 26 | 22 | 27 | 23 | 30 | 28 | 24 | 306.1 | 348.8 | 240.8 | 204.4 | 163.9 | 117.1 | 185.1 | 150.9 | 221.6 | 202.6 | 150.0 |
| **HSDA32** | 136 | 151 | 125 | 151 | 149 | 155 | 138 | 155 | 136 | 120 | 124 | 328.8 | 469.8 | 286.9 | 335.6 | 294.7 | 412.2 | 286.7 | 317.1 | 317.7 | 257.8 | 250.7 |
| **HSDA33** | 58 | 83 | 65 | 63 | 70 | 95 | 83 | 87 | 95 | 85 | 69 | 238.6 | 352.9 | 238.6 | 240.4 | 238.2 | 335.4 | 312.5 | 273.7 | 293.5 | 290.3 | 242.5 |
| **HSDA41** | 109 | 129 | 105 | 115 | 157 | 133 | 156 | 161 | 147 | 147 | 111 | 410.8 | 339.6 | 271.0 | 307.4 | 397.7 | 350.1 | 386.8 | 355.4 | 380.6 | 343.8 | 291.5 |
| **HSDA42** | 100 | 111 | 99 | 124 | 127 | 138 | 142 | 131 | 153 | 158 | 154 | 385.3 | 341.6 | 287.1 | 369.1 | 349.2 | 345.5 | 409.9 | 329.2 | 371.9 | 352.3 | 351.6 |
| **HSDA43** | 42 | 55 | 49 | 54 | 48 | 61 | 68 | 66 | 64 | 73 | 65 | 352.9 | 463.7 | 402.7 | 379.2 | 373.4 | 414.1 | 430.0 | 544.8 | 329.2 | 372.8 | 344.4 |
| **HSDA51** | 20 | 22 | 9 | 21 | 14 | 20 | 22 | 13 | 20 | 23 | 28 | 418.0 | 414.0 | 169.9 | 429.0 | 222.2 | 302.1 | 340.3 | 191.3 | 286.7 | 342.0 | 399.7 |
| **HSDA52** | 41 | 42 | 52 | 50 | 74 | 55 | 69 | 63 | 75 | 61 | 69 | 402.4 | 370.0 | 427.7 | 421.8 | 611.7 | 423.6 | 499.6 | 466.4 | 482.5 | 405.0 | 441.7 |
| **HSDA53** | 22 | 19 | 14 | 22 | 20 | 22 | 29 | 16 | 22 | 16 | 19 | 525.8 | 453.2 | 308.4 | 451.0 | 387.1 | 426.7 | 530.6 | 275.8 | 390.4 | 274.5 | 618.5 |
|  | **Number of CVD-specific deaths** | | | | | | | | | | | **Sex and age-standardized CVD-specific mortality rate (per 100,000 COPD patients)** | | | | | | | | | | |
| **Total** | 2258 | 2288 | 2424 | 2449 | 2409 | 2609 | 2657 | 2757 | 2780 | 2690 | 2702 | 609.3 | 566.3 | 566.6 | 541.1 | 510.3 | 536.0 | 525.6 | 563.3 | 527.0 | 507.6 | 501.1 |
| **HSDA11** | 41 | 55 | 64 | 50 | 60 | 79 | 66 | 69 | 60 | 60 | 67 | 482.8 | 556.7 | 670.4 | 493.0 | 632.2 | 707.3 | 520.0 | 535.0 | 504.8 | 496.9 | 534.7 |
| **HSDA12** | 77 | 68 | 72 | 73 | 84 | 71 | 79 | 78 | 82 | 92 | 80 | 759.9 | 602.9 | 605.6 | 659.5 | 794.5 | 587.1 | 639.6 | 587.6 | 676.4 | 671.2 | 711.8 |
| **HSDA13** | 322 | 325 | 360 | 362 | 312 | 399 | 375 | 437 | 381 | 367 | 343 | 630.0 | 567.8 | 575.4 | 576.0 | 475.4 | 615.5 | 586.5 | 700.8 | 584.0 | 551.2 | 517.9 |
| **HSDA14** | 149 | 168 | 165 | 186 | 182 | 188 | 219 | 216 | 210 | 214 | 204 | 659.3 | 573.2 | 530.4 | 665.5 | 558.9 | 906.6 | 621.6 | 595.7 | 602.8 | 669.7 | 656.5 |
| **HSDA21** | 129 | 131 | 150 | 151 | 177 | 188 | 175 | 180 | 176 | 206 | 166 | 677.6 | 530.4 | 612.0 | 548.3 | 582.2 | 614.2 | 492.3 | 852.9 | 491.1 | 642.2 | 446.1 |
| **HSDA22** | 222 | 219 | 239 | 222 | 246 | 267 | 270 | 282 | 286 | 236 | 283 | 528.5 | 466.7 | 492.7 | 431.1 | 446.0 | 452.9 | 472.9 | 499.1 | 457.7 | 383.6 | 480.9 |
| **HSDA23** | 305 | 284 | 276 | 274 | 287 | 338 | 338 | 343 | 351 | 322 | 328 | 684.8 | 563.3 | 610.8 | 500.3 | 475.4 | 531.7 | 547.9 | 550.0 | 525.6 | 459.5 | 461.3 |
| **HSDA31** | 66 | 76 | 62 | 54 | 47 | 47 | 53 | 51 | 45 | 48 | 44 | 607.1 | 586.6 | 487.2 | 453.3 | 337.7 | 322.6 | 335.2 | 393.0 | 333.5 | 424.8 | 234.7 |
| **HSDA32** | 227 | 223 | 234 | 251 | 230 | 237 | 231 | 244 | 238 | 244 | 238 | 504.1 | 495.4 | 453.5 | 503.6 | 411.7 | 395.0 | 413.3 | 416.1 | 412.4 | 405.0 | 400.9 |
| **HSDA33** | 114 | 119 | 142 | 150 | 141 | 147 | 134 | 162 | 150 | 152 | 138 | 525.4 | 537.3 | 632.9 | 501.0 | 504.4 | 472.2 | 400.2 | 560.8 | 441.8 | 407.1 | 452.8 |
| **HSDA41** | 208 | 209 | 247 | 244 | 222 | 227 | 239 | 237 | 280 | 228 | 272 | 500.8 | 536.1 | 580.8 | 589.0 | 549.2 | 474.3 | 518.3 | 494.4 | 575.5 | 465.4 | 511.8 |
| **HSDA42** | 201 | 189 | 195 | 204 | 218 | 199 | 231 | 233 | 265 | 248 | 250 | 665.9 | 581.2 | 519.7 | 524.7 | 578.2 | 451.4 | 563.2 | 526.3 | 596.6 | 529.0 | 496.9 |
| **HSDA43** | 69 | 72 | 82 | 79 | 67 | 79 | 85 | 86 | 81 | 121 | 121 | 562.5 | 560.9 | 545.0 | 489.1 | 408.5 | 454.6 | 457.9 | 466.7 | 416.4 | 620.7 | 643.2 |
| **HSDA51** | 24 | 34 | 23 | 37 | 36 | 25 | 36 | 33 | 43 | 31 | 36 | 517.3 | 740.7 | 444.0 | 643.3 | 605.5 | 431.2 | 639.9 | 561.0 | 628.0 | 482.6 | 479.3 |
| **HSDA52** | 50 | 75 | 80 | 72 | 62 | 88 | 92 | 75 | 92 | 84 | 103 | 553.4 | 856.4 | 745.9 | 596.1 | 467.3 | 693.5 | 621.7 | 542.8 | 635.1 | 501.4 | 667.9 |
| **HSDA53** | 45 | 32 | 21 | 34 | 38 | 30 | 34 | 31 | 37 | 36 | 25 | 1100.7 | 774.2 | 419.4 | 729.7 | 740.3 | 555.6 | 698.4 | 579.3 | 626.0 | 590.6 | 385.2 |
| ***** Rates are expressed as hospitalizations per 100,000 persons with COPD per year (equivalent to per 100,000 COPD patient-years).  Abbreviations: COPD: Chronic Obstructive Pulmonary Disease, CVD: Cardio-vascular Disease; HSDA: Health Services Delivery Area; HSDA11: East Kootenay, HSDA 12: Kootenay Boundary, HSDA 13: Okanagan, HSDA 14: Thompson Cariboo Shuswap, HSDA 21: Fraser East, HSDA 22: Fraser North, HSDA 23: Fraser South, HSDA 31: Richmond, HSDA 32: Vancouver, HSDA 33: North Shore/Coast Garibaldi, HSDA 41: South Vancouver Island, HSDA 42: Central Vancouver Island, HSDA 43: North Vancouver Island, HSDA 51: Northwest, HSDA 52: Northern Interior, HSDA 53: Northeast. | | | | | | | | | | | | | | | | | | | | | | |

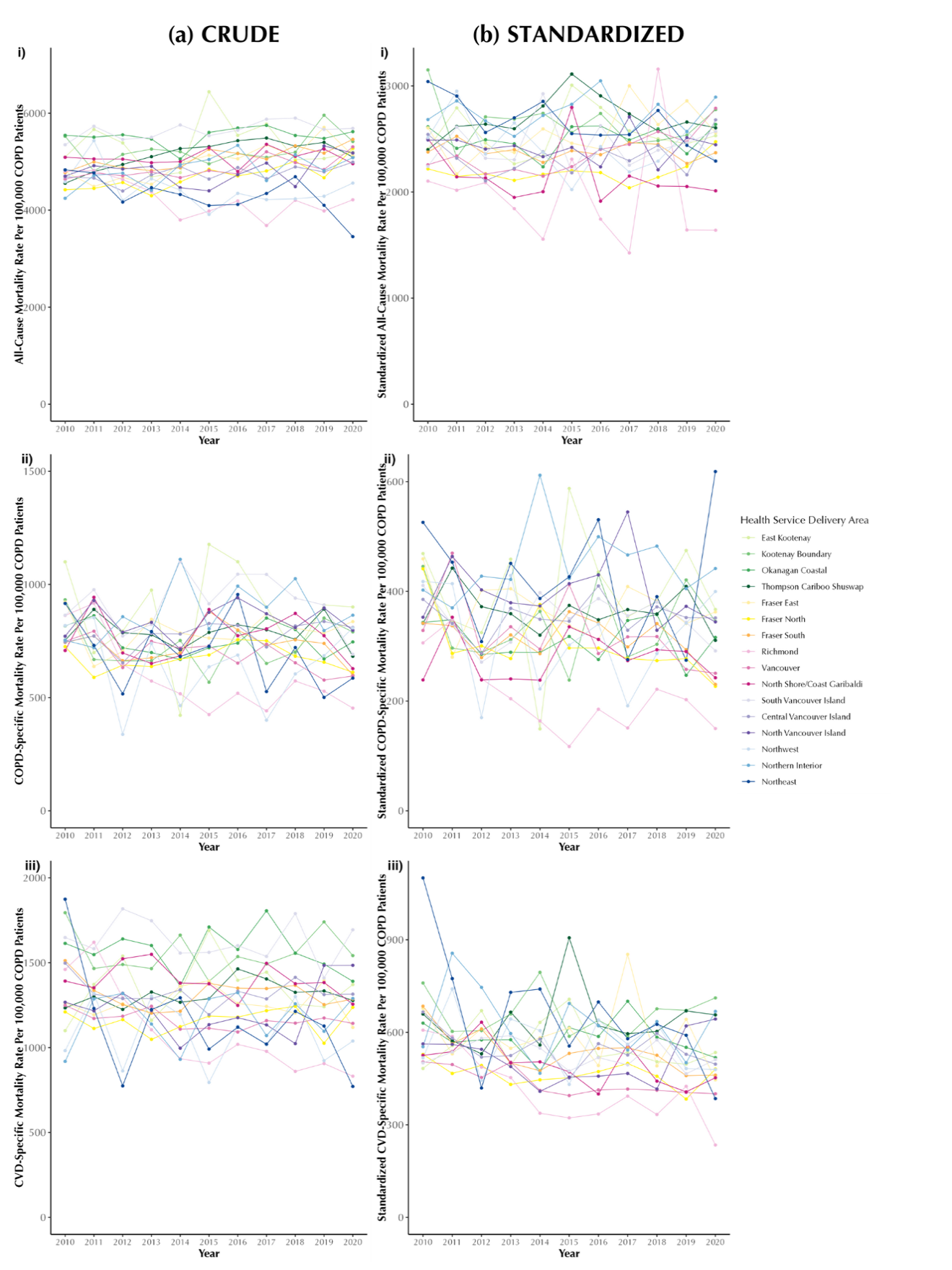
**All-cause Mortality**

**COPD-specific Mortality**

**CVD-specific Mortality**

### **Figure A5: Annual crude (a) and age- and sex-standardized (b) all-cause (i), COPD-specific (ii), and CVD-specific (iii) mortality rates (per 100,000 persons with COPD) in British Columbia, Canada, 2001–2020, stratified by Health Service Delivery Area**.

Rates are expressed as mortality per 100,000 persons with COPD per year (equivalent to per 100,000 COPD patient-years

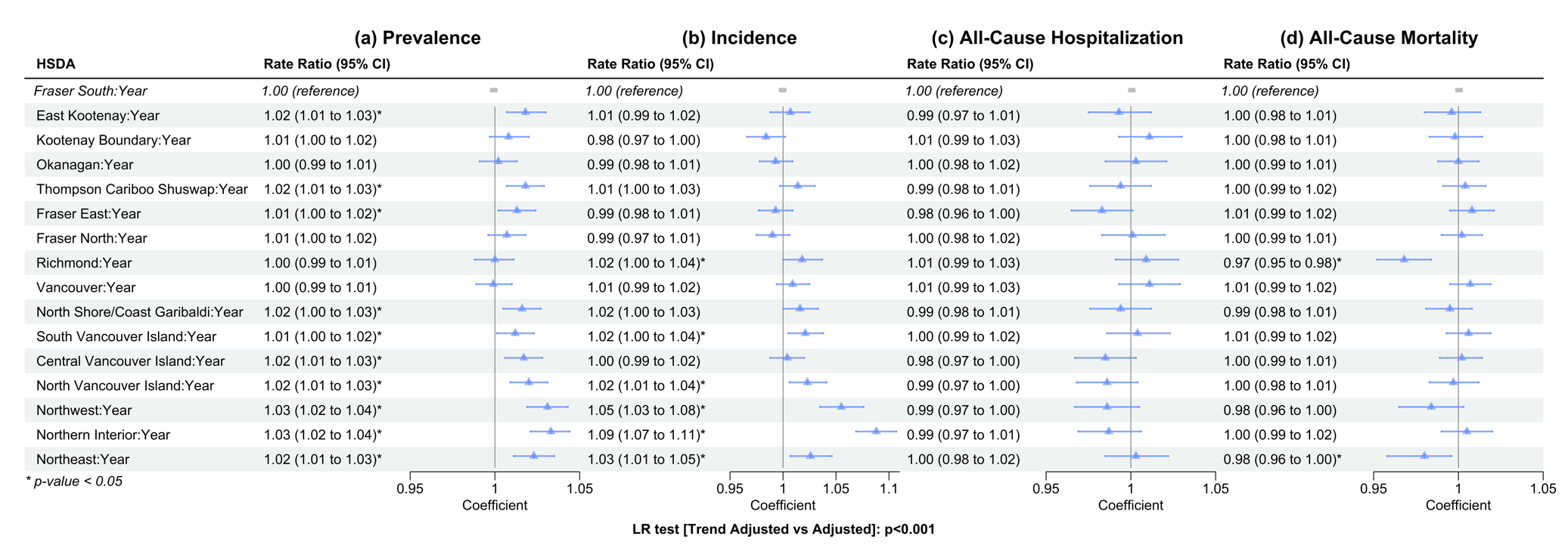

### **Figure A6: Rate ratios presented in a forest plot for the trend adjusted models for each primary outcome Prevalence (a), Incidence (b), All-cause hospitalization (c) and All-cause mortality (d). Confidence bands correspond to 95% CI.**

Reference levels for HSDA: HSDA23 (Fraser South). Adjusted variables: Sex, Socio-Economic Status, Area of Residence, Age group.

Abbreviations: LR test: Likelihood-Ratio test; HSDA: Health Service Delivery Area, CI: Confidence Interval. Adjusted variables: Sex, Socio-Economic Status, Area of Residence, Age group. Abbreviations: LR test: Likelihood-Ratio test; HSDA: Health Service Delivery Area, CI: Confidence Interval

### **Table A10: Average Length of Stay and In-hospital Mortality (COPD specific admission) from 2001 to 2020, stratified by Health Service Delivery Area**

| Admission Year | | | | | | | | | | | | | | | | | | | | | | |
| --- | --- | --- | --- | --- | --- | --- | --- | --- | --- | --- | --- | --- | --- | --- | --- | --- | --- | --- | --- | --- | --- | --- |
|  | **Average Length of Stay (COPD specific admission)** | | | | | | | | | | | **In-Hospital Mortality Rate (per 100) (COPD specific admission)** | | | | | | | | | | |
|  | **2010** | **2011** | **2012** | **2013** | **2014** | **2015** | **2016** | **2017** | **2018** | **2019** | **2020** | **2010** | **2011** | **2012** | **2013** | **2014** | **2015** | **2016** | **2017** | **2018** | **2019** | **2020** |
| Total | 10.1 | 10.2 | 10.1 | 9.7 | 9.7 | 9.2 | 8.8 | 8.8 | 9.0 | 8.6 | 7.9 | 9.9 | 9.5 | 9.3 | 8.4 | 8.9 | 8.4 | 8.6 | 8.2 | 8.1 | 7.6 | 8.1 |
| HSDA11 | 10.7 | 9.0 | 8.4 | 8.0 | 7.1 | 7.8 | 7.4 | 7.5 | 6.1 | 6.8 | 6.5 | 15.5 | 4.7 | 9.4 | 5.8 | 4.2 | 11.4 | 9.5 | 8.2 | 6.2 | 7.1 | 9.4 |
| HSDA12 | 8.4 | 7.7 | 8.1 | 8.3 | 7.5 | 7.9 | 7.3 | 6.8 | 7.3 | 7.4 | 7.3 | 12.7 | 7.2 | 6.3 | 6.3 | 6.1 | 6.8 | 9.4 | 8.0 | 7.0 | 8.8 | 7.7 |
| HSDA13 | 8.6 | 8.7 | 9.0 | 8.8 | 8.8 | 7.8 | 8.0 | 8.1 | 8.3 | 7.7 | 6.9 | 9.8 | 9.5 | 7.9 | 8.2 | 8.5 | 6.9 | 8.5 | 8.7 | 7.8 | 7.5 | 8.7 |
| HSDA14 | 8.2 | 8.9 | 8.4 | 8.5 | 9.3 | 8.8 | 9.0 | 8.4 | 8.2 | 8.2 | 7.4 | 9.6 | 8.9 | 11.0 | 7.9 | 9.2 | 7.3 | 11.8 | 9.1 | 9.4 | 7.3 | 7.3 |
| HSDA21 | 9.4 | 9.3 | 9.4 | 9.2 | 9.5 | 8.6 | 8.4 | 8.7 | 8.1 | 8.6 | 7.3 | 9.2 | 7.9 | 7.3 | 9.3 | 8.0 | 8.5 | 6.6 | 8.3 | 9.4 | 9.2 | 8.8 |
| HSDA22 | 11.5 | 11.8 | 12.2 | 11.7 | 12.1 | 10.2 | 9.8 | 8.9 | 10.0 | 9.2 | 8.6 | 9.5 | 9.5 | 12.2 | 9.9 | 9.0 | 9.6 | 9.1 | 8.5 | 9.6 | 7.5 | 10.6 |
| HSDA23 | 11.8 | 13.0 | 12.0 | 11.2 | 10.8 | 10.0 | 9.3 | 9.1 | 9.1 | 8.5 | 8.5 | 11.6 | 11.8 | 9.6 | 8.6 | 9.7 | 9.3 | 7.7 | 7.9 | 7.3 | 7.4 | 7.5 |
| HSDA31 | 9.5 | 9.0 | 9.8 | 10.4 | 9.0 | 7.8 | 8.4 | 8.2 | 8.7 | 8.5 | 8.1 | 13.4 | 10.5 | 10.5 | 5.9 | 7.8 | 8.5 | 9.1 | 6.3 | 8.2 | 8.7 | 6.8 |
| HSDA32 | 10.1 | 10.2 | 9.9 | 8.5 | 9.2 | 9.2 | 8.2 | 8.4 | 8.5 | 8.8 | 8.3 | 8.5 | 7.4 | 8.3 | 8.0 | 7.1 | 7.9 | 8.1 | 7.3 | 6.6 | 7.4 | 7.1 |
| HSDA33 | 9.5 | 9.9 | 9.0 | 9.1 | 9.3 | 8.8 | 7.9 | 8.5 | 8.9 | 7.9 | 7.1 | 8.6 | 11.5 | 10.9 | 6.9 | 8.1 | 10.6 | 8.8 | 10.3 | 8.9 | 7.7 | 9.7 |
| HSDA41 | 10.8 | 10.6 | 10.9 | 10.4 | 9.0 | 11.3 | 9.6 | 10.2 | 11.2 | 10.0 | 7.8 | 9.7 | 8.6 | 9.5 | 8.0 | 9.7 | 7.8 | 8.2 | 9.0 | 10.1 | 8.2 | 6.4 |
| HSDA42 | 10.7 | 10.8 | 10.4 | 9.8 | 8.9 | 9.0 | 8.7 | 9.5 | 8.3 | 8.7 | 8.8 | 11.0 | 13.6 | 9.2 | 9.9 | 10.8 | 8.7 | 7.7 | 7.9 | 7.9 | 7.8 | 8.1 |
| HSDA43 | 9.5 | 8.8 | 9.4 | 9.5 | 8.8 | 9.5 | 9.1 | 8.5 | 13.3 | 12.6 | 8.4 | 8.3 | 10.0 | 9.2 | 10.3 | 9.3 | 7.1 | 8.6 | 7.8 | 9.0 | 7.1 | 6.7 |
| HSDA51 | 8.7 | 8.0 | 8.6 | 7.6 | 9.0 | 7.7 | 8.1 | 8.6 | 8.4 | 10.2 | 7.9 | 6.3 | 11.3 | 7.1 | 6.2 | 7.4 | 7.0 | 3.7 | 5.0 | 6.7 | 9.7 | 8.7 |
| HSDA52 | 10.1 | 9.2 | 10.1 | 9.7 | 10.2 | 8.6 | 8.8 | 10.1 | 8.6 | 7.4 | 7.2 | 6.9 | 8.1 | 7.8 | 8.8 | 14.1 | 7.8 | 12.2 | 7.6 | 7.1 | 6.7 | 9.1 |
| HSDA 53 | 11.4 | 9.8 | 11.7 | 10.4 | 13.5 | 8.9 | 9.8 | 9.1 | 11.1 | 7.3 | 9.4 | 11.1 | 7.5 | 7.5 | 7.3 | 9.9 | 6.5 | 8.2 | 3.6 | 6.7 | 7.1 | 2.9 |
| Abbreviations: COPD: Chronic Obstructive Pulmonary Disease; HSDA11: East Kootenay, HSDA 12: Kootenay Boundary, HSDA 13: Okanagan, HSDA 14: Thompson Cariboo Shuswap, HSDA 21: Fraser East, HSDA 22: Fraser North, HSDA 23: Fraser South, HSDA 31: Richmond, HSDA 32: Vancouver, HSDA 33: North Shore/Coast Garibaldi, HSDA 41: South Vancouver Island, HSDA 42: Central Vancouver Island, HSDA 43: North Vancouver Island, HSDA 51: Northwest, HSDA 52: Northern Interior, HSDA 53: Northeast. | | | | | | | | | | | | | | | | | | | | | | |

### **Table A11: Readmission Rate (28-day and 90-day) in COPD specific hospitalizations from 2001 to 2020, stratified by Health Service Delivery Area**

| **Admit Year** | **Readmission rates (COPD specific readmission within 28 and 90 days)** | | | | | | | | | | | | | | | | | | | | | |
| --- | --- | --- | --- | --- | --- | --- | --- | --- | --- | --- | --- | --- | --- | --- | --- | --- | --- | --- | --- | --- | --- | --- |
|  | **28-day Hospital Readmission Rate (%)** | | | | | | | | | | | **90-day Hospital Readmission Rate (%)** | | | | | | | | | | |
|  | **2010** | **2011** | **2012** | **2013** | **2014** | **2015** | **2016** | **2017** | **2018** | **2019** | **2020** | **2010** | **2011** | **2012** | **2013** | **2014** | **2015** | **2016** | **2017** | **2018** | **2019** | **2020** |
| **Total** | 10.9 | 12.1 | 12.3 | 11.7 | 13.2 | 12.8 | 12.4 | 13.0 | 12.2 | 12.3 | 12.6 | 18.5 | 21.4 | 21.9 | 21.4 | 22.4 | 23.1 | 22.3 | 23.2 | 22.4 | 21.9 | 23.6 |
| **HSDA11** | 14.0 | 10.4 | 10.2 | 12.9 | 14.4 | 14.3 | 14.3 | 11.6 | 13.6 | 16.9 | 14.9 | 18.5 | 17.9 | 22.3 | 26.3 | 26.3 | 23.8 | 23.0 | 21.6 | 20.9 | 26.7 | 24.9 |
| **HSDA12** | 16.0 | 9.2 | 11.3 | 12.6 | 7.6 | 13.1 | 9.3 | 12.1 | 11.6 | 10.6 | 10.9 | 24.5 | 17.4 | 19.0 | 19.4 | 17.2 | 18.6 | 21.5 | 19.2 | 17.8 | 19.0 | 23.5 |
| **HSDA13** | 12.0 | 13.4 | 11.4 | 10.0 | 9.9 | 11.3 | 10.6 | 11.7 | 13.3 | 11.8 | 13.3 | 19.1 | 21.4 | 20.8 | 17.6 | 18.6 | 19.5 | 19.8 | 23.0 | 22.4 | 22.8 | 24.2 |
| **HSDA14** | 9.1 | 12.6 | 12.5 | 8.9 | 9.1 | 9.2 | 12.4 | 13.6 | 7.7 | 12.4 | 13.6 | 16.5 | 25.0 | 22.8 | 19.9 | 19.7 | 20.4 | 21.4 | 22.9 | 15.8 | 20.1 | 27.2 |
| **HSDA21** | 10.1 | 12.1 | 13.8 | 14.1 | 16.5 | 15.2 | 13.9 | 13.5 | 12.7 | 12.4 | 15.7 | 19.0 | 19.8 | 24.4 | 23.6 | 26.6 | 26.4 | 23.9 | 23.9 | 23.8 | 21.6 | 23.1 |
| **HSDA22** | 9.2 | 11.7 | 10.3 | 12.7 | 11.3 | 13.5 | 12.0 | 12.7 | 11.1 | 11.4 | 13.1 | 18.2 | 22.0 | 20.5 | 22.5 | 20.6 | 23.8 | 22.8 | 24.2 | 22.3 | 19.6 | 25.5 |
| **HSDA23** | 10.4 | 11.1 | 12.8 | 12.9 | 14.6 | 14.8 | 14.3 | 14.0 | 14.7 | 15.3 | 13.5 | 16.9 | 19.6 | 22.6 | 24.1 | 24.3 | 27.3 | 25.2 | 25.3 | 27.7 | 26.3 | 24.0 |
| **HSDA31** | 9.8 | 7.9 | 15.2 | 6.5 | 8.8 | 11.2 | 12.2 | 18.3 | 17.6 | 13.5 | 4.2 | 14.4 | 15.7 | 22.5 | 15.6 | 17.1 | 22.8 | 22.4 | 26.7 | 26.7 | 22.1 | 12.7 |
| **HSDA32** | 14.7 | 13.7 | 11.9 | 13.6 | 19.5 | 13.3 | 9.4 | 14.1 | 13.5 | 13.9 | 15.0 | 23.2 | 26.4 | 22.5 | 25.0 | 29.5 | 25.4 | 20.6 | 24.3 | 26.7 | 24.7 | 26.2 |
| **HSDA33** | 10.3 | 10.4 | 13.2 | 13.7 | 15.0 | 9.7 | 13.7 | 15.5 | 11.1 | 10.0 | 12.0 | 19.0 | 19.5 | 23.1 | 22.5 | 22.2 | 18.6 | 21.8 | 22.8 | 17.9 | 21.0 | 19.5 |
| **HSDA41** | 8.4 | 11.0 | 9.0 | 9.4 | 11.4 | 14.1 | 13.4 | 11.2 | 11.6 | 10.0 | 8.2 | 14.5 | 18.7 | 18.0 | 17.6 | 21.6 | 23.2 | 24.6 | 20.5 | 18.5 | 18.4 | 18.3 |
| **HSDA42** | 10.5 | 14.3 | 11.2 | 9.6 | 12.1 | 10.0 | 13.2 | 12.8 | 12.8 | 12.2 | 11.5 | 19.3 | 23.4 | 20.0 | 17.6 | 19.0 | 20.5 | 20.1 | 21.8 | 25.1 | 22.5 | 23.8 |
| **HSDA43** | 7.8 | 10.6 | 14.2 | 11.0 | 12.7 | 10.2 | 8.2 | 8.5 | 7.2 | 10.3 | 13.3 | 15.2 | 18.6 | 21.9 | 18.3 | 19.4 | 18.6 | 18.1 | 18.9 | 16.8 | 19.1 | 25.2 |
| **HSDA51** | 10.8 | 18.5 | 20.1 | 12.3 | 11.8 | 17.5 | 9.7 | 11.9 | 11.8 | 11.0 | 11.1 | 17.1 | 28.0 | 26.0 | 22.9 | 17.6 | 26.3 | 14.2 | 18.9 | 21.0 | 18.1 | 23.8 |
| **HSDA52** | 11.7 | 12.1 | 15.9 | 12.6 | 15.1 | 12.4 | 14.4 | 11.5 | 8.0 | 8.6 | 8.8 | 17.7 | 20.2 | 25.4 | 24.1 | 23.5 | 21.7 | 22.6 | 21.2 | 16.7 | 16.9 | 22.4 |
| **HSDA53** | 8.1 | 7.5 | 17.0 | 11.8 | 11.4 | 14.6 | 17.5 | 15.0 | 16.0 | 9.2 | 8.1 | 16.3 | 16.8 | 25.5 | 19.1 | 17.4 | 21.1 | 28.7 | 25.9 | 24.2 | 20.7 | 16.2 |
| Abbreviations: COPD: Chronic Obstructive Pulmonary Disease; HSDA11: East Kootenay, HSDA 12: Kootenay Boundary, HSDA 13: Okanagan, HSDA 14: Thompson Cariboo Shuswap, HSDA 21: Fraser East, HSDA 22: Fraser North, HSDA 23: Fraser South, HSDA 31: Richmond, HSDA 32: Vancouver, HSDA 33: North Shore/Coast Garibaldi, HSDA 41: South Vancouver Island, HSDA 42: Central Vancouver Island, HSDA 43: North Vancouver Island, HSDA 51: Northwest, HSDA 52: Northern Interior, HSDA 53: Northeast. | | | | | | | | | | | | | | | | | | | | | | |

### **Table A12: Treatment Classifications for COPD-related Therapies**

| **Treatment Category** | **Full name of Treatment Category** | **DINPIN** |
| --- | --- | --- |
| ICS | Inhaled Glucocorticoid Steroid | 374407, 828521, 828548, 851752, 851760, 852074, 872334, 893633, 897353, 1949993, 1950002, 1978918, 1978926, 2079976, 2174731, 2174758, 2174766, 2174774, 2213583, 2213591, 2213605, 2213613, 2213710, 2213729, 2215039, 2215047, 2215055, 2216531, 2229099, 2237244, 2237245, 2237246, 2237247, 2242029, 2242030, 2244291, 2244292, 2244293, 2285606, 2285614, 2303671, 2243595, 2243596, 2438690, 2446561, 2446588, 2467895, 2467909, 2467917, 897353, 2494272, 2494280, 2465949, 2465957, 2285592, 2510987, 2526557, 2528428, 2503158, 2503166, 2503174, 2503115, 2503123, 2503131. |
| LABA | Long-acting beta agonists | 2136139, 2136147, 2211742, 2214261, 2230898, 2231129, 2237224, 2237225, 2376938, 2407868. |
| ICS+LABA | Inhaled corticosteroid and Long-acting beta agonists | 2240835, 2240836, 2240837, 2245126, 2245127, 2245385, 2245386, 2408872, 2444186, 2361744, 2361752, 2361760, 2248218, 2245217, 2474611, 2474638, 2474646, 2494507, 2494515, 2494523, 2495597, 2495600, 2495619, 2498685, 2498693, 2498707 |
| LAMA | Long-acting muscarinic antagonist | 2246793, 2435381, 2423596, 2394936, 2409720 |
| LAMA+LABA | Long-acting beta agonists and long-acting muscarinic antagonists | 2418401, 2418282, 2441888, 2439530 |
| ICS+LAMA+LABA | Inhaled corticosteroid, long-acting muscarinic antagonist and long-acting beta agonists | 2474522, 2515776, 2501244 |

### **Table A13: Long-acting medication use in COPD by geographic level, aged 35 or older, BC, 2010 to 2020 (combined)**

| HSDA | Total Person-year with diagnosed COPD | Medication | Proportion of long-acting medication users |
| --- | --- | --- | --- |
| Total | 2,113,706 | 893070 | 42.2 |
| Fraser South | 261731 | 115446 | 44.1 |
| Fraser North | 239058 | 97402 | 40.7 |
| Vancouver | 223772 | 87288 | 39 |
| South Vancouver Island | 160403 | 71097 | 44.3 |
| Okanagan | 253414 | 100763 | 39.8 |
| Central Vancouver Island | 183789 | 75783 | 41.2 |
| North Shore-Coast Garibaldi | 111322 | 49310 | 44.3 |
| Fraser East | 144733 | 63537 | 43.9 |
| Thompson Cariboo Shuswap | 159642 | 66354 | 41.6 |
| Richmond | 55324 | 21883 | 39.6 |
| North Vancouver Island | 76759 | 34879 | 45.4 |
| Northern Interior | 74078 | 35878 | 48.4 |
| Kootenay Boundary | 55019 | 23334 | 42.4 |
| East Kootenay | 49386 | 20177 | 40.9 |
| Northwest | 33263 | 15782 | 47.4 |
| Northeast | 32013 | 13286 | 41.5 |
| Abbreviations: COPD: Chronic Obstructive Pulmonary Disease; HSDA: Health Service Delivery Area  HSDAs are sorted based on the population in 2020  Long-Acting Medication contains: ICS, LAMA, LABA, ICS+LABA, LAMA+LABA, LAMA+LABA+ICS | | | |

### **Table A14: Number and proportion of long-acting medication users (%, among COPD patients)**

| **Medication Year** | | | | | | | | | | | | | | | | | | | | | | | |
| --- | --- | --- | --- | --- | --- | --- | --- | --- | --- | --- | --- | --- | --- | --- | --- | --- | --- | --- | --- | --- | --- | --- | --- |
|  | **Number of Users** | | | | | | | | | | | | **Proportion of users (crude)** | | | | | | | | | | |
|  | **2010** | **2011** | **2012** | **2013** | **2014** | **2015** | **2016** | **2017** | **2018** | **2019** | **2020** | **2010** | | **2011** | **2012** | **2013** | **2014** | **2015** | **2016** | **2017** | **2018** | **2019** | **2020** |
| **Total** | 77276 | 78751 | 78486 | 79536 | 80764 | 83022 | 83051 | 83190 | 82857 | 82970 | 83167 | 47.8 | | 45.8 | 43.8 | 42.8 | 42.2 | 42.2 | 41.5 | 40.7 | 40.0 | 39.7 | 39.7 |
| **HSDA11** | 1595 | 1638 | 1636 | 1685 | 1775 | 1867 | 1917 | 1931 | 1992 | 2035 | 2106 | 42.8 | | 40.9 | 39.3 | 39.1 | 39.4 | 40.0 | 40.6 | 40.4 | 41.7 | 42.1 | 43.1 |
| **HSDA12** | 1976 | 2013 | 2073 | 2091 | 2109 | 2151 | 2174 | 2169 | 2208 | 2170 | 2200 | 46.0 | | 43.4 | 42.9 | 42.0 | 41.7 | 42.1 | 42.3 | 41.5 | 41.9 | 41.0 | 42.4 |
| **HSDA13** | 8878 | 8990 | 8892 | 8967 | 9120 | 9334 | 9285 | 9315 | 9283 | 9215 | 9484 | 44.5 | | 42.8 | 40.5 | 39.7 | 39.9 | 40.0 | 39.1 | 38.5 | 37.9 | 37.5 | 38.4 |
| **HSDA14** | 5615 | 5772 | 5832 | 5986 | 6037 | 6128 | 6088 | 6125 | 6189 | 6249 | 6333 | 46.5 | | 44.7 | 43.3 | 42.7 | 42.0 | 42.0 | 40.7 | 39.8 | 39.1 | 38.9 | 39.7 |
| **HSDA21** | 5620 | 5732 | 5706 | 5641 | 5759 | 5897 | 5838 | 5832 | 5798 | 5874 | 5840 | 55.1 | | 52.2 | 47.9 | 45.0 | 44.3 | 43.7 | 42.0 | 40.3 | 39.6 | 39.8 | 39.4 |
| **HSDA22** | 8435 | 8618 | 8576 | 8536 | 8777 | 9092 | 9118 | 9185 | 9026 | 9038 | 9001 | 46.0 | | 43.8 | 41.8 | 40.3 | 40.1 | 40.4 | 39.8 | 39.7 | 39.3 | 39.3 | 39.4 |
| **HSDA23** | 10093 | 10162 | 10137 | 10280 | 10457 | 10720 | 10715 | 10790 | 10695 | 10757 | 10640 | 50.0 | | 47.9 | 46.0 | 45.1 | 44.2 | 43.7 | 42.8 | 42.4 | 41.7 | 41.9 | 41.7 |
| **HSDA31** | 1961 | 2005 | 1952 | 1933 | 1975 | 1995 | 2053 | 2019 | 2012 | 2016 | 1962 | 43.4 | | 42.8 | 40.7 | 39.6 | 39.3 | 38.6 | 39.5 | 38.7 | 38.5 | 38.1 | 37.1 |
| **HSDA32** | 7846 | 7976 | 7944 | 7994 | 8008 | 8147 | 8000 | 8039 | 7880 | 7798 | 7656 | 43.2 | | 41.9 | 40.2 | 39.6 | 38.6 | 38.3 | 37.8 | 38.2 | 37.9 | 37.5 | 36.8 |
| **HSDA33** | 4204 | 4361 | 4294 | 4462 | 4464 | 4576 | 4554 | 4617 | 4636 | 4588 | 4554 | 51.3 | | 49.6 | 46.1 | 46.1 | 43.7 | 42.8 | 42.4 | 42.7 | 42.5 | 41.8 | 41.4 |
| **HSDA41** | 6439 | 6580 | 6420 | 6394 | 6368 | 6646 | 6568 | 6585 | 6433 | 6319 | 6345 | 51.0 | | 49.8 | 47.2 | 45.8 | 44.7 | 45.7 | 44.0 | 42.7 | 41.1 | 39.1 | 39.5 |
| **HSDA42** | 6419 | 6569 | 6551 | 6750 | 6856 | 7010 | 7159 | 7062 | 7095 | 7130 | 7182 | 47.8 | | 45.7 | 43.3 | 42.6 | 42.2 | 42.0 | 41.3 | 39.0 | 37.9 | 37.7 | 37.8 |
| **HSDA43** | 2911 | 3005 | 3026 | 3077 | 3141 | 3321 | 3342 | 3254 | 3247 | 3290 | 3265 | 53.5 | | 50.8 | 48.7 | 47.5 | 46.7 | 47.7 | 46.2 | 42.9 | 41.0 | 40.4 | 40.1 |
| **HSDA51** | 1255 | 1259 | 1283 | 1369 | 1481 | 1529 | 1528 | 1507 | 1497 | 1522 | 1552 | 51.4 | | 48.9 | 48.1 | 48.2 | 49.1 | 48.6 | 47.8 | 46.4 | 45.3 | 45.3 | 44.8 |
| **HSDA52** | 2871 | 2918 | 2976 | 3140 | 3221 | 3300 | 3350 | 3383 | 3507 | 3563 | 3649 | 52.8 | | 50.1 | 49.1 | 49.7 | 48.4 | 48.3 | 48.2 | 48.3 | 47.9 | 46.5 | 45.7 |
| **HSDA53** | 1069 | 1069 | 1104 | 1149 | 1136 | 1235 | 1277 | 1297 | 1282 | 1339 | 1329 | 44.5 | | 41.1 | 40.7 | 41.4 | 38.7 | 40.8 | 42.1 | 42.7 | 42.0 | 41.9 | 41.0 |

Abbreviations: COPD: Chronic Obstructive Pulmonary Disease; HSDA11: East Kootenay, HSDA 12: Kootenay Boundary, HSDA 13: Okanagan, HSDA 14: Thompson Cariboo Shuswap, HSDA 21: Fraser East, HSDA 22: Fraser North, HSDA 23: Fraser South, HSDA 31: Richmond, HSDA 32: Vancouver, HSDA 33: North Shore/Coast Garibaldi, HSDA 41: South Vancouver Island, HSDA 42: Central Vancouver Island, HSDA 43: North Vancouver Island, HSDA 51: Northwest, HSDA 52: Northern Interior, HSDA 53: Northeast.

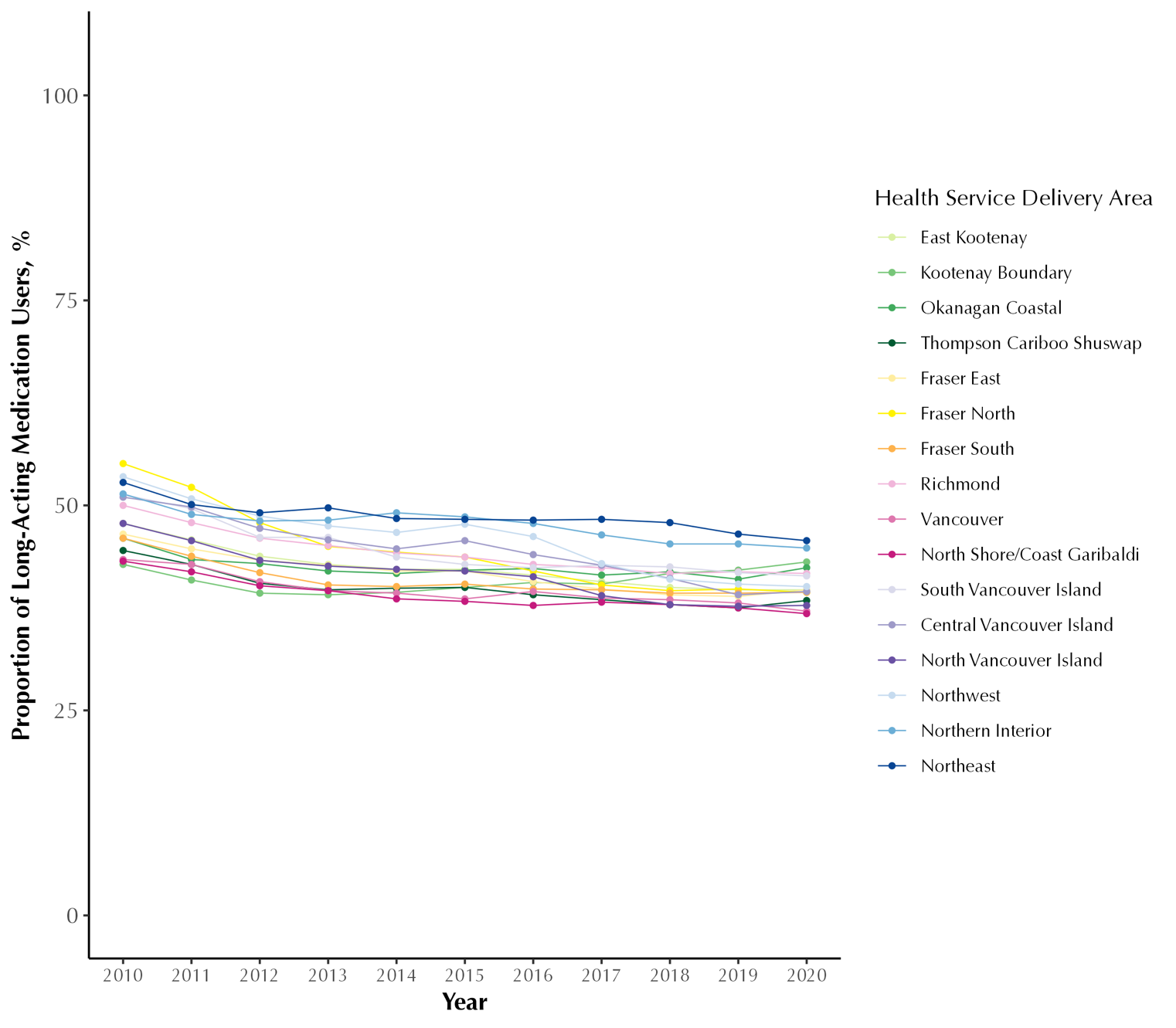

### **Figure A7: Proportion of long-acting medication users (%, among COPD patients) in British Columbia, Canada, from 2001 to 2020, stratified by Health Service Delivery Area**

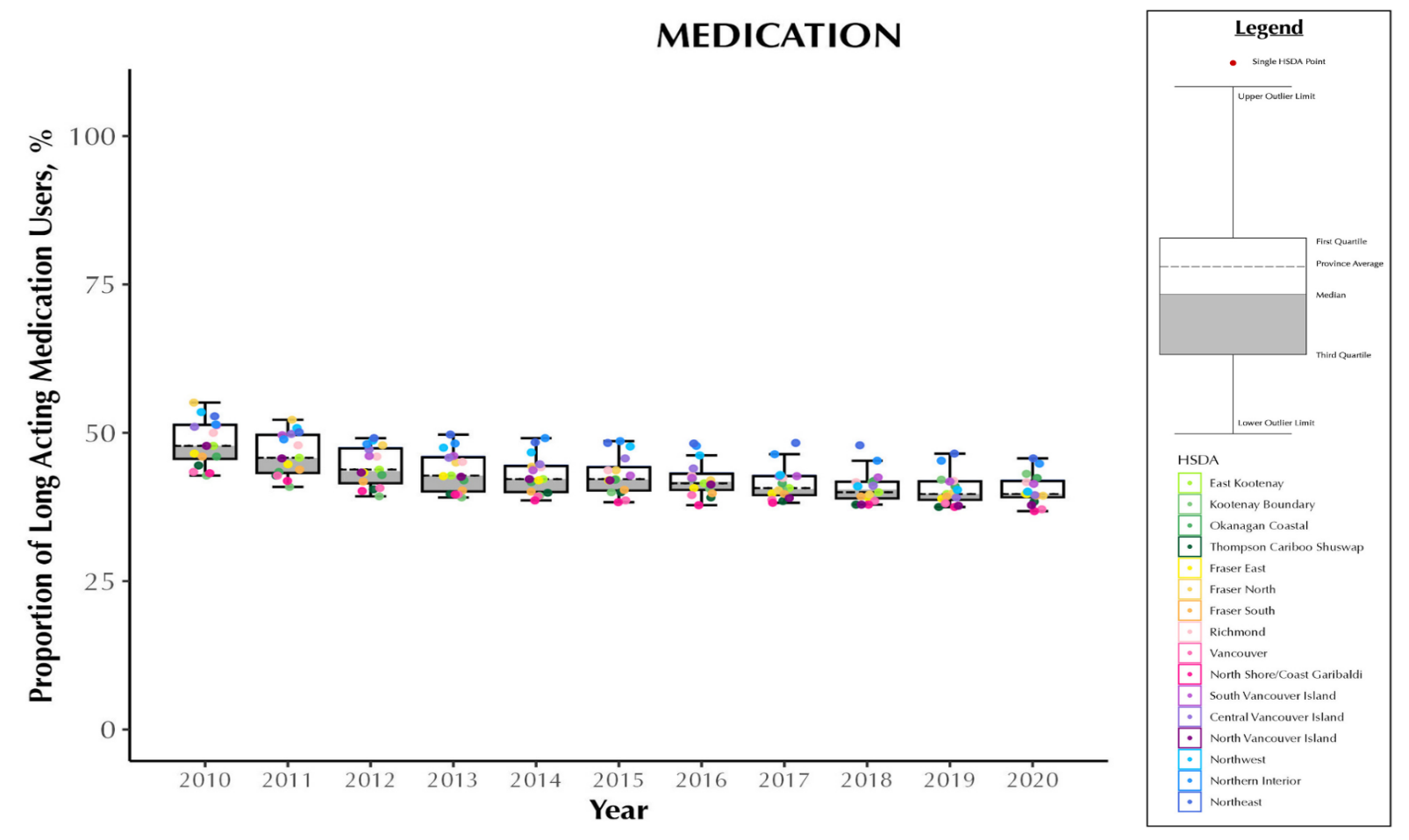

### **Figure A8: Proportion of long-acting medication users (%) in British Columbia, Canada, from 2001 to 2020, stratified by Health Service Delivery Area**

Abbreviations: COPD: chronic obstructive pulmonary disease; Q1: quartile 1; Q3: quartile 3 Note: Each dot represents an HSDA. The horizontal line cutting through the plot is the overall provincial average. The median is the line separating the upper (white) and lower (dark grey) boxes

### **Table A15: Rate ratios and 95% confidence interval for the unadjusted, adjusted and trend adjusted models for medication use (long acting)**

|  | **Unadjusted Model** | |  |  | **Adjusted Model** | |  |  | **Trend Adjusted Model** | | | |
| --- | --- | --- | --- | --- | --- | --- | --- | --- | --- | --- | --- | --- |
| **Term** | **p value** | **medication rate ratio** | **95% CI (lower)** | **95% CI (upper)** | **p value** | **medication rate ratio** | **95% CI (lower)** | **95% CI (upper)** | **p value** | **Medication rate ratio** | **95% CI (lower)** | **95% CI (upper)** |
| **(Intercept)** | 0.000 | 0.495 | 0.479 | 0.511 | 0.000 | 0.366 | 0.337 | 0.398 | 0.000 | 0.353 | 0.327 | 0.381 |
| **HSDA11** | 0.350 | 1.024 | 0.975 | 1.075 | 0.233 | 1.044 | 0.973 | 1.120 | 0.068 | 0.927 | 0.854 | 1.006 |
| **HSDA12** | 0.115 | 1.040 | 0.991 | 1.092 | 0.000 | 1.121 | 1.056 | 1.190 | 0.336 | 1.036 | 0.964 | 1.113 |
| **HSDA13** | 0.096 | 1.040 | 0.993 | 1.090 | 0.004 | 1.064 | 1.020 | 1.111 | 0.360 | 1.027 | 0.970 | 1.086 |
| **HSDA14** | 0.004 | 1.072 | 1.023 | 1.123 | 0.004 | 1.093 | 1.029 | 1.161 | 0.123 | 1.055 | 0.986 | 1.129 |
| **HSDA21** | 0.259 | 1.027 | 0.980 | 1.077 | 0.000 | 1.109 | 1.068 | 1.151 | 0.000 | 1.185 | 1.123 | 1.250 |
| **HSDA22** | 0.049 | 0.954 | 0.911 | 1.000 | 0.099 | 0.971 | 0.937 | 1.006 | 0.129 | 0.961 | 0.912 | 1.012 |
| **HSDA31** | 0.233 | 0.971 | 0.925 | 1.019 | 0.122 | 1.032 | 0.992 | 1.073 | 0.822 | 0.993 | 0.938 | 1.052 |
| **HSDA32** | 0.056 | 0.956 | 0.912 | 1.001 | 0.858 | 1.004 | 0.962 | 1.048 | 0.412 | 0.977 | 0.924 | 1.033 |
| **HSDA33** | 0.049 | 1.049 | 1.000 | 1.099 | 0.057 | 0.940 | 0.883 | 1.002 | 0.056 | 0.933 | 0.869 | 1.002 |
| **HSDA41** | 0.001 | 1.082 | 1.032 | 1.134 | 0.000 | 1.090 | 1.051 | 1.130 | 0.000 | 1.123 | 1.065 | 1.185 |
| **HSDA42** | 0.711 | 1.009 | 0.963 | 1.057 | 0.755 | 1.007 | 0.965 | 1.050 | 0.213 | 1.037 | 0.980 | 1.097 |
| **HSDA43** | 0.000 | 1.093 | 1.042 | 1.147 | 0.000 | 1.139 | 1.089 | 1.190 | 0.000 | 1.190 | 1.121 | 1.263 |
| **HSDA51** | 0.000 | 1.134 | 1.079 | 1.191 | 0.000 | 1.201 | 1.122 | 1.286 | 0.000 | 1.160 | 1.076 | 1.250 |
| **HSDA52** | 0.000 | 1.154 | 1.100 | 1.210 | 0.000 | 1.215 | 1.163 | 1.269 | 0.000 | 1.183 | 1.116 | 1.254 |
| **HSDA53** | 0.571 | 1.014 | 0.966 | 1.065 | 0.151 | 0.946 | 0.876 | 1.021 | 0.000 | 0.854 | 0.785 | 0.929 |
| **HSDA23 (Ref)** |  |  |  |  |  |  |  |  |  |  |  |  |
| **Female** |  |  |  |  | 0.000 | 1.095 | 1.080 | 1.110 | 0.000 | 1.103 | 1.090 | 1.116 |
| **Male (Ref)** |  |  |  |  |  |  |  |  |  |  |  |  |
| **50-64 years** |  |  |  |  | 0.000 | 0.752 | 0.737 | 0.769 | 0.000 | 0.751 | 0.738 | 0.765 |
| **65-79 years** |  |  |  |  | 0.000 | 0.776 | 0.759 | 0.793 | 0.000 | 0.771 | 0.756 | 0.786 |
| **>=80 years** |  |  |  |  | 0.000 | 0.721 | 0.705 | 0.737 | 0.000 | 0.718 | 0.705 | 0.732 |
| **35-49 years (Ref)** | |  |  |  |  |  |  |  |  |  |  |  |
| **Rural** |  |  |  |  | 0.370 | 0.889 | 0.688 | 1.150 | 0.502 | 1.081 | 0.861 | 1.356 |
| **Neighborhood income quintile** | | | | | | | | | | | | |
| **2** |  |  |  |  | 0.000 | 2.783 | 1.987 | 3.897 | 0.000 | 4.013 | 2.992 | 5.383 |
| **3** |  |  |  |  | 0.000 | 2.293 | 1.588 | 3.313 | 0.000 | 3.097 | 2.250 | 4.265 |
| **4** |  |  |  |  | 0.000 | 2.964 | 2.072 | 4.241 | 0.000 | 3.494 | 2.547 | 4.793 |
| **5 (highest income quintile)** |  |  |  |  | 0.000 | 4.276 | 3.008 | 6.078 | 0.000 | 4.790 | 3.527 | 6.505 |
| **Unknown** |  |  |  |  | 0.028 | 4.296 | 1.170 | 15.783 | 0.003 | 5.489 | 1.770 | 17.022 |
| **Year** |  |  |  |  |  |  |  |  | 0.000 | 0.981 | 0.975 | 0.987 |
| **HSDA11:year** |  |  |  |  |  |  |  |  | 0.005 | 1.014 | 1.004 | 1.024 |
| **HSDA12:year** |  |  |  |  |  |  |  |  | 0.048 | 1.010 | 1.000 | 1.019 |
| **HSDA13:year** |  |  |  |  |  |  |  |  | 0.448 | 1.003 | 0.995 | 1.012 |
| **HSDA14:year** |  |  |  |  |  |  |  |  | 0.793 | 0.999 | 0.990 | 1.008 |
| **HSDA21:year** |  |  |  |  |  |  |  |  | 0.004 | 0.987 | 0.978 | 0.996 |
| **HSDA22:year** |  |  |  |  |  |  |  |  | 0.657 | 1.002 | 0.993 | 1.011 |
| **HSDA31:year** |  |  |  |  |  |  |  |  | 0.077 | 1.009 | 0.999 | 1.018 |
| **HSDA32:year** |  |  |  |  |  |  |  |  | 0.082 | 1.008 | 0.999 | 1.016 |
| **HSDA33:year** |  |  |  |  |  |  |  |  | 0.611 | 0.998 | 0.989 | 1.007 |
| **HSDA41:year** |  |  |  |  |  |  |  |  | 0.220 | 0.995 | 0.986 | 1.003 |
| **HSDA42:year** |  |  |  |  |  |  |  |  | 0.060 | 0.992 | 0.983 | 1.000 |
| **HSDA43:year** |  |  |  |  |  |  |  |  | 0.006 | 0.987 | 0.978 | 0.996 |
| **HSDA51:year** |  |  |  |  |  |  |  |  | 0.829 | 1.001 | 0.991 | 1.011 |
| **HSDA52:year** |  |  |  |  |  |  |  |  | 0.543 | 1.003 | 0.994 | 1.012 |
| **HSDA53:year** |  |  |  |  |  |  |  |  | 0.002 | 1.016 | 1.006 | 1.026 |
| **HSDA23: Year (Ref)** | | | | | | | | | | | | |
| Reference levels of factor variables have a rate ratio=1.00.  Adjusted variables for both adjusted and trend-adjusted models: Sex, Socio-Economic Status, Area of Residence, Age group.  Abbreviations: HSDA: Health Service Delivery Area, CI: Confidence Interval, HSDA11: East Kootenay, HSDA 12: Kootenay Boundary, HSDA 13: Okanagan, HSDA 14: Thompson Cariboo Shuswap, HSDA 21: Fraser East, HSDA 22: Fraser North, HSDA 23: Fraser South, HSDA 31: Richmond, HSDA 32: Vancouver, HSDA 33: North Shore/Coast Garibaldi, HSDA 41: South Vancouver Island, HSDA 42: Central Vancouver Island, HSDA 43: North Vancouver Island, HSDA 51: Northwest, HSDA 52: Northern Interior, HSDA 53: Northeast. | | | | | | | | | | | | |

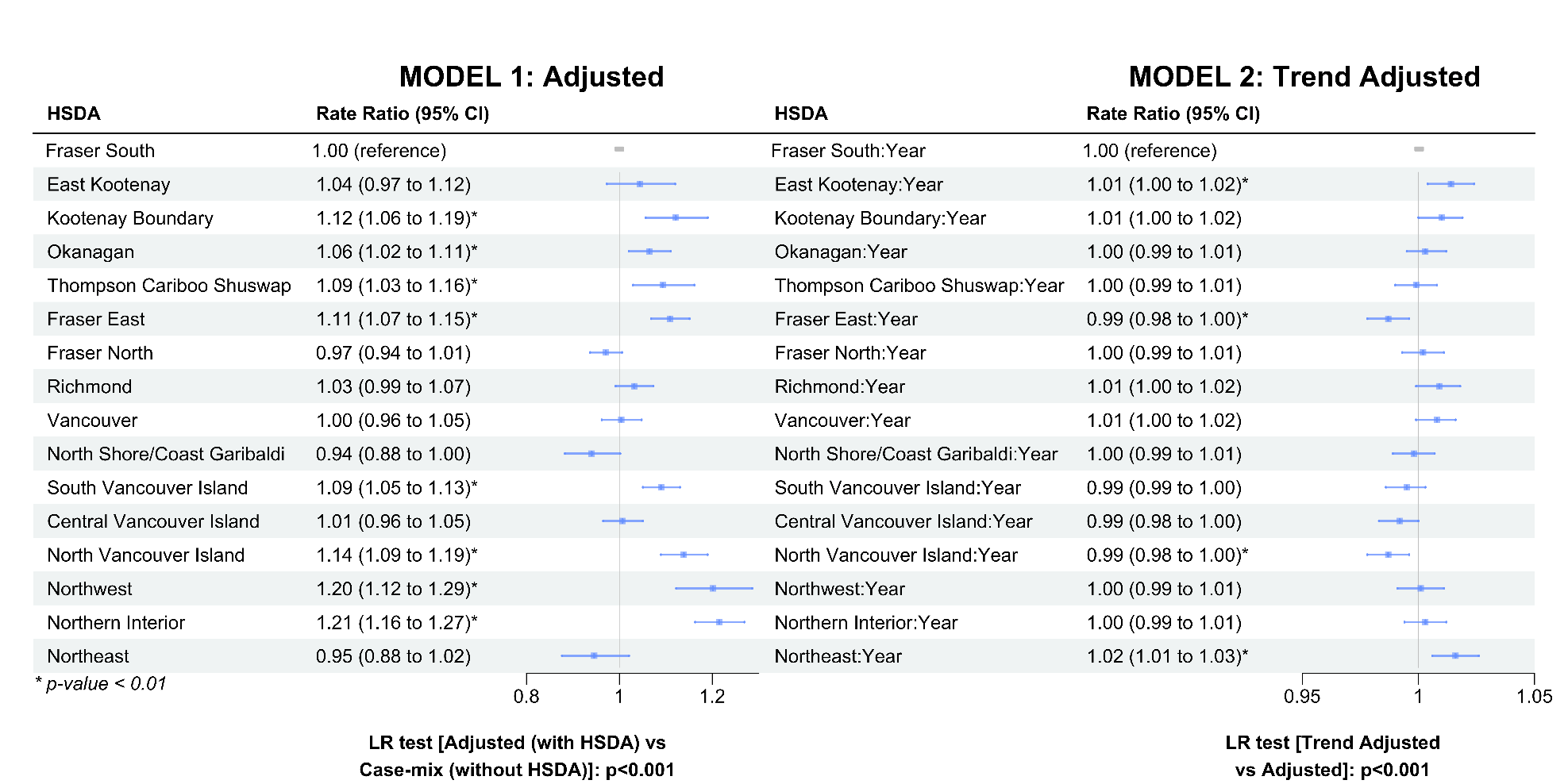

**Figure A9: Rate ratios presented in a stacked forest plot for the adjusted (a) and trend adjusted (b) models for number of long-acting medication users.**

Confidence bands correspond to 95% CI. HSDA11: East Kootenay, HSDA 12: Kootenay Boundary, HSDA 13: Okanagan, HSDA 14: Thompson Cariboo Shuswap, HSDA 21: Fraser East, HSDA 22: Fraser North, HSDA 23: Fraser South, HSDA 31: Richmond, HSDA 32: Vancouver, HSDA 33: North Shore/Coast Garibaldi, HSDA 41: South Vancouver Island, HSDA 42: Central Vancouver Island, HSDA 43: North Vancouver Island, HSDA 51: Northwest, HSDA 52: Northern Interior, HSDA 53: Northeast. Reference levels for HSDA: HSDA23 (Fraser South). Reference levels for HSDA: HSDA23 (Fraser South). Adjusted variables: Sex, Socio-Economic Status, Area of Residence, Age group. Abbreviations: LR test: Likelihood-Ratio test; HSDA: Health Service Delivery Area, CI: Confidence Interval. Adjusted variables: Sex, Socio-Economic Status, Area of Residence, Age group. Abbreviations: LR test: Likelihood-Ratio test; HSDA: Health Service Delivery Area, CI: Confidence Interval

### **Table A16: Number of Long-acting medication users by drug class**

|  | **Medication Year** | | | | | | | | | | | | | | | | | | | | | |
| --- | --- | --- | --- | --- | --- | --- | --- | --- | --- | --- | --- | --- | --- | --- | --- | --- | --- | --- | --- | --- | --- | --- |
|  | **2010** | **2011** | **2012** | **2013** | **2014** | **2015** | **2016** | **2017** | **2018** | **2019** | **2020** | **2010** | **2011** | **2012** | **2013** | **2014** | **2015** | **2016** | **2017** | **2018** | **2019** | **2020** |
|  | **COMBINATION THERAPY** | | | | | | | | | | | **SINGLE THERAPY** | | | | | | | | | | |
|  | **LAMA+LABA** | | | | | | | | | | | **LAMA** | | | | | | | | | | |
| **Total** | 498 | 468 | 465 | 524 | 950 | 2078 | 4778 | 7119 | 9076 | 10148 | 10560 | 12089 | 13451 | 14204 | 15163 | 16526 | 17909 | 18507 | 18330 | 18628 | 18070 | 18726 |
| **HSDA11** | 12 | 8 | 7 | 8 | 15 | 26 | 84 | 144 | 271 | 359 | 390 | 227 | 292 | 291 | 313 | 345 | 395 | 428 | 495 | 566 | 567 | 553 |
| **HSDA12** | 36 | 30 | 25 | 31 | 26 | 43 | 177 | 259 | 328 | 389 | 416 | 471 | 497 | 533 | 534 | 534 | 566 | 610 | 536 | 583 | 533 | 570 |
| **HSDA13** | 156 | 146 | 150 | 145 | 172 | 340 | 718 | 1058 | 1311 | 1562 | 1678 | 2041 | 2207 | 2229 | 2363 | 2545 | 2738 | 2748 | 2556 | 2557 | 2446 | 2510 |
| **HSDA14** | 37 | 39 | 39 | 36 | 46 | 89 | 193 | 303 | 397 | 504 | 550 | 904 | 992 | 1045 | 1095 | 1132 | 1211 | 1291 | 1287 | 1331 | 1355 | 1501 |
| **HSDA21** | 27 | 23 | 26 | 37 | 82 | 173 | 332 | 486 | 603 | 661 | 667 | 721 | 764 | 800 | 862 | 962 | 1036 | 1029 | 1007 | 1117 | 1135 | 1141 |
| **HSDA22** | 29 | 25 | 28 | 32 | 119 | 293 | 588 | 851 | 1081 | 1133 | 1093 | 1264 | 1465 | 1580 | 1696 | 1878 | 2036 | 2104 | 2125 | 2064 | 1952 | 1937 |
| **HSDA23** | 40 | 36 | 42 | 58 | 139 | 345 | 720 | 998 | 1255 | 1343 | 1403 | 1292 | 1470 | 1584 | 1705 | 2048 | 2208 | 2269 | 2292 | 2341 | 2272 | 2311 |
| **HSDA31** | 11 | 15 | 8 | 8 | 28 | 53 | 146 | 173 | 203 | 223 | 226 | 304 | 327 | 332 | 369 | 391 | 432 | 447 | 389 | 416 | 406 | 391 |
| **HSDA32** | 27 | 30 | 30 | 43 | 106 | 234 | 556 | 755 | 905 | 992 | 1021 | 910 | 1108 | 1232 | 1393 | 1543 | 1642 | 1690 | 1645 | 1642 | 1569 | 1638 |
| **HSDA33** | 27 | 23 | 23 | 22 | 44 | 86 | 201 | 336 | 417 | 459 | 484 | 633 | 757 | 771 | 875 | 961 | 1012 | 1091 | 1159 | 1135 | 1109 | 1155 |
| **HSDA41** | 32 | 34 | 30 | 38 | 87 | 171 | 433 | 675 | 837 | 882 | 905 | 1187 | 1230 | 1294 | 1354 | 1407 | 1548 | 1509 | 1508 | 1489 | 1427 | 1418 |
| **HSDA42** | 33 | 35 | 29 | 32 | 40 | 128 | 363 | 615 | 770 | 825 | 836 | 1139 | 1244 | 1310 | 1329 | 1417 | 1524 | 1570 | 1521 | 1503 | 1420 | 1514 |
| **HSDA43** | 6 | <5 | 12 | 11 | 21 | 38 | 113 | 194 | 261 | 290 | 322 | 455 | 505 | 520 | 543 | 573 | 702 | 750 | 772 | 816 | 836 | 886 |
| **HSDA51** | <5 | <5 | <5 | 6 | 7 | 10 | 27 | 61 | 111 | 125 | 124 | 111 | 114 | 144 | 160 | 169 | 202 | 218 | 258 | 265 | 253 | 306 |
| **HSDA52** | 12 | 9 | <5 | 8 | 12 | 34 | 90 | 144 | 238 | 305 | 334 | 303 | 321 | 368 | 399 | 434 | 464 | 510 | 521 | 543 | 529 | 617 |
| **HSDA53** | 10 | 10 | 7 | 7 | 6 | 14 | 34 | 60 | 84 | 91 | 104 | 112 | 142 | 153 | 149 | 168 | 174 | 220 | 239 | 241 | 248 | 263 |
|  | **ICS+LAMA** | | | | | | | | | | | **LABA** | | | | | | | | | | |
| **Total** | 1434 | 1546 | 1528 | 1563 | 1546 | 1618 | 1621 | 1531 | 1448 | 1481 | 1637 | 2205 | 2006 | 1831 | 1766 | 1909 | 1884 | 1480 | 1343 | 1227 | 1134 | 984 |
| **HSDA11** | 23 | 28 | 20 | 24 | 25 | 15 | 22 | 28 | 29 | 35 | 35 | 60 | 45 | 48 | 45 | 41 | 54 | 45 | 48 | 44 | 34 | 30 |
| **HSDA12** | 79 | 69 | 73 | 78 | 65 | 77 | 60 | 50 | 49 | 35 | 47 | 112 | 109 | 87 | 89 | 84 | 78 | 66 | 67 | 63 | 58 | 47 |
| **HSDA13** | 256 | 251 | 229 | 243 | 227 | 216 | 203 | 174 | 163 | 151 | 188 | 500 | 473 | 437 | 394 | 395 | 392 | 274 | 265 | 231 | 203 | 171 |
| **HSDA14** | 139 | 146 | 152 | 138 | 131 | 120 | 128 | 130 | 122 | 130 | 150 | 185 | 171 | 167 | 156 | 156 | 155 | 109 | 106 | 99 | 90 | 87 |
| **HSDA21** | 96 | 111 | 101 | 117 | 89 | 113 | 106 | 95 | 91 | 112 | 99 | 131 | 115 | 113 | 131 | 151 | 151 | 128 | 100 | 85 | 72 | 70 |
| **HSDA22** | 127 | 138 | 153 | 137 | 145 | 164 | 166 | 154 | 136 | 132 | 161 | 159 | 133 | 122 | 141 | 157 | 170 | 97 | 91 | 84 | 88 | 64 |
| **HSDA23** | 132 | 182 | 200 | 208 | 215 | 251 | 237 | 234 | 229 | 249 | 249 | 198 | 184 | 174 | 161 | 184 | 181 | 140 | 115 | 111 | 90 | 73 |
| **HSDA31** | 32 | 26 | 31 | 30 | 26 | 39 | 52 | 41 | 36 | 34 | 40 | 43 | 37 | 35 | 40 | 40 | 46 | 34 | 21 | 18 | 20 | 19 |
| **HSDA32** | 109 | 104 | 109 | 106 | 129 | 135 | 130 | 122 | 124 | 124 | 135 | 150 | 146 | 140 | 136 | 168 | 149 | 116 | 107 | 117 | 115 | 92 |
| **HSDA33** | 58 | 71 | 82 | 95 | 95 | 100 | 92 | 86 | 88 | 91 | 85 | 130 | 126 | 100 | 98 | 112 | 100 | 72 | 68 | 59 | 56 | 46 |
| **HSDA41** | 113 | 106 | 88 | 90 | 96 | 92 | 100 | 111 | 97 | 103 | 109 | 189 | 170 | 151 | 133 | 154 | 152 | 148 | 127 | 115 | 105 | 101 |
| **HSDA42** | 124 | 150 | 131 | 128 | 134 | 144 | 140 | 135 | 122 | 107 | 140 | 154 | 126 | 108 | 99 | 107 | 107 | 99 | 87 | 67 | 79 | 68 |
| **HSDA43** | 54 | 58 | 52 | 68 | 68 | 62 | 73 | 71 | 69 | 77 | 74 | 57 | 51 | 47 | 44 | 63 | 57 | 70 | 69 | 63 | 55 | 49 |
| **HSDA51** | 17 | 19 | 18 | 22 | 20 | 27 | 29 | 24 | 21 | 21 | 29 | 21 | 15 | 17 | 16 | 23 | 14 | 13 | 16 | 18 | 16 | 15 |
| **HSDA52** | 51 | 53 | 60 | 52 | 57 | 44 | 50 | 44 | 44 | 49 | 63 | 63 | 56 | 47 | 47 | 43 | 43 | 39 | 36 | 39 | 42 | 41 |
| **HSDA53** | 22 | 32 | 26 | 25 | 23 | 18 | 31 | 30 | 27 | 31 | 31 | 49 | 46 | 36 | 34 | 28 | 33 | 29 | 19 | 14 | 10 | 11 |
|  | **ICS+LABA** | | | | | | | | | | | **ICS** | | | | | | | | | | |
| **Total** | 44622 | 45829 | 46608 | 47738 | 48496 | 49765 | 48911 | 47140 | 45105 | 43388 | 42751 | 32480 | 31839 | 30069 | 29479 | 28640 | 28769 | 27581 | 26590 | 25392 | 24253 | 22335 |
| **HSDA11** | 1031 | 1108 | 1110 | 1165 | 1241 | 1319 | 1302 | 1272 | 1179 | 1104 | 1090 | 520 | 491 | 454 | 439 | 459 | 438 | 458 | 405 | 426 | 454 | 427 |
| **HSDA12** | 996 | 1080 | 1159 | 1216 | 1237 | 1247 | 1208 | 1101 | 1111 | 1063 | 1010 | 868 | 841 | 820 | 778 | 747 | 745 | 702 | 666 | 614 | 542 | 505 |
| **HSDA13** | 5086 | 5251 | 5294 | 5353 | 5406 | 5614 | 5397 | 5171 | 4852 | 4554 | 4601 | 3379 | 3173 | 2983 | 2877 | 2735 | 2719 | 2560 | 2519 | 2418 | 2252 | 2227 |
| **HSDA14** | 2808 | 3002 | 3067 | 3286 | 3317 | 3427 | 3414 | 3414 | 3334 | 3278 | 3311 | 2738 | 2678 | 2609 | 2547 | 2467 | 2434 | 2262 | 2206 | 2147 | 2014 | 1831 |
| **HSDA21** | 3044 | 3096 | 3215 | 3185 | 3211 | 3332 | 3297 | 3186 | 3092 | 3025 | 2973 | 2660 | 2701 | 2525 | 2417 | 2390 | 2377 | 2280 | 2196 | 2054 | 1968 | 1835 |
| **HSDA22** | 5215 | 5310 | 5425 | 5388 | 5533 | 5600 | 5573 | 5341 | 5019 | 4822 | 4724 | 3277 | 3167 | 2915 | 2824 | 2796 | 2887 | 2714 | 2685 | 2502 | 2422 | 2263 |
| **HSDA23** | 5944 | 5933 | 6048 | 6184 | 6284 | 6393 | 6244 | 6020 | 5703 | 5547 | 5455 | 4352 | 4286 | 4060 | 4069 | 3914 | 3903 | 3761 | 3651 | 3471 | 3383 | 3062 |
| **HSDA31** | 1221 | 1278 | 1229 | 1214 | 1266 | 1266 | 1252 | 1186 | 1123 | 1106 | 1115 | 724 | 744 | 682 | 639 | 644 | 619 | 655 | 643 | 610 | 580 | 515 |
| **HSDA32** | 4640 | 4694 | 4762 | 4894 | 4865 | 4866 | 4668 | 4544 | 4289 | 4105 | 3930 | 3480 | 3369 | 3200 | 3068 | 2869 | 2893 | 2735 | 2635 | 2530 | 2376 | 2099 |
| **HSDA33** | 2731 | 2841 | 2860 | 2957 | 2954 | 2981 | 2896 | 2826 | 2776 | 2718 | 2680 | 1505 | 1491 | 1384 | 1412 | 1404 | 1403 | 1349 | 1318 | 1318 | 1231 | 1128 |
| **HSDA41** | 4032 | 4069 | 4079 | 4069 | 4057 | 4222 | 4096 | 3842 | 3588 | 3442 | 3408 | 2244 | 2320 | 2105 | 2005 | 1945 | 2034 | 1945 | 1853 | 1728 | 1630 | 1510 |
| **HSDA42** | 3728 | 3838 | 3889 | 4089 | 4160 | 4262 | 4275 | 3992 | 3892 | 3667 | 3601 | 2587 | 2536 | 2420 | 2437 | 2398 | 2391 | 2373 | 2250 | 2197 | 2127 | 1942 |
| **HSDA43** | 1576 | 1649 | 1676 | 1723 | 1800 | 1909 | 1883 | 1821 | 1744 | 1688 | 1617 | 1316 | 1310 | 1264 | 1262 | 1213 | 1241 | 1226 | 1100 | 1055 | 1002 | 872 |
| **HSDA51** | 630 | 647 | 660 | 721 | 786 | 812 | 837 | 835 | 810 | 746 | 736 | 656 | 648 | 628 | 649 | 668 | 694 | 649 | 604 | 542 | 538 | 495 |
| **HSDA52** | 1366 | 1453 | 1509 | 1645 | 1705 | 1807 | 1871 | 1852 | 1850 | 1793 | 1786 | 1558 | 1514 | 1462 | 1468 | 1461 | 1431 | 1309 | 1272 | 1293 | 1264 | 1200 |
| **HSDA53** | 527 | 534 | 588 | 608 | 629 | 667 | 653 | 687 | 698 | 692 | 670 | 573 | 530 | 522 | 550 | 501 | 532 | 568 | 561 | 466 | 456 | 408 |
|  | **LAMA+LABA+ICS** | | | | | | | | | | |  |  |  |  |  |  |  |  |  |  |  |
| **Total** | 9518 | 10494 | 11299 | 12304 | 12953 | 14020 | 14864 | 15425 | 16379 | 18256 | 19326 |  |  |  |  |  |  |  |  |  |  |  |
| **HSDA11** | 194 | 225 | 231 | 245 | 280 | 307 | 353 | 414 | 456 | 556 | 599 |  |  |  |  |  |  |  |  |  |  |  |
| **HSDA12** | 310 | 345 | 394 | 429 | 421 | 447 | 490 | 503 | 505 | 555 | 554 |  |  |  |  |  |  |  |  |  |  |  |
| **HSDA13** | 1581 | 1691 | 1737 | 1850 | 1936 | 2034 | 2152 | 2201 | 2194 | 2311 | 2498 |  |  |  |  |  |  |  |  |  |  |  |
| **HSDA14** | 660 | 727 | 771 | 860 | 886 | 942 | 997 | 1095 | 1217 | 1340 | 1480 |  |  |  |  |  |  |  |  |  |  |  |
| **HSDA21** | 578 | 593 | 680 | 701 | 741 | 788 | 853 | 886 | 994 | 1198 | 1251 |  |  |  |  |  |  |  |  |  |  |  |
| **HSDA22** | 1107 | 1313 | 1362 | 1500 | 1526 | 1697 | 1749 | 1839 | 1944 | 2128 | 2208 |  |  |  |  |  |  |  |  |  |  |  |
| **HSDA23** | 1161 | 1230 | 1344 | 1480 | 1655 | 1808 | 1971 | 2018 | 2148 | 2388 | 2480 |  |  |  |  |  |  |  |  |  |  |  |
| **HSDA31** | 234 | 244 | 258 | 300 | 302 | 332 | 340 | 321 | 344 | 351 | 370 |  |  |  |  |  |  |  |  |  |  |  |
| **HSDA32** | 815 | 935 | 1073 | 1215 | 1292 | 1416 | 1399 | 1399 | 1483 | 1593 | 1612 |  |  |  |  |  |  |  |  |  |  |  |
| **HSDA33** | 450 | 545 | 596 | 677 | 705 | 750 | 822 | 850 | 889 | 981 | 1039 |  |  |  |  |  |  |  |  |  |  |  |
| **HSDA41** | 824 | 860 | 920 | 983 | 998 | 1069 | 1127 | 1107 | 1120 | 1227 | 1290 |  |  |  |  |  |  |  |  |  |  |  |
| **HSDA42** | 843 | 938 | 1007 | 1032 | 1077 | 1158 | 1173 | 1219 | 1331 | 1557 | 1662 |  |  |  |  |  |  |  |  |  |  |  |
| **HSDA43** | 348 | 397 | 412 | 447 | 496 | 558 | 609 | 642 | 660 | 752 | 806 |  |  |  |  |  |  |  |  |  |  |  |
| **HSDA51** | 91 | 94 | 108 | 122 | 133 | 155 | 178 | 224 | 266 | 323 | 372 |  |  |  |  |  |  |  |  |  |  |  |
| **HSDA52** | 226 | 252 | 285 | 330 | 357 | 402 | 473 | 502 | 570 | 657 | 737 |  |  |  |  |  |  |  |  |  |  |  |
| **HSDA53** | 85 | 95 | 105 | 120 | 138 | 142 | 158 | 190 | 240 | 321 | 353 |  |  |  |  |  |  |  |  |  |  |  |
| Abbreviations: HSDA11: East Kootenay, HSDA 12: Kootenay Boundary, HSDA 13: Okanagan, HSDA 14: Thompson Cariboo Shuswap, HSDA 21: Fraser East, HSDA 22: Fraser North, HSDA 23: Fraser South, HSDA 31: Richmond, HSDA 32: Vancouver, HSDA 33: North Shore/Coast Garibaldi, HSDA 41: South Vancouver Island, HSDA 42: Central Vancouver Island, HSDA 43: North Vancouver Island, HSDA 51: Northwest, HSDA 52: Northern Interior, HSDA 53: Northeast. ICS: Inhaled Corticosteroids; LAMA: Long-Acting Muscarinic Antagonists; LABA: Long-Acting Beta-Agonists | | | | | | | | | | | | | | | | | | | | | | |

### **Table A17: Proportion of Long-acting medication users (%, among COPD patients) by drug class**

|  | **Medication Year** | | | | | | | | | | | | | | | | | | | | | |
| --- | --- | --- | --- | --- | --- | --- | --- | --- | --- | --- | --- | --- | --- | --- | --- | --- | --- | --- | --- | --- | --- | --- |
|  | **2010** | **2011** | **2012** | **2013** | **2014** | **2015** | **2016** | **2017** | **2018** | **2019** | **2020** | **2010** | **2011** | **2012** | **2013** | **2014** | **2015** | **2016** | **2017** | **2018** | **2019** | **2020** |
|  | **COMBINATION THERAPY** | | | | | | | | | | | **SINGLE THERAPY** | | | | | | | | | | |
|  | **LAMA+LABA** | | | | | | | | | | | **LAMA** | | | | | | | | | | |
| **Total** | 0.3 | 0.3 | 0.3 | 0.3 | 0.5 | 1.1 | 2.4 | 3.5 | 4.4 | 4.9 | 5.0 | 7.5 | 7.8 | 7.9 | 8.2 | 8.6 | 9.1 | 9.2 | 9.0 | 9.0 | 8.6 | 8.9 |
| **HSDA11** | 0.3 | 0.2 | 0.2 | 0.2 | 0.3 | 0.6 | 1.8 | 3.0 | 5.7 | 7.4 | 8.0 | 6.1 | 7.3 | 7.0 | 7.3 | 7.7 | 8.5 | 9.1 | 10.4 | 11.8 | 11.7 | 11.3 |
| **HSDA12** | 0.8 | 0.6 | 0.5 | 0.6 | 0.5 | 0.8 | 3.4 | 5.0 | 6.2 | 7.4 | 8.0 | 11.0 | 10.7 | 11.0 | 10.7 | 10.6 | 11.1 | 11.9 | 10.3 | 11.1 | 10.1 | 11.0 |
| **HSDA13** | 0.8 | 0.7 | 0.7 | 0.6 | 0.8 | 1.5 | 3.0 | 4.4 | 5.4 | 6.3 | 6.8 | 10.2 | 10.5 | 10.2 | 10.5 | 11.1 | 11.7 | 11.6 | 10.6 | 10.4 | 9.9 | 10.2 |
| **HSDA14** | 0.3 | 0.3 | 0.3 | 0.3 | 0.3 | 0.6 | 1.3 | 2.0 | 2.5 | 3.1 | 3.4 | 7.5 | 7.7 | 7.8 | 7.8 | 7.9 | 8.3 | 8.6 | 8.4 | 8.4 | 8.4 | 9.4 |
| **HSDA21** | 0.3 | 0.2 | 0.2 | 0.3 | 0.6 | 1.3 | 2.4 | 3.4 | 4.1 | 4.5 | 4.5 | 7.1 | 7.0 | 6.7 | 6.9 | 7.4 | 7.7 | 7.4 | 7.0 | 7.6 | 7.7 | 7.7 |
| **HSDA22** | 0.2 | 0.1 | 0.1 | 0.2 | 0.5 | 1.3 | 2.6 | 3.7 | 4.7 | 4.9 | 4.8 | 6.9 | 7.4 | 7.7 | 8.0 | 8.6 | 9.0 | 9.2 | 9.2 | 9.0 | 8.5 | 8.5 |
| **HSDA23** | 0.2 | 0.2 | 0.2 | 0.3 | 0.6 | 1.4 | 2.9 | 3.9 | 4.9 | 5.2 | 5.5 | 6.4 | 6.9 | 7.2 | 7.5 | 8.7 | 9.0 | 9.1 | 9.0 | 9.1 | 8.8 | 9.1 |
| **HSDA31** | 0.2 | 0.3 | 0.2 | 0.2 | 0.6 | 1.0 | 2.8 | 3.3 | 3.9 | 4.2 | 4.3 | 6.7 | 7.0 | 6.9 | 7.6 | 7.8 | 8.3 | 8.6 | 7.5 | 8.0 | 7.7 | 7.4 |
| **HSDA32** | 0.1 | 0.2 | 0.2 | 0.2 | 0.5 | 1.1 | 2.6 | 3.6 | 4.4 | 4.8 | 4.9 | 5.0 | 5.8 | 6.2 | 6.9 | 7.4 | 7.7 | 8.0 | 7.8 | 7.9 | 7.6 | 7.9 |
| **HSDA33** | 0.3 | 0.3 | 0.2 | 0.2 | 0.4 | 0.8 | 1.9 | 3.1 | 3.8 | 4.2 | 4.4 | 7.7 | 8.6 | 8.3 | 9.0 | 9.4 | 9.5 | 10.2 | 10.7 | 10.4 | 10.1 | 10.5 |
| **HSDA41** | 0.3 | 0.3 | 0.2 | 0.3 | 0.6 | 1.2 | 2.9 | 4.4 | 5.3 | 5.5 | 5.6 | 9.4 | 9.3 | 9.5 | 9.7 | 9.9 | 10.6 | 10.1 | 9.8 | 9.5 | 8.8 | 8.8 |
| **HSDA42** | 0.2 | 0.2 | 0.2 | 0.2 | 0.2 | 0.8 | 2.1 | 3.4 | 4.1 | 4.4 | 4.4 | 8.5 | 8.7 | 8.7 | 8.4 | 8.7 | 9.1 | 9.1 | 8.4 | 8.0 | 7.5 | 8.0 |
| **HSDA43** | 0.1 | 0.1 | 0.2 | 0.2 | 0.3 | 0.5 | 1.6 | 2.6 | 3.3 | 3.6 | 4.0 | 8.4 | 8.5 | 8.4 | 8.4 | 8.5 | 10.1 | 10.4 | 10.2 | 10.3 | 10.3 | 10.9 |
| **HSDA51** | 0.1 | 0.0 | 0.1 | 0.2 | 0.2 | 0.3 | 0.8 | 1.9 | 3.4 | 3.7 | 3.6 | 4.5 | 4.4 | 5.4 | 5.6 | 5.6 | 6.4 | 6.8 | 7.9 | 8.0 | 7.5 | 8.8 |
| **HSDA52** | 0.2 | 0.2 | 0.1 | 0.1 | 0.2 | 0.5 | 1.3 | 2.1 | 3.3 | 4.0 | 4.2 | 5.6 | 5.5 | 6.1 | 6.3 | 6.5 | 6.8 | 7.3 | 7.4 | 7.4 | 6.9 | 7.7 |
| **HSDA53** | 0.4 | 0.4 | 0.3 | 0.3 | 0.2 | 0.5 | 1.1 | 2.0 | 2.8 | 2.8 | 3.2 | 4.7 | 5.5 | 5.6 | 5.4 | 5.7 | 5.7 | 7.2 | 7.9 | 7.9 | 7.8 | 8.1 |
|  | **ICS+LAMA** | | | | | | | | | | | **LABA** | | | | | | | | | | |
| **Total** | 0.9 | 0.9 | 0.9 | 0.8 | 0.8 | 0.8 | 0.8 | 0.7 | 0.7 | 0.7 | 0.8 | 1.4 | 1.2 | 1.0 | 1.0 | 1.0 | 1.0 | 0.7 | 0.7 | 0.6 | 0.5 | 0.5 |
| **HSDA11** | 0.6 | 0.7 | 0.5 | 0.6 | 0.6 | 0.3 | 0.5 | 0.6 | 0.6 | 0.7 | 0.7 | 1.6 | 1.1 | 1.2 | 1.0 | 0.9 | 1.2 | 1.0 | 1.0 | 0.9 | 0.7 | 0.6 |
| **HSDA12** | 1.8 | 1.5 | 1.5 | 1.6 | 1.3 | 1.5 | 1.2 | 1.0 | 0.9 | 0.7 | 0.9 | 2.6 | 2.4 | 1.8 | 1.8 | 1.7 | 1.5 | 1.3 | 1.3 | 1.2 | 1.1 | 0.9 |
| **HSDA13** | 1.3 | 1.2 | 1.0 | 1.1 | 1.0 | 0.9 | 0.9 | 0.7 | 0.7 | 0.6 | 0.8 | 2.5 | 2.3 | 2.0 | 1.7 | 1.7 | 1.7 | 1.2 | 1.1 | 0.9 | 0.8 | 0.7 |
| **HSDA14** | 1.2 | 1.1 | 1.1 | 1.0 | 0.9 | 0.8 | 0.9 | 0.8 | 0.8 | 0.8 | 0.9 | 1.5 | 1.3 | 1.2 | 1.1 | 1.1 | 1.1 | 0.7 | 0.7 | 0.6 | 0.6 | 0.5 |
| **HSDA21** | 0.9 | 1.0 | 0.8 | 0.9 | 0.7 | 0.8 | 0.8 | 0.7 | 0.6 | 0.8 | 0.7 | 1.3 | 1.0 | 0.9 | 1.0 | 1.2 | 1.1 | 0.9 | 0.7 | 0.6 | 0.5 | 0.5 |
| **HSDA22** | 0.7 | 0.7 | 0.7 | 0.6 | 0.7 | 0.7 | 0.7 | 0.7 | 0.6 | 0.6 | 0.7 | 0.9 | 0.7 | 0.6 | 0.7 | 0.7 | 0.8 | 0.4 | 0.4 | 0.4 | 0.4 | 0.3 |
| **HSDA23** | 0.7 | 0.9 | 0.9 | 0.9 | 0.9 | 1.0 | 0.9 | 0.9 | 0.9 | 1.0 | 1.0 | 1.0 | 0.9 | 0.8 | 0.7 | 0.8 | 0.7 | 0.6 | 0.5 | 0.4 | 0.4 | 0.3 |
| **HSDA31** | 0.7 | 0.6 | 0.6 | 0.6 | 0.5 | 0.8 | 1.0 | 0.8 | 0.7 | 0.6 | 0.8 | 1.0 | 0.8 | 0.7 | 0.8 | 0.8 | 0.9 | 0.7 | 0.4 | 0.3 | 0.4 | 0.4 |
| **HSDA32** | 0.6 | 0.5 | 0.6 | 0.5 | 0.6 | 0.6 | 0.6 | 0.6 | 0.6 | 0.6 | 0.6 | 0.8 | 0.8 | 0.7 | 0.7 | 0.8 | 0.7 | 0.5 | 0.5 | 0.6 | 0.6 | 0.4 |
| **HSDA33** | 0.7 | 0.8 | 0.9 | 1.0 | 0.9 | 0.9 | 0.9 | 0.8 | 0.8 | 0.8 | 0.8 | 1.6 | 1.4 | 1.1 | 1.0 | 1.1 | 0.9 | 0.7 | 0.6 | 0.5 | 0.5 | 0.4 |
| **HSDA41** | 0.9 | 0.8 | 0.6 | 0.6 | 0.7 | 0.6 | 0.7 | 0.7 | 0.6 | 0.6 | 0.7 | 1.5 | 1.3 | 1.1 | 1.0 | 1.1 | 1.0 | 1.0 | 0.8 | 0.7 | 0.6 | 0.6 |
| **HSDA42** | 0.9 | 1.0 | 0.9 | 0.8 | 0.8 | 0.9 | 0.8 | 0.7 | 0.7 | 0.6 | 0.7 | 1.1 | 0.9 | 0.7 | 0.6 | 0.7 | 0.6 | 0.6 | 0.5 | 0.4 | 0.4 | 0.4 |
| **HSDA43** | 1.0 | 1.0 | 0.8 | 1.0 | 1.0 | 0.9 | 1.0 | 0.9 | 0.9 | 0.9 | 0.9 | 1.0 | 0.9 | 0.8 | 0.7 | 0.9 | 0.8 | 1.0 | 0.9 | 0.8 | 0.7 | 0.6 |
| **HSDA51** | 0.7 | 0.7 | 0.7 | 0.8 | 0.7 | 0.9 | 0.9 | 0.7 | 0.6 | 0.6 | 0.8 | 0.9 | 0.6 | 0.6 | 0.6 | 0.8 | 0.4 | 0.4 | 0.5 | 0.5 | 0.5 | 0.4 |
| **HSDA52** | 0.9 | 0.9 | 1.0 | 0.8 | 0.9 | 0.6 | 0.7 | 0.6 | 0.6 | 0.6 | 0.8 | 1.2 | 1.0 | 0.8 | 0.7 | 0.6 | 0.6 | 0.6 | 0.5 | 0.5 | 0.5 | 0.5 |
| **HSDA53** | 0.9 | 1.2 | 1.0 | 0.9 | 0.8 | 0.6 | 1.0 | 1.0 | 0.9 | 1.0 | 1.0 | 2.0 | 1.8 | 1.3 | 1.2 | 1.0 | 1.1 | 1.0 | 0.6 | 0.5 | 0.3 | 0.3 |
|  | **ICS+LABA** | | | | | | | | | | | **ICS** | | | | | | | | | | |
| **Total** | 27.6 | 26.7 | 26.0 | 25.7 | 25.4 | 25.3 | 24.4 | 23.1 | 21.8 | 20.7 | 20.4 | 20.1 | 18.5 | 16.8 | 15.9 | 15.0 | 14.6 | 13.8 | 13.0 | 12.3 | 11.6 | 10.7 |
| **HSDA11** | 27.7 | 27.6 | 26.7 | 27.0 | 27.6 | 28.2 | 27.5 | 26.6 | 24.7 | 22.8 | 22.3 | 13.9 | 12.3 | 10.9 | 10.2 | 10.2 | 9.4 | 9.7 | 8.5 | 8.9 | 9.4 | 8.7 |
| **HSDA12** | 23.2 | 23.3 | 24.0 | 24.4 | 24.5 | 24.4 | 23.5 | 21.1 | 21.1 | 20.1 | 19.5 | 20.2 | 18.1 | 17.0 | 15.6 | 14.8 | 14.6 | 13.6 | 12.7 | 11.6 | 10.3 | 9.7 |
| **HSDA13** | 25.5 | 25.0 | 24.1 | 23.7 | 23.6 | 24.1 | 22.7 | 21.4 | 19.8 | 18.5 | 18.7 | 16.9 | 15.1 | 13.6 | 12.7 | 12.0 | 11.7 | 10.8 | 10.4 | 9.9 | 9.2 | 9.0 |
| **HSDA14** | 23.3 | 23.2 | 22.8 | 23.5 | 23.1 | 23.5 | 22.8 | 22.2 | 21.0 | 20.4 | 20.7 | 22.7 | 20.7 | 19.4 | 18.2 | 17.2 | 16.7 | 15.1 | 14.3 | 13.6 | 12.6 | 11.5 |
| **HSDA21** | 29.8 | 28.2 | 27.0 | 25.4 | 24.7 | 24.7 | 23.7 | 22.0 | 21.1 | 20.5 | 20.0 | 26.1 | 24.6 | 21.2 | 19.3 | 18.4 | 17.6 | 16.4 | 15.2 | 14.0 | 13.3 | 12.4 |
| **HSDA22** | 28.4 | 27.0 | 26.4 | 25.4 | 25.3 | 24.9 | 24.4 | 23.1 | 21.8 | 21.0 | 20.7 | 17.9 | 16.1 | 14.2 | 13.3 | 12.8 | 12.8 | 11.9 | 11.6 | 10.9 | 10.5 | 9.9 |
| **HSDA23** | 29.5 | 27.9 | 27.5 | 27.2 | 26.6 | 26.1 | 24.9 | 23.7 | 22.2 | 21.6 | 21.4 | 21.6 | 20.2 | 18.4 | 17.9 | 16.6 | 15.9 | 15.0 | 14.3 | 13.5 | 13.2 | 12.0 |
| **HSDA31** | 27.0 | 27.2 | 25.6 | 24.8 | 25.2 | 24.5 | 24.1 | 22.8 | 21.5 | 20.9 | 21.1 | 16.0 | 15.9 | 14.2 | 13.1 | 12.8 | 12.0 | 12.6 | 12.3 | 11.7 | 10.9 | 9.7 |
| **HSDA32** | 25.5 | 24.7 | 24.1 | 24.2 | 23.5 | 22.9 | 22.1 | 21.6 | 20.6 | 19.8 | 18.9 | 19.2 | 17.7 | 16.2 | 15.2 | 13.8 | 13.6 | 12.9 | 12.5 | 12.2 | 11.4 | 10.1 |
| **HSDA33** | 33.3 | 32.3 | 30.7 | 30.5 | 28.9 | 27.9 | 27.0 | 26.1 | 25.5 | 24.7 | 24.4 | 18.4 | 16.9 | 14.8 | 14.6 | 13.7 | 13.1 | 12.6 | 12.2 | 12.1 | 11.2 | 10.3 |
| **HSDA41** | 32.0 | 30.8 | 30.0 | 29.1 | 28.4 | 29.0 | 27.4 | 24.9 | 22.9 | 21.3 | 21.2 | 17.8 | 17.6 | 15.5 | 14.4 | 13.6 | 14.0 | 13.0 | 12.0 | 11.0 | 10.1 | 9.4 |
| **HSDA42** | 27.8 | 26.7 | 25.7 | 25.8 | 25.6 | 25.6 | 24.7 | 22.0 | 20.8 | 19.4 | 18.9 | 19.3 | 17.6 | 16.0 | 15.4 | 14.7 | 14.3 | 13.7 | 12.4 | 11.7 | 11.3 | 10.2 |
| **HSDA43** | 28.9 | 27.9 | 27.0 | 26.6 | 26.8 | 27.4 | 26.1 | 24.0 | 22.0 | 20.7 | 19.8 | 24.2 | 22.1 | 20.4 | 19.5 | 18.0 | 17.8 | 17.0 | 14.5 | 13.3 | 12.3 | 10.7 |
| **HSDA51** | 25.8 | 25.1 | 24.7 | 25.4 | 26.1 | 25.8 | 26.2 | 25.7 | 24.5 | 22.2 | 21.2 | 26.8 | 25.1 | 23.5 | 22.9 | 22.2 | 22.1 | 20.3 | 18.6 | 16.4 | 16.0 | 14.3 |
| **HSDA52** | 25.1 | 24.9 | 24.9 | 26.0 | 25.6 | 26.4 | 26.9 | 26.4 | 25.3 | 23.4 | 22.4 | 28.6 | 26.0 | 24.1 | 23.2 | 21.9 | 20.9 | 18.8 | 18.2 | 17.7 | 16.5 | 15.0 |
| **HSDA53** | 21.9 | 20.5 | 21.7 | 21.9 | 21.4 | 22.0 | 21.5 | 22.6 | 22.9 | 21.7 | 20.7 | 23.9 | 20.4 | 19.3 | 19.8 | 17.1 | 17.6 | 18.7 | 18.5 | 15.3 | 14.3 | 12.6 |
|  | **LAMA+LABA+ICS** | | | | | | | | | | |  |  |  |  |  |  |  |  |  |  |  |
| **Total** | 5.9 | 6.1 | 6.3 | 6.6 | 6.8 | 7.1 | 7.4 | 7.6 | 7.9 | 8.7 | 9.2 |  |  |  |  |  |  |  |  |  |  |  |
| **HSDA11** | 5.2 | 5.6 | 5.6 | 5.7 | 6.2 | 6.6 | 7.5 | 8.7 | 9.5 | 11.5 | 12.3 |  |  |  |  |  |  |  |  |  |  |  |
| **HSDA12** | 7.2 | 7.4 | 8.2 | 8.6 | 8.3 | 8.8 | 9.5 | 9.6 | 9.6 | 10.5 | 10.7 |  |  |  |  |  |  |  |  |  |  |  |
| **HSDA13** | 7.9 | 8.1 | 7.9 | 8.2 | 8.5 | 8.7 | 9.1 | 9.1 | 9.0 | 9.4 | 10.1 |  |  |  |  |  |  |  |  |  |  |  |
| **HSDA14** | 5.5 | 5.6 | 5.7 | 6.1 | 6.2 | 6.5 | 6.7 | 7.1 | 7.7 | 8.4 | 9.3 |  |  |  |  |  |  |  |  |  |  |  |
| **HSDA21** | 5.7 | 5.4 | 5.7 | 5.6 | 5.7 | 5.8 | 6.1 | 6.1 | 6.8 | 8.1 | 8.4 |  |  |  |  |  |  |  |  |  |  |  |
| **HSDA22** | 6.0 | 6.7 | 6.6 | 7.1 | 7.0 | 7.5 | 7.6 | 7.9 | 8.5 | 9.3 | 9.7 |  |  |  |  |  |  |  |  |  |  |  |
| **HSDA23** | 5.8 | 5.8 | 6.1 | 6.5 | 7.0 | 7.4 | 7.9 | 7.9 | 8.4 | 9.3 | 9.7 |  |  |  |  |  |  |  |  |  |  |  |
| **HSDA31** | 5.2 | 5.2 | 5.4 | 6.1 | 6.0 | 6.4 | 6.5 | 6.2 | 6.6 | 6.6 | 7.0 |  |  |  |  |  |  |  |  |  |  |  |
| **HSDA32** | 4.5 | 4.9 | 5.4 | 6.0 | 6.2 | 6.7 | 6.6 | 6.6 | 7.1 | 7.7 | 7.7 |  |  |  |  |  |  |  |  |  |  |  |
| **HSDA33** | 5.5 | 6.2 | 6.4 | 7.0 | 6.9 | 7.0 | 7.7 | 7.9 | 8.2 | 8.9 | 9.5 |  |  |  |  |  |  |  |  |  |  |  |
| **HSDA41** | 6.5 | 6.5 | 6.8 | 7.0 | 7.0 | 7.4 | 7.6 | 7.2 | 7.2 | 7.6 | 8.0 |  |  |  |  |  |  |  |  |  |  |  |
| **HSDA42** | 6.3 | 6.5 | 6.7 | 6.5 | 6.6 | 6.9 | 6.8 | 6.7 | 7.1 | 8.2 | 8.7 |  |  |  |  |  |  |  |  |  |  |  |
| **HSDA43** | 6.4 | 6.7 | 6.6 | 6.9 | 7.4 | 8.0 | 8.4 | 8.5 | 8.3 | 9.2 | 9.9 |  |  |  |  |  |  |  |  |  |  |  |
| **HSDA51** | 3.7 | 3.6 | 4.0 | 4.3 | 4.4 | 4.9 | 5.6 | 6.9 | 8.0 | 9.6 | 10.7 |  |  |  |  |  |  |  |  |  |  |  |
| **HSDA52** | 4.2 | 4.3 | 4.7 | 5.2 | 5.4 | 5.9 | 6.8 | 7.2 | 7.8 | 8.6 | 9.2 |  |  |  |  |  |  |  |  |  |  |  |
| **HSDA53** | 3.5 | 3.7 | 3.9 | 4.3 | 4.7 | 4.7 | 5.2 | 6.3 | 7.9 | 10.1 | 10.9 |  |  |  |  |  |  |  |  |  |  |  |
| Abbreviations: HSDA: Health Service Delivery Area; HSDA11: East Kootenay, HSDA 12: Kootenay Boundary, HSDA 13: Okanagan, HSDA 14: Thompson Cariboo Shuswap, HSDA 21: Fraser East, HSDA 22: Fraser North, HSDA 23: Fraser South, HSDA 31: Richmond, HSDA 32: Vancouver, HSDA 33: North Shore/Coast Garibaldi, HSDA 41: South Vancouver Island, HSDA 42: Central Vancouver Island, HSDA 43: North Vancouver Island, HSDA 51: Northwest, HSDA 52: Northern Interior, HSDA 53: Northeast. ICS: Inhaled Corticosteroids; LAMA: Long-Acting Muscarinic Antagonists; LABA: Long-Acting Beta-Agonists | | | | | | | | | | | | | | | | | | | | | | |

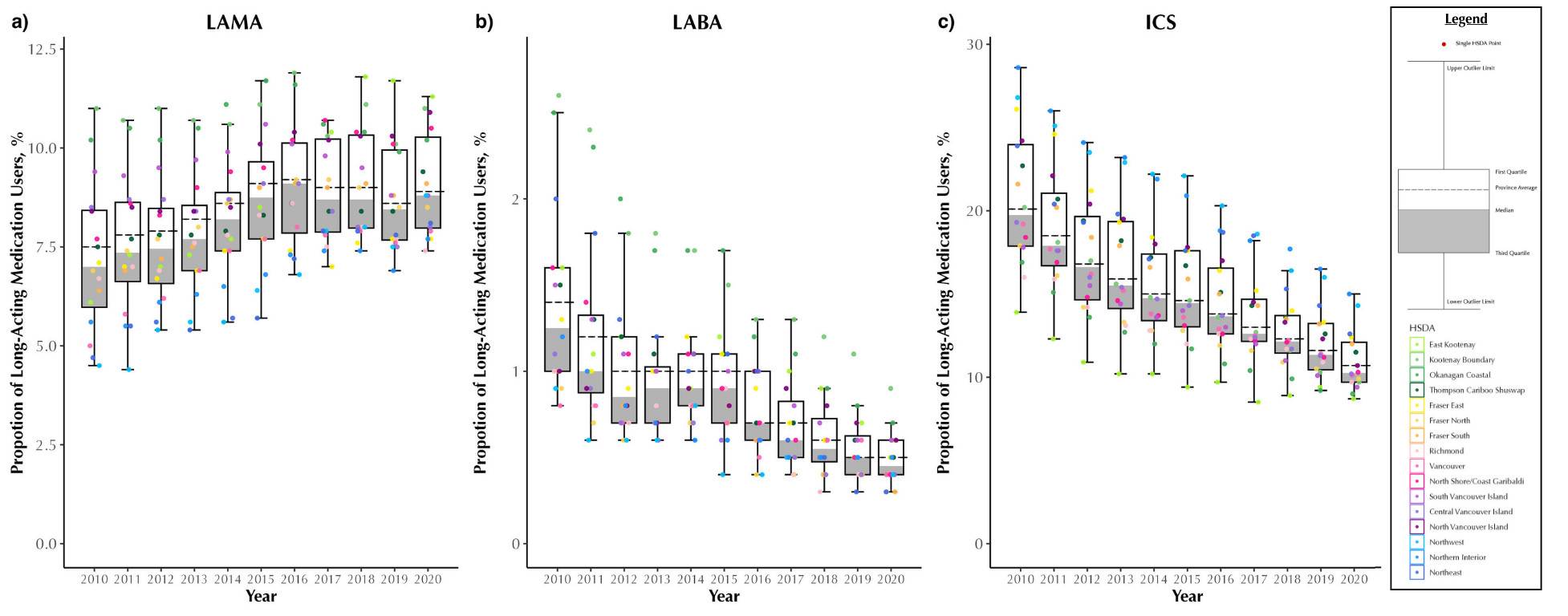

### **Figure A10: Proportion of single therapy long-acting medication users (%, among COPD patients) of LAMA (a), LABA (b) and ICS (c) in British Columbia, Canada, from 2001 to 2020, stratified by Health Service Delivery Areas (HSDAs)**

Abbreviations: COPD: chronic obstructive pulmonary disease, ICS: Inhaled Corticosteroids, LABA: Long-Acting Beta Agonists, LAMA: Long-Acting Muscarinic Antagonists; Q1: quartile 1; Q3: quartile 3 Note: Each dot represents an HSDA. The horizontal line cutting through the plot is the overall provincial average. The median is the line separating the upper (white) and lower (dark grey) boxes

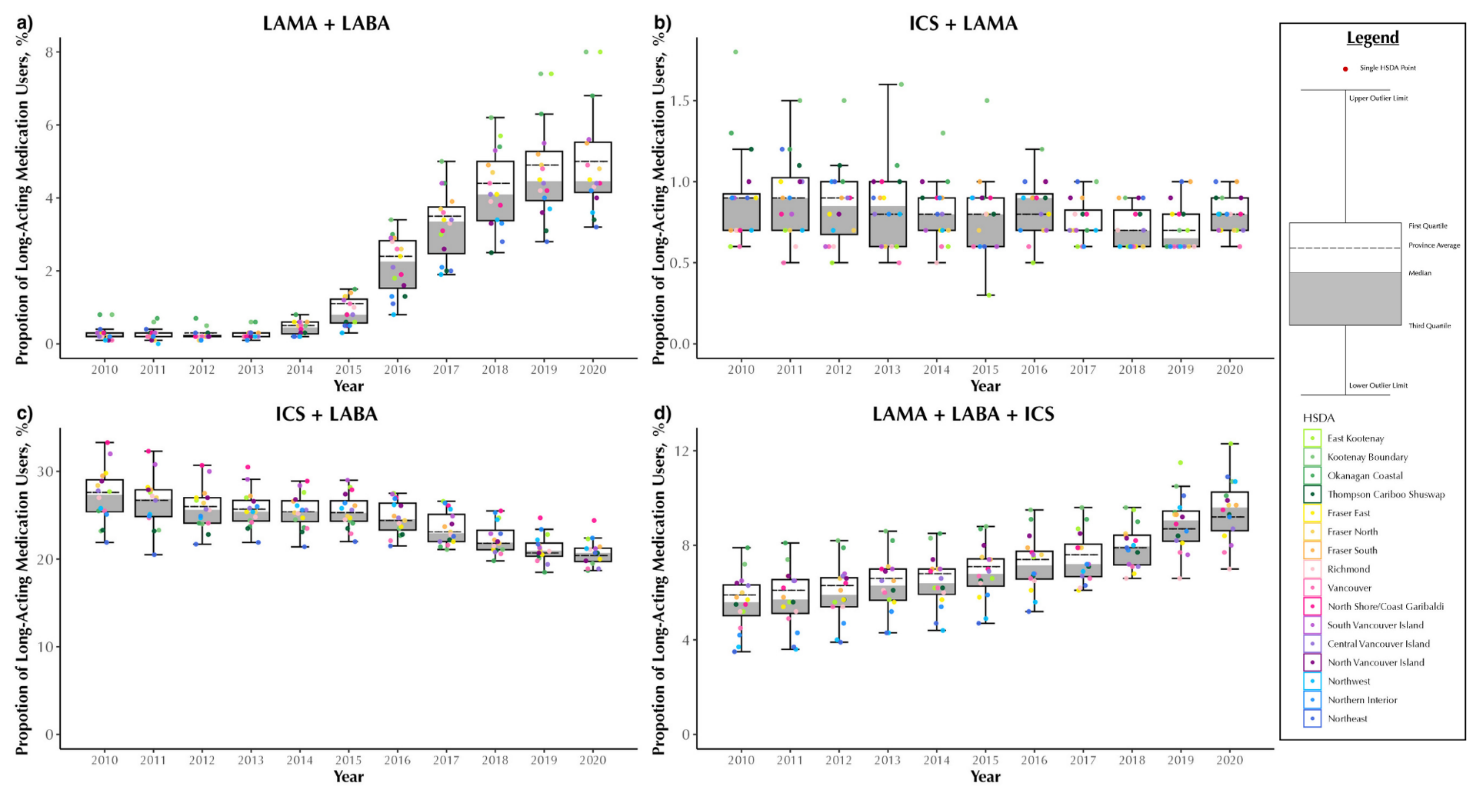

### **Figure A11: Proportion of combination therapies long-acting medication users (%, among COPD patients) of LAMA and LABA (a), ICS and LAMA (b), ICS and LABA (c) and ICS and LAMA and LABA (d) in British Columbia, Canada, from 2001 to 2020, stratified by Health Service Delivery Areas (HSDAs)**

Combination therapies are based on either single inhalers containing multiple ingredients or from separate inhalers with 14 days of overlap. Abbreviations: COPD: chronic obstructive pulmonary disease, ICS: Inhaled Corticosteroids, LABA: Long-Acting Beta Agonists, LAMA: Long-Acting Muscarinic Antagonists; Q1: quartile 1; Q3: quartile 3 Note: Each dot represents an HSDA. The horizontal line cutting through the plot is the overall provincial average. The median is the line separating the upper (white) and lower (dark grey) boxes
